# Rare neurological and neurodevelopmental variants in ALS link to onset, survival and family history

**DOI:** 10.64898/2026.06.09.26354977

**Authors:** Ciara O’Donoghue, Emrah Kacar, Tomas Gomes, Emmet Costello, Niall Pender, Colm Peelo, Marie Ryan, Mark Heverin, Susan Byrne, Peter Bede, Orla Hardiman, Russell Lewis McLaughlin, Ross Patrick Byrne

**Affiliations:** Smurfit Institute of Genetics, Trinity College Dublin, Ireland, D02 VF25; Academic Unit of Neurology, Trinity Biomedical Sciences Institute, Trinity College Dublin, Dublin, Ireland, D02 R590; Department of Neurology, Beaumont Hospital, Dublin, Ireland, D09 V2N0; Department of Psychology, Beaumont Hospital, Dublin, Ireland, D09 V2N0; Department of Clinical Genetics and Department of Neurology, Children’s Health Ireland (CHI) at Crumlin, Dublin, Ireland, D12 N512; Computational Neuroimaging Group, School of Medicine, Trinity College Dublin, Dublin, Ireland, D02 R590; Department of Neurology, St. James’s Hospital, Dublin, Ireland, D08 NHY1

**Author notes:** Correspondence to: Ciara O’Donoghue Full address Smurfit Institute of Genetics, Trinity College Dublin, Ireland, D02 VF25.

**Keywords:** Amyotrophic Lateral Sclerosis (ALS), pleiotropy, annotation, Whole genome sequencing (WGS), family history, phenotype

## Abstract

**Background:** Neurological, neuropsychiatric, and neurodevelopmental disorders cluster in ALS families, sharing a common genetic architecture with ALS. Pathogenic variants in genes associated with other neurological, neurodevelopmental, or neuropsychiatric disorders may also co-occur in ALS and modify phenotype. We have sought to determine the prevalence and clinical pattern of likely-pathogenic/pathogenic (LP/P) non-ALS neurological, neurodevelopmental, and neuropsychiatric variants, alone and in combination with ALS-gene variants, in two large ALS cohorts.

**Methods:** Whole-genome sequencing (WGS) of 469 Irish and 774 Answer ALS people with ALS (pwALS) was analysed for ClinVar LP/P variants associated with other neurological (*n* = 15541), neurodevelopmental (*n* = 9761), and neuropsychiatric (*n* = 321) phenotypes. Inheritance patterns for associated genes (autosomal recessive/autosomal dominant) along with the associated phenotype were validated using OMIM. Standardised clinical data included family history, site and age of onset, El Escorial category, survival, motor decline, and cognitive and behavioural assessments. Known ALS-gene variants and *C9orf72* repeat expansion status were included for each cohort.

**Results:** Non-ALS neurological variants were identified in 47/469 (10.0%) Irish and 69/774 (8.9%) Answer ALS participants, most frequently in hereditary spastic paraplegia-associated genes (3.2% Irish; 2.8% Answer ALS). Irish neurological variant carriers showed higher frequency of respiratory onset (10.6% vs 1.2%, Fisher’s exact *p* = 0.002, Φ = 0.20) and fewer premorbid behavioural symptoms (0.92 ± 0.56 vs 3.08 ± 0.97, Cohen’s *d* = −0.40). Neurodevelopmental variants occurred in 12/469 (2.6%) Irish and 20/774 (2.6%) Answer ALS participants. In the Irish cohort, neurodevelopmental variant carriers had significantly shorter survival in Cox proportional hazards model (log-rank *p* = 0.005), corresponding to a more than two-fold increased hazard of death (HR = 2.25, 95% CI 1.26-4.00), and had significantly increased familial burden of neuropsychiatric disorders among first- and second-degree relatives (negative binomial IRR for carriers = 2.41, 95% CI: 1.12-5.18, *p* = 0.025). Across combined cohorts, 18 individuals (Irish *n* = 8; Answer ALS *n* = 10) carried ≥2 LP/P variants spanning ALS and non-ALS genes.

**Conclusion:** Rare LP/P variants in genes associated with other neurological and neurodevelopmental disorders occur in up to 12% of pwALS across two independent cohorts. Carriers show distinct phenotypes, shorter survival, and characteristic family history patterns. These findings suggest that extended pleiotropic and oligogenic architectures may contribute to ALS heterogeneity.

## Introduction

Amyotrophic lateral sclerosis (ALS) is a clinically and genetically heterogeneous disease. Patterns of motor weakness, cognitive or behavioural involvement, and rates of progression are variable across people with ALS (pwALS), and this phenotypic variability is incompletely explained by known ALS-associated mutations. Rare variants may play an important but underestimated role in ALS,^1^ as suggested by substantial differences in heritability estimates between epidemiological (50-60%)^2,3^ and SNP-based studies (3-10%).^4,5^ Multiple factors contribute to ALS genetic architecture, including oligogenic inheritance,^6,7^ incomplete penetrance,^8^ and polygenic influences,^4,9,10^ making it more complex than single-gene Mendelian models alone. Adding to this complexity, large-scale genetic studies show that ALS risk loci sit within broader neurodegenerative^11^ and neuropsychiatric^4,9^ networks, rather than a clearly defined, disease-specific set of genes.

Higher burdens of neurodegenerative, neuropsychiatric, and neurodevelopmental disorders are found within ALS kindreds,^12–14^ consistent with shared polygenic liability across these traits.^4,11,15^ In accordance with this, rare variants in genes primarily associated with other neurodegenerative^11^ and neuromuscular disorders^16^ may influence ALS risk and presentation, while, conversely, ALS genes such as *C9orf72*, *FUS*, *TBK1*, and *KIF5A*^17–19^ appear in gene sets for dementia, parkinsonism, and psychiatric diseases.^20^ Cross-disorder GWAS has also shown considerable overlap among common variants associated with neuropsychiatric and neurodevelopmental conditions.^21^ Within this context, schizophrenia polygenic risk scores account for a modest proportion of variance between ALS cases and controls,^4^ suggesting that rare genetic variants from neuropsychiatric disorders may also contribute to ALS risk. Taken together, these observations underscore genetic pleiotropy across neurological and neuropsychiatric disorders^15,21,22^ and further support the presence of shared genetic backgrounds across these disease groups.

The *C9orf72* repeat expansion provides an example of such pleiotropy by its association with both ALS and frontotemporal dementia along a clinicopathological continuum.^23^ Beyond the ALS-FTD spectrum, *C9orf72* expansions have also been reported in individuals with parkinsonism and other movement disorders,^24^ as well as in a range of psychiatric^25,26^ and neurodevelopmental presentations.^25,27,28^ Although cognitive and behaviour change occurs more frequently in those carrying the *C9orf72* repeat expansion,^29,30^ extra-motor features are also detectable in pwALS in the absence of the *C9orf72* repeat expansion,^31,32^ and increased rates of psychiatric and neurodevelopmental disorders are observed in ALS families irrespective of *C9orf72* status.^13,14^ Such findings indicate that neuropsychiatric and neurodevelopmental variants are likely to influence ALS risk beyond well-characterised monogenic loci, and that the occurrence of neuropsychiatric diagnoses within ALS kindreds are likely to reflect genetic pleiotropy rather than coincidental comorbidity.

Converging evidence supports an oligogenic mode of inheritance in a subgroup of pwALS, characterised by the presence of multiple rare variants within ALS-associated genes.^6,7^ Extending this concept, it is plausible that variants in genes primarily implicated in other neurological, neuropsychiatric, and neurodevelopmental diseases could occur alongside ALS mutations more often than expected by chance, acting as modifiers within shared disease networks rather than as isolated monogenic causes. However, there is limited information regarding the prevalence, spectrum, and phenotypic consequences of non-ALS genetic variants in ALS populations, particularly within neurodevelopmental and neuropsychiatric gene sets.

Here we explore the presence and relevance of rare variants in genes primarily associated with other neurological, neuropsychiatric (NP), and neurodevelopmental (ND) disorders in pwALS. To do this, we analysed whole-genome sequencing data (WGS) from two large ALS cohorts, including cases from the Irish ALS Register^33^ (sequenced as part of Project MinE^34^) and participants from Answer ALS.^35^ We systematically annotated ClinVar^-^reported^36^ likely-pathogenic/pathogenic (LP/P) variants linked to non-ALS neurological, neuropsychiatric, and neurodevelopmental phenotypes relevant to ALS, as identified through a focused literature review. By combining variant-level annotation with detailed clinical, genetic, and family history data, this work aims to i) quantify the burden and spectrum of non-ALS LP/P variants in pwALS, ii) examine their associations with motor, cognitive, behavioural, and survival phenotypes, and iii) provide a curated catalogue of pleiotropic variants and oligogenic carriers to inform future rare-variant burden analyses in ALS.

## Materials and methods

### Study Population

ALS cases from the Irish ALS register^33^, comprising those with Possible, Probable, and Definite ALS using the El Escorial Research Criteria, and Answer ALS^35^ with whole-genome sequencing (WGS) data were included. Primary lateral sclerosis, FTD-only diagnoses, and mimics (e.g., hereditary spastic paraplegia) were excluded. Phenotypes inconsistently recorded in free-text fields (e.g., flail arm variant) were omitted to minimise misclassification bias. For both ALS cohorts, carrier status of ALS-associated mutations was derived from existing cohort-level genetic reports. *C9orf72* expansion status was estimated by repeat-primed PCR as described elsewhere (ref PMID 21944779) and defined in Irish as positive if ≥30 discernible repeats were present, intermediate for 17-29 repeats, and negative if <17. In Answer ALS, a positive result was defined as ≥26 repeats.^35^ Carriers of ALS variants were included in analysis.

#### Irish ALS Cohort

Irish WGS data was available for 469 pwALS (individual VCF files; PCR-free library preparation; genomic DNA extracted from peripheral blood). Phenotype and family history data were extracted from the Irish ALS Register (REDCap)^33^ and included site and age of onset, survival (symptom onset to death, months), El Escorial category, Edinburgh Cognitive and Behavioural ALS Screen (ECAS),^30^ and Beaumont Behavioural Inventory (BBI).^38^ ECAS and BBI abnormality was defined using validated, demographically adjusted thresholds.^30,38^ WGS from 230 Irish population controls was available to estimate population-specific carrier frequencies and for enrichment analysis.

#### Answer ALS Cohort

Answer ALS data (Version 3, 2024) included 774 ALS participants with WGS (joint VCFs; TruSeq, PCR-free, peripheral blood). Phenotype data included site and age of onset, survival (symptom onset to death, months), El Escorial category, and ALS Cognitive Behavioural Screen (ALS-CBS).^39^ ALS-CBS abnormality was defined as any visit with a total cognitive score ≤ 16 (ALSci/possible FTLD) or behavioural subscale score ≤36 (ALSbi/possible FTLD-behavioural).^40^ In addition, WGS from 94 Answer ALS controls was available to estimate population-specific carrier frequencies and for enrichment analysis.

#### Variant Calling

Variants in Answer ALS and Irish data were called against the human reference genome build GRCh38. Answer ALS used Senteion for joint calling with variant quality score recalibration run using GATK (truth sensitivity level = 99.0), ensuring only high quality variants were included. Irish variants were called as part of Project MinE (ref PMID 29955173) using GATK haplotype caller. Called variants were additionally filtered to minimum variant and genotype qualities of 20. Variants were annotated using bcftools^37^ with the ClinVar^36^ GRCh38 variant callset as a reference.

### Literature Review and Phenotype Selection

A literature review was conducted to identify neurological, neuropsychiatric, and neurodevelopmental phenotypes associated with ALS, either through co-aggregation or genetic overlap. The identified phenotypes were used as ClinVar search terms. Neurological phenotypes searched in ClinVar included Alzheimer’s disease, cerebellar ataxia, Charcot-Marie-Tooth disease, cognitive impairment, hereditary spastic paraplegia, Lewy body dementia, multiple sclerosis, multiple system atrophy, Parkinson’s disease, progressive supranuclear palsy, peripheral neuropathy, inclusion body myopathy, and multisystem proteinopathy. Neuropsychiatric and neurodevelopmental traits searched in ClinVar included anxiety, depression, bipolar disorder, psychosis, schizophrenia, alcohol use disorder, attention-deficit hyperactivity disorder, autism spectrum disorder, speech and language delay, and intellectual disability. All ClinVar phenotypic search terms, along with the references and rationale for inclusion, are available in Supplementary Table 1.1.

### ClinVar Variant Annotation

ClinVar^36^ GRCh38 VCFs (downloaded 18-June-13 July 2025; updated April 2026) were filtered to ≥1 star expert-reviewed “likely pathogenic”/“pathogenic” (LP/P) variants matching selected phenotypes yielding 15541 neurological, 321 psychiatric, and 9761 neurodevelopmental variants. A carrier was defined as possessing ≥1 variant from the neurological, neuropsychiatric, or neurodevelopmental ClinVar search lists. All observed carriers were heterozygous and carried autosomal variants. The genes relating to each variant were reviewed in OMIM to document mode of inheritance (autosomal recessive versus autosomal dominant) and to confirm associated phenotypes. For several neurodevelopmental genes, the limited information on OMIM was resolved by searching SFARI Gene and GeneCards databases. Variants were excluded from analysis if from genes where the phenotype recorded in ClinVar differed from all other genetic databases. PwALS carrying variants in non-ALS genes with an autosomal dominant pattern were reviewed by a consultant neurologist to confirm a clinical diagnosis of ALS and exclude ALS mimics.

Variants in known ALS genes (e.g., *FIG4*) were retained if ClinVar annotation only demonstrated non-ALS phenotypes for the specific variant. Entries annotated “pathogenic” or “likely pathogenic” for intellectual disability or syndromic neurodevelopmental phenotypes were retained as they may manifest as isolated neuropsychiatric traits or act as oligogenic modifiers rather than monogenic ALS cause.^22,58^

Allele frequencies for each variant were obtained from gnomAD v4.0,^59^ using the non-Finnish European population where possible to reflect the vast majority of the Irish and Answer ALS cohorts. Where available, allele frequencies from TOPMed^60^ and the 1000 Genomes Project^61^ were also recorded for reference. All reported allele frequencies for neurological and neurodevelopmental variants identified during annotation are provided in Supplementary Tables 3.1 and 4.1, respectively.

### Statistical Analysis

#### Software and General Approach

All analyses were conducted in R version 4.1.2. Demographic and clinical variables were compared between cohorts (Irish versus Answer ALS) and between variant carriers and non-carriers within each cohort, with statistical tests, p-values and effect sizes reported in Supplementary Table S2.1-5.

#### Cohort-Level Descriptive Comparisons

For overall cohort comparisons between Irish and Answer ALS ALS cases (Supplementary Table S2.1), categorical variables (biological sex, site of onset, El Escorial category, family history, ethnicity, and *C9orf72* repeat expansion status) were analysed using χ² tests when all expected counts were ≥5, Yates’ continuity correction was added when any expected count was 5-9, and Fisher’s exact tests were used when any expected cell count was <5. Continuous variables (age at symptom onset, survival from symptom onset to death, and ALSFRS-R baseline score) were compared using Welch’s two-sample t-tests. For 2⋅2 tables, Φ is reported as the effect size; while variables with >2 categories Cramér’s V is used. For continuous outcomes, effect size is summarised as Cohen’s *d*. Across 12 prespecified cross-cohort comparisons, a Bonferroni-corrected significance threshold of p<0.0042 was applied; a p-value <0.05 but above Bonferroni threshold was considered nominally significant.

#### Carrier versus Non-Carrier Clinical Comparisons

Within each cohort, clinical features of ClinVar-LP/P neurological and neurodevelopmental variant carriers were compared with non-carriers (Supplementary Table S2.2 and Supplementary Table S2.3, respectively). Categorical variables (sex, site of onset, El Escorial category, family history, ethnicity, abnormal ECAS, BBI, or CBS scores) were analysed using χ² or Fisher’s exact tests as above, with Φ and Cramér’s V reported as effect sizes. Continuous variables (age at onset, survival, ALSFRS-R, ECAS, BBI, and CBS baseline scores) were compared using Welch’s t-tests with Cohen’s *d* reported. Because clinical data were incomplete for some participants, the total *n* contributing to each clinical feature is shown in the tables and may differ from the total cohort size. For carrier vs non-carrier analyses, 19 independent comparisons in the Irish cohort and 17 in the Answer ALS were prespecified. Bonferroni-corrected thresholds of p<0.00263 (Irish) and p<0.00294 (Answer ALS) were used, and p-values between 0.05 and the cohort-specific Bonferroni threshold are described as nominally significant.

#### Family History Data Processing

Complete family history data for analyses were available from the Irish ALS Register (*n*=455). Answer ALS family history data were excluded from analysis as all family history categories, including ALS and FTD, had significantly higher frequency in Irish probands and family size was unavailable in Answer ALS. Only first- and second-degree relatives were included in family history analysis as previously published.^13,14^ Baseline family size was calculated as the sum of reported brothers, sisters, paternal uncles and aunts, and maternal uncles and aunts. Individuals with incomplete family history forms or with zero family size were excluded from analysis. Neurological diseases reported in family members included Alzheimer’s disease, unspecified dementia, multiple sclerosis, Parkinson disease, and ‘other neurological’ conditions. Neuropsychiatric diseases included autism, bipolar disorder, depression, psychosis, schizophrenia, suicide, alcoholism, intravenous drug abuse, and ‘other neuropsychiatric’ conditions. Affected relative counts were binary-coded per relative (“Checked” vs not) and summed by individual. Proportions were derived as counts divided by family size. Wilcoxon rank-sum tests compared absolute counts, proportions of affected relatives, and family sizes between carriers and non-carriers due to right-skewed distributions. Negative binomial regression modelled counts of affected relatives (Poisson offset by log family size). Incidence rate ratios (IRRs) with 95% confidence intervals quantified carrier enrichment or depletion. Analyses were independent by carrier group (neurological and neurodevelopmental).

#### Survival Analysis

Survival was defined as time from symptom onset to death, with mortality in Answer ALS updated using version 4 of the clinical dataset (Supplementary Table S2.3-4). We initially screened for survival differences using a Welch’s t-test between carriers and non-carriers, and then performed formal survival modelling on neurodevelopmental (ND) variant carriers to validate initial screening results. Kaplan-Meier curves and log-rank tests were used to compare survival between ND variant carriers and non-carriers within and across cohorts. Cox-proportional hazards models were fitted with carrier status as the main exposure and cohort was included as an adjustment variable. Hazard ratios (HRs) with 95% confidence intervals are reported. In the Irish cohort, site of onset (bulbar vs non-bulbar) was added as a covariate as a higher trend of bulbar-onset was noted in ND carriers, which has been associated with shorter survival.^62^ We assessed the proportional hazards assumption using Schoenfeld residuals (see Supplementary Methods; S1). As the proportional hazards assumption was violated for “Cohort”, we used a stratified Cox model allowing the Irish and Answer ALS cohorts to have separate baseline hazard functions, while estimating a single hazard ratio for carrier status across both groups. We also tested carrier status ⋅ cohort interaction terms to assess whether the effect of carrier status differed between cohorts.

Although our primary analysis focused on rare ClinVar LP/P variants, the available sample size and modest carrier counts limited power to detect case-control enrichment. Exploratory rare-variant burden testing using Sequence Kernel Association Test (SKAT/SKAT-O) across neurological and neurodevelopmental variant sets did not identify significant enrichment and are described in the Supplementary Methods and Supplementary Results (S1).

## Ethics Approval

This study was conducted in accordance with the declaration of Helsinki. All participants gave informed consent. The study was approved by the Beaumont Hospital Ethics (Medical Research) Committee (REF 15/40). Answer ALS was approved by the Johns Hopkins institutional review board (nos. 00082277 and 00240000). The Answer ALS program is registered at clinicaltrials.gov (NCT02574390).

## Data availability

Irish genetic data available on reasonable request from project MinE (https://projectmine.com/datasharing/). Data from Answer ALS available through application (https://dataportal.answerals.org/home).

## Results

### Cohort Demographics

Demographic and clinical characteristics for pwALS with WGS from the Irish (*n* = 469) and Answer ALS (*n* = 774) cohorts are summarised in Table 1. After Bonferroni correction for 12 independent comparisons (*p* < 0.0042), cohorts differed significantly in El Escorial category, with a higher proportion of “Definite ALS” in the Irish cohort (54.4% vs 30.0%; *p* = 2.2e-16, Cramér’s V=0.15), and higher family history rate in all categories including ALS (20.7% vs. 13.3%; *p* = 0.001, *Φ* = 0.09), FTD (3.5% vs. 0.6%; Fisher’s exact *p* = 0.0003, *Φ* = 0.11), other neurological disease (51.4% vs. 42.0%; *p* = 0.002, *Φ* = 0.09), and psychiatric disorders (54.9% vs. 10.7%; *p* < 2.2e-16, *Φ* = 0.48) in the Irish cohort. Irish pwALS had older age of onset (62.96 ± 1.02 vs. 56.54 ± 0.82; *p* = 4.26e-18, Cohen’s *d* = 0.57), and higher ALSFRS-R baseline scores (36.72 ± 0.71 vs. 33.97 ± 0.61; *p* = 1.17e-08, Cohen’s *d* = 0.35) compared to Answer ALS. All Irish participants were White Non-Hispanic versus 88.1% in Answer ALS (Fisher’s exact *p* < 2.2e-16, *Φ* = 0.22).

**Table 1.**
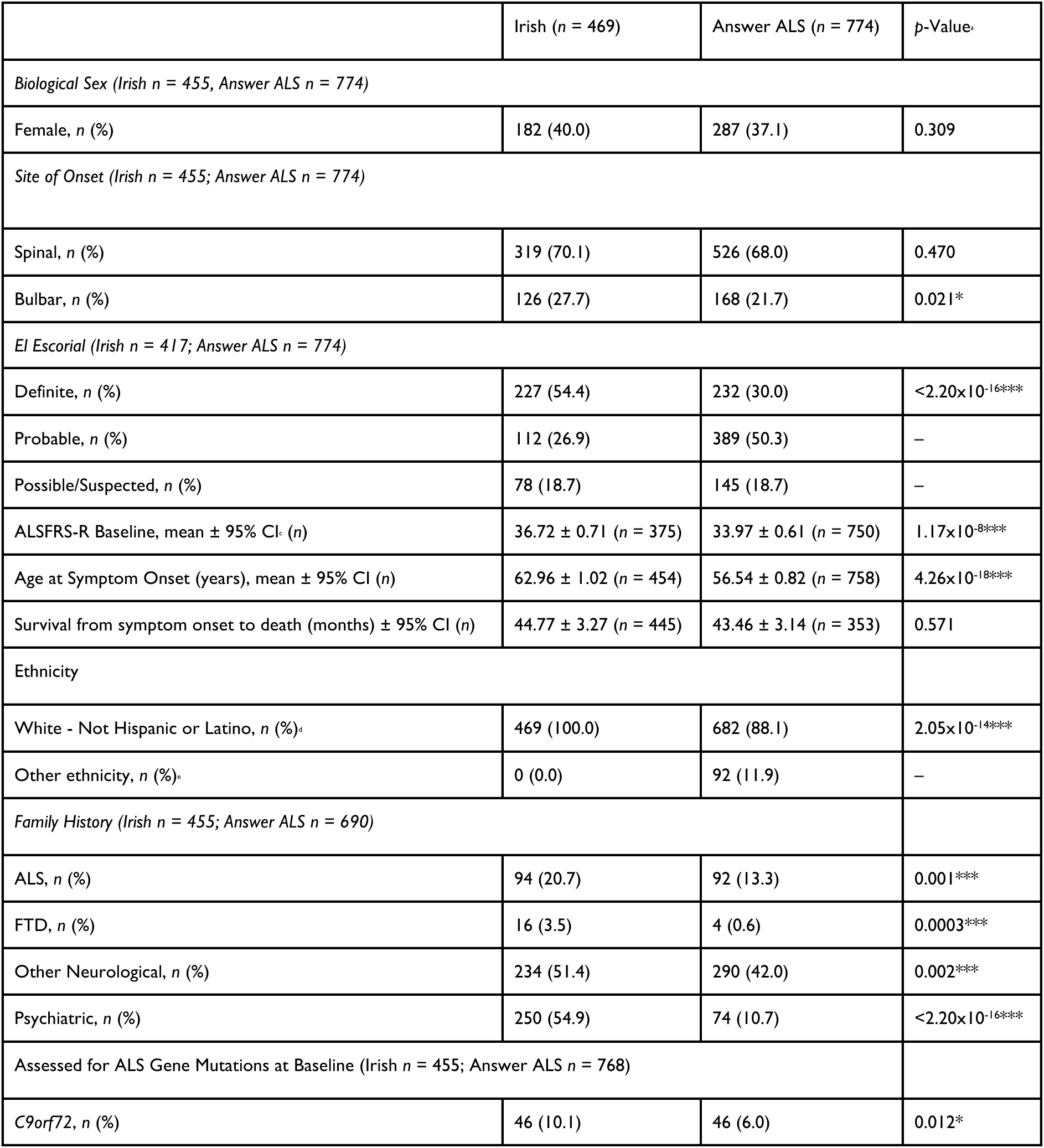
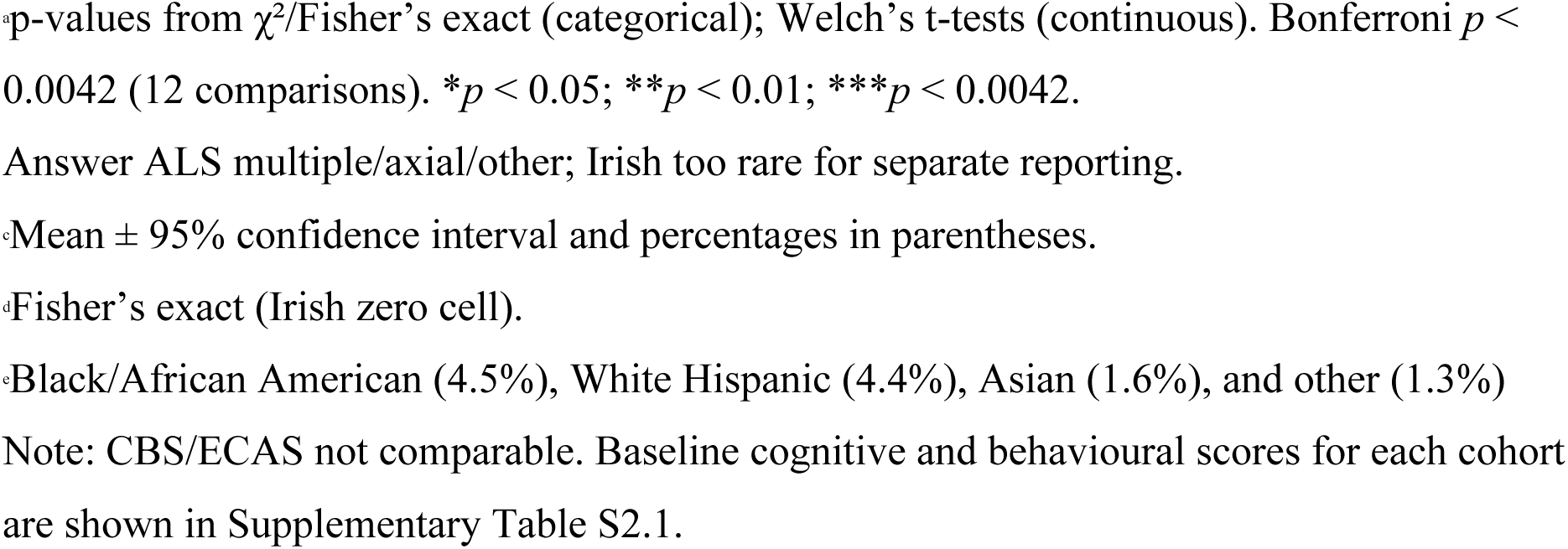
Demographic, clinical, and cognitive and behavioural characteristics of pwALS with whole-genome sequencing (WGS) data from the Irish and Answer ALS cohorts.

|  | Irish (n = 469) | Answer ALS (n = 774) | p-Value |
| --- | --- | --- | --- |
| *Biological Sex (Irish n = 455; Answer ALS n = 774)* |  |  |  |
| Female, n (%) | 182 (40.0) | 287 (37.1) | 0.309 |
| *Site of Onset (Irish n = 455; Answer ALS n = 774)* |  |  |  |
| Spinal, n (%) | 319 (70.1) | 526 (68.0) | 0.470 |
| Bulbar, n (%) | 126 (27.7) | 168 (21.7) | 0.021* |
| *El Escorial (Irish n = 417; Answer ALS n = 774)* |  |  |  |
| Definite, n (%) | 227 (54.4) | 232 (30.0) | $<2.20 \times 10^{-16}***$ |
| Probable, n (%) | 112 (26.9) | 389 (50.3) | – |
| Possible/Suspected, n (%) | 78 (18.7) | 145 (18.7) | – |
| ALSFRS-R Baseline, mean ± 95% CI (n) | 36.72 ± 0.71 (n = 375) | 33.97 ± 0.61 (n = 750) | $1.17 \times 10^{-8}***$ |
| Age at Symptom Onset (years), mean ± 95% CI (n) | 62.96 ± 1.02 (n = 454) | 56.54 ± 0.82 (n = 758) | $4.26 \times 10^{-18}***$ |
| Survival from symptom onset to death (months) ± 95% CI (n) | 44.77 ± 3.27 (n = 445) | 43.46 ± 3.14 (n = 353) | 0.571 |
| *Ethnicity* |  |  |  |
| White - Not Hispanic or Latino, n (%) | 469 (100.0) | 682 (88.1) | $2.05 \times 10^{-14}***$ |
| Other ethnicity, n (%) | 0 (0.0) | 92 (11.9) | – |
| *Family History (Irish n = 455; Answer ALS n = 690)* |  |  |  |
| ALS, n (%) | 94 (20.7) | 92 (13.3) | 0.001*** |
| FTD, n (%) | 16 (3.5) | 4 (0.6) | 0.0003*** |
| Other Neurological, n (%) | 234 (51.4) | 290 (42.0) | 0.002*** |
| Psychiatric, n (%) | 250 (54.9) | 74 (10.7) | $<2.20 \times 10^{-16}***$ |
| *Assessed for ALS Gene Mutations at Baseline (Irish n = 455; Answer ALS n = 768)* |  |  |  |
| *C9orf72*, n (%) | 46 (10.1) | 46 (6.0) | 0.012* |

p-values from $\chi^2$ /Fisher's exact (categorical); Welch's t-tests (continuous). Bonferroni $p < 0.0042$ (12 comparisons). \* $p < 0.05$ ; \*\* $p < 0.01$ ; \*\*\* $p < 0.0042$ .
Answer ALS multiple/axial/other; Irish too rare for separate reporting.
Mean ± 95% confidence interval and percentages in parentheses.
Fisher's exact (Irish zero cell).
Black/African American (4.5%), White Hispanic (4.4%), Asian (1.6%), and other (1.3%)
Note: CBS/ECAS not comparable. Baseline cognitive and behavioural scores for each cohort are shown in Supplementary Table S2.1.

Nominally significant differences (*p* < 0.05) included a higher proportion of bulbar onset (*p* = 0.0211, *Φ* = 0.07) and *C9orf72* repeat expansion carriers (*p* = 0.012, Φ = 0.07) in the Irish data. Biological sex and survival did not differ significantly between cohorts.

Detailed summary statistics results for cohort analyses are summarised in Supplementary Table S2.1.

### Neurological Variants in Irish and Answer ALS

ClinVar LP/P neurological-associated variants were identified in a subset of pwALS in both cohorts (Table 2). In the Irish cohort, 47 of 469 individuals (carrier frequency 10.0%) carried such variants across 26 genes including *PRKN* (*n* = 6), *ANO10* (*n* = 4), *POLG* (*n* = 4), *MPO* (*n* = 3), *SORD* (*n* = 3), *AP5Z1* (*n* = 2), *FARS2* (*n* = 2), *FBXO7* (*n* = 2), *FIG4* (*n* = 2), *MFN2* (*n* = 2), *SPG7* (*n* = 2), *AMPD2* (*n* = 1), *AP4M1* (*n* = 1), *AP4S1* (*n* = 1), *CYP7B1* (*n* = 1), *GBA1* (*n* = 1), *IGHMBP2* (*n* = 1), *MSTO1* (*n* = 1), *NEMF* (*n* = 1), *NUDT2* (*n* = 1), *PLA2G6* (*n* = 1), *POLR3B* (*n* = 1), *RNF216* (*n* = 1), *SH3TC2* (*n* = 1), *SPG11* (*n* = 1), *SURF1* (*n* = 1). Although *FIG4* and *SPG11* have been implicated in ALS, the specific variants identified here are classified as pathogenic in ClinVar for other neurological phenotypes rather than ALS/MND/FTD. In Answer ALS, 69 of 774 individuals (carrier frequency 8.9%) carried variants in 29 genes including *SORD* (*n* = 13), *POLG* (*n* = 11), *GBA1* (*n* = 7), *PRKN* (*n* = 6), *SPG7* (*n* = 6), *DST* (*n* = 2), *IGHMBP2* (*n* = 2), *MPO* (*n* = 2), *PRX* (*n* = 2), *VPS13C* (*n* = 2), *AARS1* (*n* = 1), *AP4B1* (*n* = 1), *AP4M1* (*n* = 1), *DDHD1* (*n* = 1), *DNAJC30* (*n* = 1), *FA2H* (*n* = 1), *FBXO7* (*n* = 1), *LRRK2* (*n* = 1), *MTRFR* (*n* = 1), *PLEKHG5* (*n* = 1), *PNPLA6* (*n* = 1), *POLR3B* (*n* = 1), *PSAP* (*n* = 1), *RFC4* (*n* = 1), *RNU12* (*n* = 1), *SH3TC2* (*n* = 1), *SURF1* (*n* = 1), *TTN* (*n* = 1), *VWA1* (*n* = 1). Heatmaps detailing the distribution of variants across associated genes, carrier frequencies, and associated neurological phenotypes are displayed in Figure 1. General population allele frequencies from gnomAD ranged from 9.0×10^-6^ to 4.8×10^-3^ (estimated carrier frequency about 1 in 55,000 to 1 in 100). Out of the 43 unique genes identified through variant annotation from both cohorts, 8 (18.6%) exhibited an autosomal dominant inheritance pattern on OMIM (*GBA1, LRRK2, PSAP, POLR3B, MFN2, AARS1, MSTO1, MPO*). ALS cases from both cohorts carrying these variants (21/116, 18.1% of total carrier cohort) were reviewed by a consultant neurologist, who confirmed their condition as probable or definite ALS based on available clinical data and unlikely to be ALS mimics. Thus, these individuals were retained in subsequent analysis.

**Figure 1.**
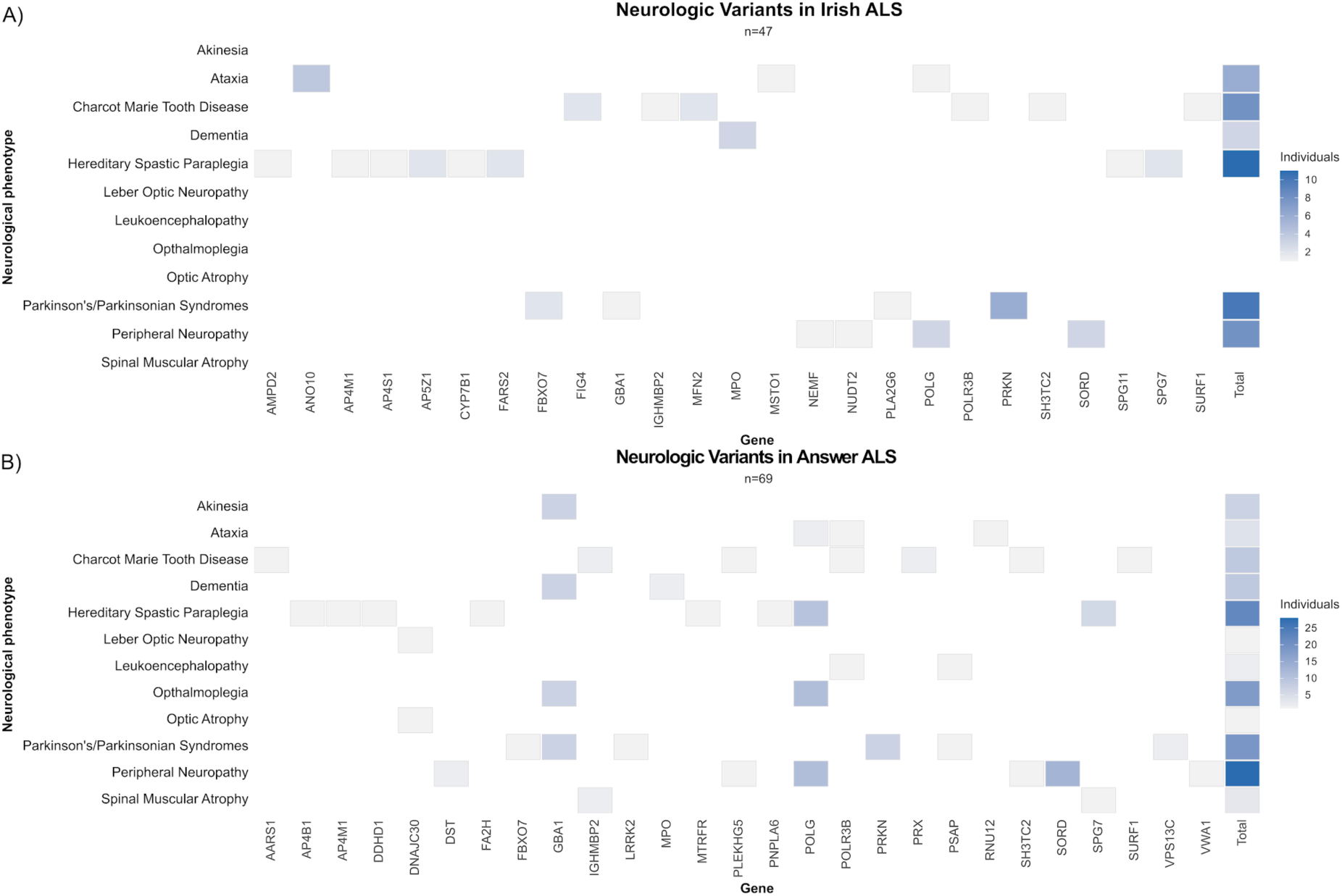
Genotype-phenotype heatmaps for LP/P neurological variants in Irish and Answer ALS pwALS. Each row represents a neurological phenotype associated with the annotated variants while each column represents a gene associated with each variant in A) Irish (*n* = 47) and B) Answer ALS (*n* = 69) cohorts. Variants linked to multiple phenotypes are accounted for. Each individual is counted only once per gene-phenotype cell as all individuals were heterozygous. The rightmost ‘Total’ column shows, for each phenotype, the total number of individuals with any LP/P neurological variant across all genes.

**Table 2.** Demographic and clinical characteristics of pwALS with neurological variants in the Irish and Answer ALS cohorts.

| | Irish ($n = 47$) | Answer ALS ($n = 69$) |
| --- | --- | --- |
| *Biological Sex* |  |  |
| Female, $n$ (%) | 16 (34.0) | 20 (28.9) |
| *Site of Onset* |  |  |
| Spinal, $n$ (%) | 33 (70.2) | 47 (68.1) |
| Bulbar, n (%) | 9 (19.1) | 17 (24.6) |
| *El Escorial (Irish n = 45, AnswerALS n = 73)* |  |  |
| Definite, n (%) | 22 (48.9) | 23 (31.5) |
| Probable, n (%) | 10 (22.2) | 32 (46.6) |
| Possible, n (%) | 12 (26.7) | 11 (15.9) |
| ALSFRS-R Baseline ± 95% CI (n) | 36.85 ± 2.22 (±6.03%) (n = 39) | 35.33 ± 2.21 (n = 69) |
| *Cognitive and Behavioural Measures* |  |  |
| ECAS ever abnormal, n/N (%) | 6/22 (27.3) | – |
| Pre-MND BBI ever abnormal, n/N (%) | 2/28 (7.14) | – |
| Post-MND BBI ever abnormal, n/N (%) | 14/28 (50.0) | – |
| CBS cognitive ever abnormal, n/N (%) |  | 27/61 (44.3) |
| CBS behavioural ever abnormal, n/N (%) |  | 16/50 (32.0) |
| Age at Symptom Onset (Years), mean ± 95% CI (n) | 62.25 ± 3.44 (n = 47) | 56.20 ± 2.63 (n = 69) |
| Survival (Months), mean ± 95% CI (n) | 49.87 ± 10.03 (n = 47) | 44.37 ± 9.44 (n = 37) |

\*Ns added per cell to reflect available data if some is missing.

In the Irish cohort, neurological variant carriers had a significantly higher proportion of respiratory onset compared with non-carriers (10.6% (*n* = 5/47) vs 1.2% (*n* = 5/408); Fisher’s exact *p* = 0.002). Variant carriers also had significantly lower Pre-MND BBI baseline scores (0.92 vs 3.08; *p* = 0.0003, Cohen’s *d* = −0.40) indicating fewer premorbid behavioural changes, and lower Post-MND BBI baseline scores (5.50 vs 9.78; *p* = 0.004, Cohen’s *d* = - 0.36325) indicating fewer behavioural changes after ALS onset at baseline assessment. The Pre-MND BBI association meets the Bonferroni-corrected threshold, while the Post-MND BBI findings are nominally significant and thus should be interpreted cautiously. Other phenotypic features in the Irish cohort, including demographic characteristics, El Escorial diagnostic group, cognitive measures, ALSFRS-R baseline scores, age of onset, survival, and neurological family history were all similar between neurological variant carriers vs non-carriers. All summary statistics for these measures can be viewed in Supplementary Table S2.2.

In the Answer ALS cohort, neurological variant carriers did not reach Bonferroni or nominal significance from non-carriers for any demographic, clinical, familial, or cognitive and behavioural variables. Notably, this cohort carried a distinct genetic pattern compared to Irish, including a trend towards higher frequency of *SORD* LP/P variant burden (6.4% Irish carriers vs 18.8% Answer ALS carriers, Fisher’s exact two-sided *p* = 0.0974, *Φ* = −0.18).

Detailed results for all summary statistics for both cohorts are summarised in Supplementary Table S2.2. Variant information including phenotypic associations, molecular consequence, and population frequency is provided in Supplementary Table S3.1. Case clinical information per neurological variant carrier is available in Supplementary Table S3.2.

#### Hereditary Spastic Paraplegia

Hereditary spastic paraplegia (HSP) associated variants were the most frequent in both cohorts, occurring in 15/469 (carrier frequency 3.2%) of Irish and 21/774 (carrier frequency 2.7%) of Answer ALS cases, yielding a combined carrier frequency of approximately 3.0%. Individual allele frequencies for these variants were low (10^-4^ to 10^-3^; gnomAD).

In the Irish cohort, HSP variant carriers showed similar age of onset (57.62 ± 6.65), survival (49.40 ± 21.85), site of onset (spinal 80.0%) compared to non-carriers. HSP carriers had a nominally lower proportion of Definite/Probable El Escorial classification compared to non-carriers (60.0% vs 82.5%, *p* = 0.043). Answer ALS carriers had no significant difference in age of onset (56.81 ± 4.20 years), survival (42.80 ± 19.20 months), El Escorial Category (81.0% Definite/Probable), or site of onset (spinal 66.7%) compared with non-carriers. All HSP associated genes were associated with an autosomal recessive phenotype in OMIM. Summary statistics between HSP carriers vs non-carriers are summarised in Summary Table S2.3.

Full variant-level population frequency data are provided in Supplementary Table S3.1, and detailed clinical characteristics of HSP-variant carriers are summarised in Supplementary Table S3.2

### Neurodevelopmental Variants in ALS Cohorts

ClinVar LP/P neurodevelopmental-associated variants were identified in a subset of pwALS in both cohorts (Table 3). In the Irish cohort, 12 of 469 individuals (carrier frequency 2.6%) carried LP/P neurodevelopmental variants across 11 genes including *ACP6* (*n* = 2), *PIGL* (*n* = 1), *POMT2* (*n* = 1), *SBDS* (*n* = 1), *PIGG* (*n* = 1), *AP4S1* (*n* = 1), *LSS* (*n* = 1), *FAM98C* (*n* = 1), *POMGNT1* and *TSPAN1* (*n* = 1), *AFG2A* (*n* = 1), and *POMT1* (*n* = 1). In Answer ALS, 20 of 774 individuals (carrier frequency 2.6%) carried neurodevelopmental variants across 11 genes including *GMPPB* (*n* = 3), *RNASEH2B* (*n* = 3), *PIGG* (*n* = 2), *PIGO* (*n* = 2), *EIF3F* (*n* = 2), *PIGL* (*n* = 2), *PNKP* (*n* = 2), *POMGNT1* and *TSPAN1* (*n* = 1), *SLC35A3* (*n* = 1), *PMM2* (n = 1), and *PGAP3* (*n* = 1). Heatmaps detailing the distribution of variants across associated genes, carrier frequencies, and associated neurological phenotypes are displayed in Figure 2. The allele frequencies in gnomAD for these variants ranged from 1×10^-5^ to 1.4×10^-4^ (estimated carrier frequency about 1 in 50,000 and 1 in 350). From the 19 total unique genes across both cohorts, two genes (*FAM98C* and *ACP6*) had limited information available in OMIM regarding inheritance patterns and phenotypic associations. The neurodevelopmental phenotype associations were validated for these two genes via GeneCards and SFARI Gene, respectively, and variants in these genes were retained for further analyses. The remaining 17 genes were deemed autosomal recessive in OMIM. Of note, no variants from neuropsychiatric-associated genes appeared in either cohort.

**Figure 2.**
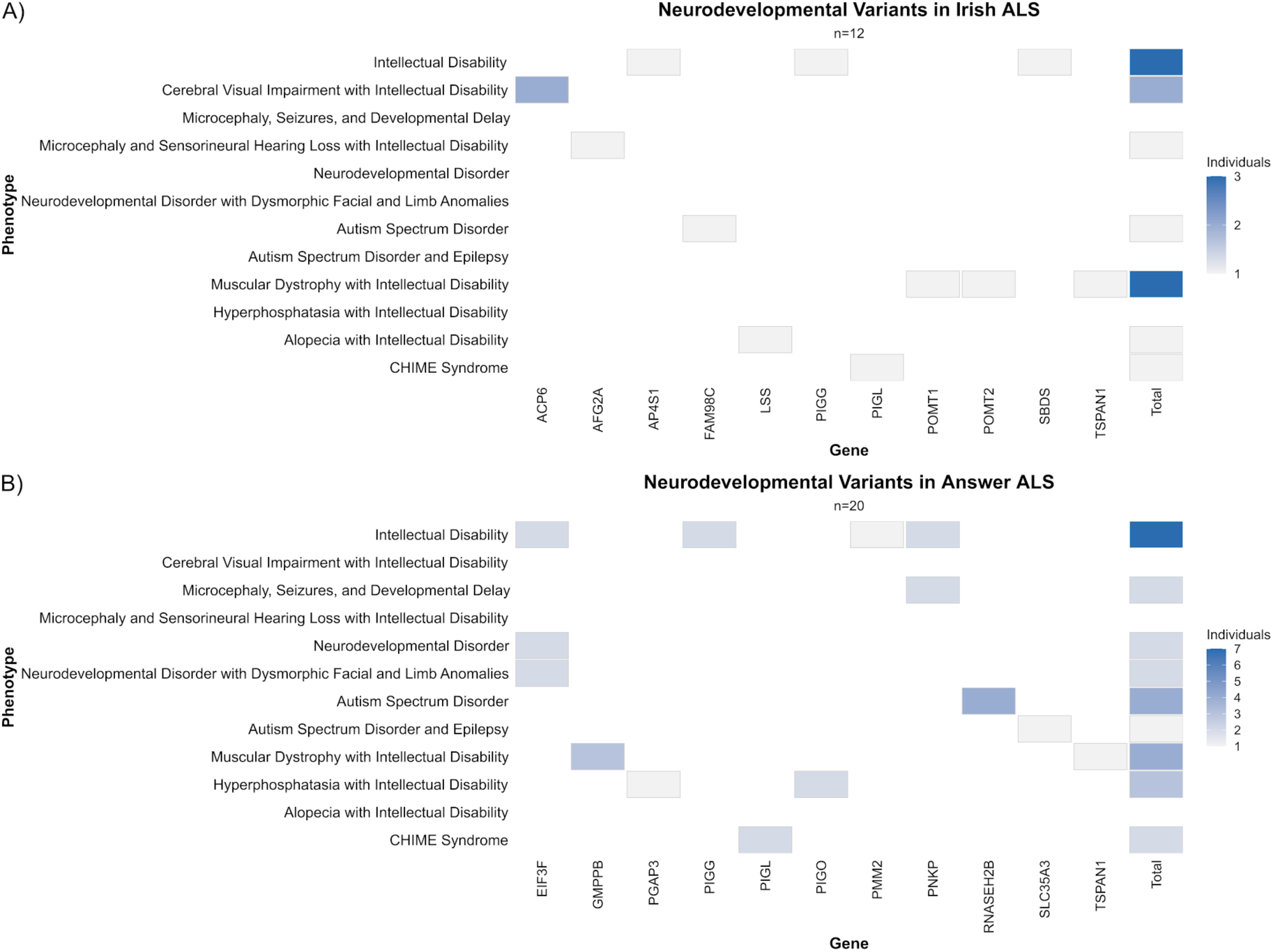
Genotype-phenotype heatmaps for LP/P neurodevelopmental variants in Irish and Answer ALS pwALS. Each row represents a neurodevelopmental phenotype associated with the annotated variants while each column represents a gene associated with each variant in A) Irish (*n* = 12) and B) Answer ALS (*n* = 20) cohorts. Variants linked to multiple phenotypes are accounted for. Each individual is counted only once per gene-phenotype cell as all individuals were heterozygous. The rightmost ‘Total’ column shows, for each phenotype, the total number of individuals with any LP/P neurodevelopmental variant across all genes.

**Table 3.**
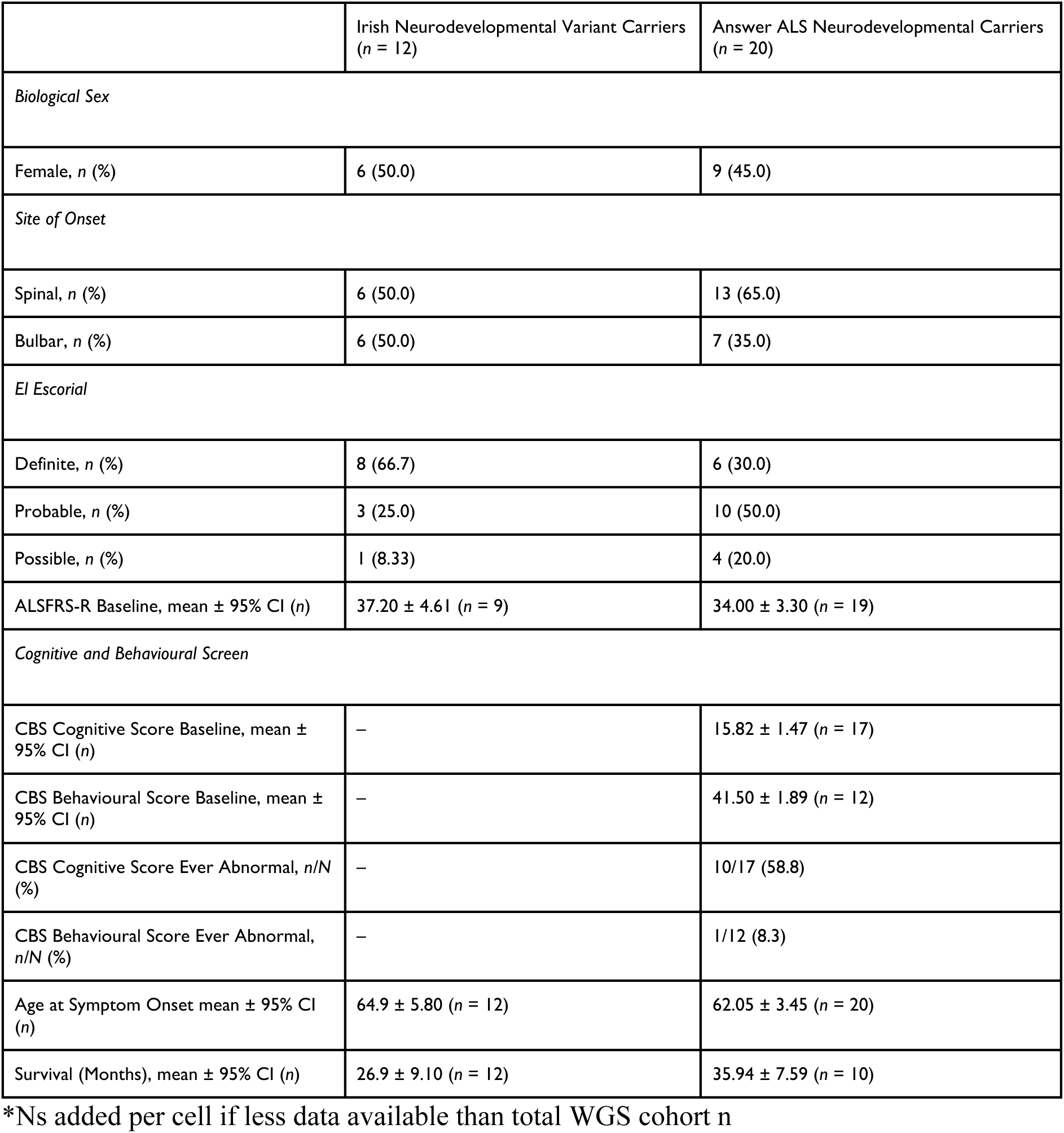
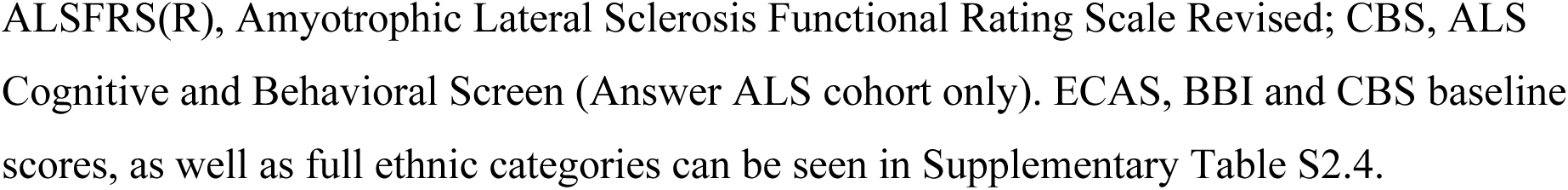
Demographic, clinical, and cognitive-behavioural characteristics of people with ALS carrying neurodevelopmental variants in the Irish and Answer ALS cohorts.

|  | Irish Neurodevelopmental Variant Carriers<br>(n = 12) | Answer ALS Neurodevelopmental Carriers<br>(n = 20) |
| --- | --- | --- |
| *Biological Sex* |  |  |
| Female, n (%) | 6 (50.0) | 9 (45.0) |
| *Site of Onset* |  |  |
| Spinal, n (%) | 6 (50.0) | 13 (65.0) |
| Bulbar, n (%) | 6 (50.0) | 7 (35.0) |
| *El Escorial* |  |  |
| Definite, n (%) | 8 (66.7) | 6 (30.0) |
| Probable, n (%) | 3 (25.0) | 10 (50.0) |
| Possible, n (%) | 1 (8.33) | 4 (20.0) |
| ALSFRS-R Baseline, mean ± 95% CI (n) | 37.20 ± 4.61 (n = 9) | 34.00 ± 3.30 (n = 19) |
| *Cognitive and Behavioural Screen* |  |  |
| CBS Cognitive Score Baseline, mean ± 95% CI (n) | – | 15.82 ± 1.47 (n = 17) |
| CBS Behavioural Score Baseline, mean ± 95% CI (n) | – | 41.50 ± 1.89 (n = 12) |
| CBS Cognitive Score Ever Abnormal, n/N (%) | – | 10/17 (58.8) |
| CBS Behavioural Score Ever Abnormal, n/N (%) | – | 1/12 (8.3) |
| Age at Symptom Onset mean ± 95% CI (n) | 64.9 ± 5.80 (n = 12) | 62.05 ± 3.45 (n = 20) |
| Survival (Months), mean ± 95% CI (n) | 26.9 ± 9.10 (n = 12) | 35.94 ± 7.59 (n = 10) |

\*Ns added per cell if less data available than total WGS cohort n

Irish neurodevelopmental variant carriers showed significantly higher neuropsychiatric family history burden in first- and second-degree relatives compared to non-carriers when accounting for total family size (Figure 3; negative binomial IRR for carriers = 2.41, 95% CI 1.12-5.18, *p* = 0.025; *n* = 11 carriers/411 non-carriers) with full family history analysis results demonstrated in Supplementary Table S2.7. Irish carriers also showed a trend towards a higher proportion of bulbar-onset disease compared with WGS non-carriers (50.0% vs 26.9%), with a corresponding reduction in spinal-onset cases (50.0% vs 70.8%; Fisher’s exact *p* = 0.062, *Φ* = 0.10). In contrast, there were no statistically significant differences between carriers and non-carriers in El Escorial diagnostic category, age at symptom onset, biological sex, baseline ALSFRS-R, ECAS scores/abnormality rate, or pre- and post-MND BBI scores/abnormality rates, with all effect sizes in these domains small to negligible.

**Figure 3.**
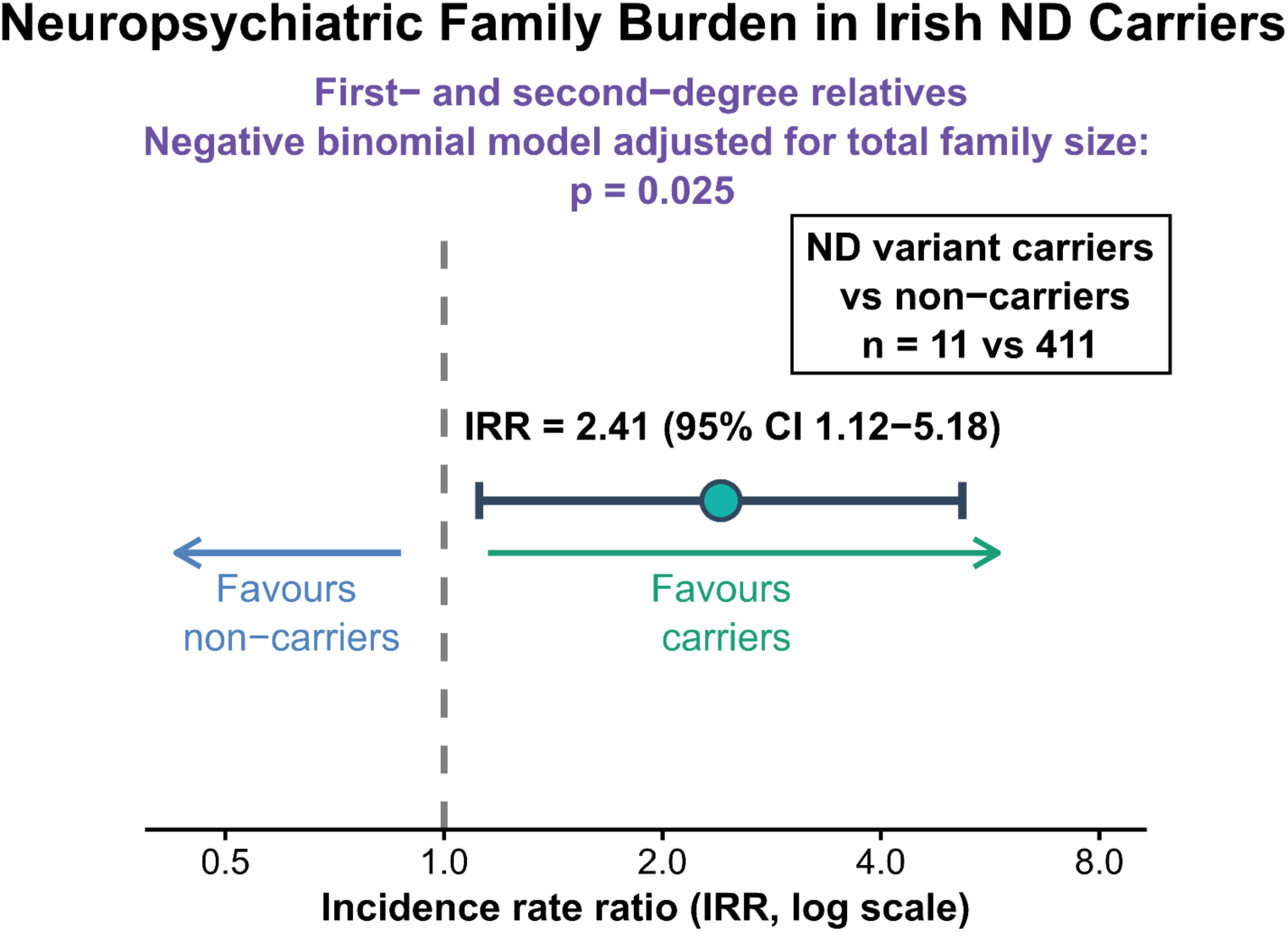
Increased neuropsychiatric family burden in Irish neurodevelopmental variant carriers. The family-size-adjusted negative binomial model supports increased neuropsychiatric burden in neurodevelopmental (ND) variant carriers (*n* = 11) compared to non-carriers (*n* = 411) (IRR = 2.41, 95% CI: 1.12-5.18, *p* = 0.025).

Kaplan-Meier survival analysis demonstrated significantly reduced survival in Irish ND variant carriers compared to non-carriers (log-rank; χ² = 9.5, *df* = 1, *n* = 12, *p* = 0.005) (Figure 4), corresponding to a more than two-fold increased hazard of death (hazard ratio = 2.25, 95% CI 1.26-4.00). This higher risk of death in ND variant carriers was independent of bulbar onset when this was included as a covariate (HR = 2.11, 95% CI 1.21-3.69, *p* = 0.009) and interaction between variant status and bulbar onset not significant (*p* = 0.960), indicating that the survival disadvantage in carriers was not driven by site of onset. No neurodevelopmental variant carriers concurrently carried established ALS-associated mutations (e.g. *C9orf72* repeat expansion).

**Figure 4.**
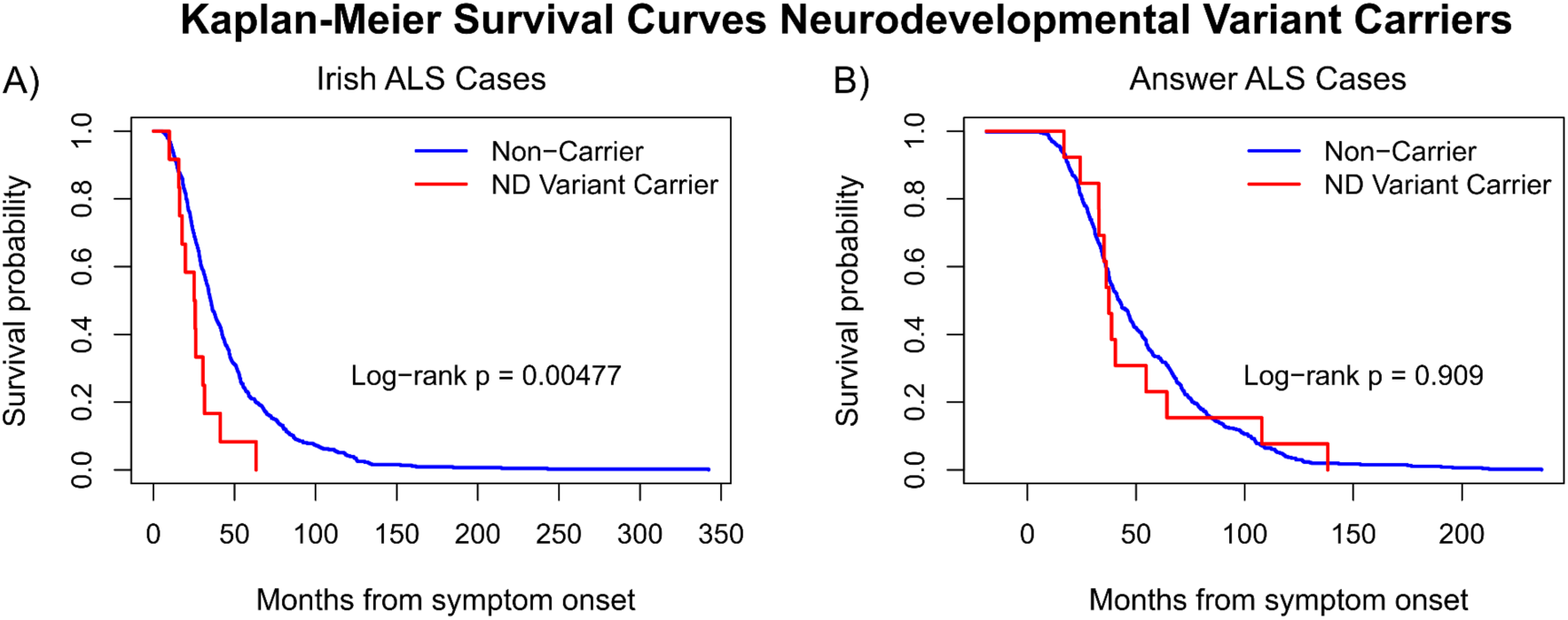
Kaplan-Meier survival curves for pwALS carrying neurodevelopmental variants. Significantly reduced survival shown in neurodevelopmental (ND) variant carriers compared to non-carriers in the A) Irish ALS cohort (carriers *n* = 12; non-carriers *n* = 433), while no survival associations were seen in B) Answer ALS cohort (carriers *n* = 13; non-carriers *n* = 463).

In Answer ALS, ND variant carriers exhibited improved baseline behavioural scores (41.50 ± 1.89 vs 37.78 ± 0.68; *p* = 0.003; Cohen’s *d* = 0.47), and a nominally lower proportion of ever meeting abnormal behaviour criteria (8.3% vs 38.9%; Fisher exact *p* = 0.035; *Φ* = 0.09), although this estimate should be interpreted cautiously due to small n with data available (*n* = 12). Baseline CBS cognitive scores were similar between carriers and non-carriers (15.82 ± 1.47 vs 16.01 ± 0.25; *p* = 0.810, Cohen’s *d* = −0.06), indicating no meaningful difference in global cognitive performance at study entry. Carriers also had nominally later age of symptom onset than non-carriers (62.05 ± 3.45 vs 56.40 ± 0.84 years; *p* = 0.005, Cohen’s *d* = 0.49). There were no meaningful differences in site of onset, El Escorial category, family history, or baseline ALSFRS-R scores. Survival analyses in Answer ALS showed no evidence for a survival effect of neurodevelopmental variant status (log-rank; χ² = 0.01; *df* = 1; *p* = 0.909; HR = 1.03; 95% CI 0.59-1.79).

Combined cohort survival analyses, as well as subgroup analyses restricted to neurodevelopmental variants versus combined neurodevelopmental and neuromuscular variants, are presented in Supplementary Results (Supplementary Fig. 1.1 and 1.2). Detailed results for all summary statistics for both cohorts are summarised in Supplementary Table S2.4. Neurodevelopmental variant information including phenotypic associations, molecular consequence, and population frequency is provided in Supplementary Table S4.1. Case clinical information per neurodevelopmental variant carrier is available in Supplementary Table S4.2.

### ALS Cases Carrying Multiple Variants in ALS and Non-ALS Associated Genes

Across both cohorts, 18 pwALS (8 Irish, 10 Answer ALS) carried at least two genetic mutations in either ALS genes and/or other neurological or neurodevelopmental associated genes and are described in Table 4. In the Irish cohort, five individuals were positive for the *C9orf72* repeat expansion and carried additional LP/P variants linked to HSP, mitochondrial neuropathy, Parkinson disease, and congenital muscular dystrophy associated with intellectual disability (*CYP7B1, POLG, PRKN, MPO, POMT1*). An additional two Irish individuals carried a separate ALS gene variant (*CHMP2B* or *ANXA11*) and a LP/P neurological variant (*PRKN* or *AP4M1*, respectively). One Irish individual carried variants in both *ACP6* (associated with intellectual disability) and *GBA1* (Parkinson disease/Lewy body dementia spectrum). In Answer ALS, six cases carried ALS-associated mutations (*C9orf72*, *TARDBP*, *SPG11*) together with neurological variants in *POLG, IGHMBP2, SPG7,* or *SORD,* including one case with three genetic hits (*C9orf72, SPG11* and *DST*). Furthermore, three individuals carried two non-ALS neurological variants in *SORD* and *PEKHG5*, *POLG* and *RFC4*, and *VPS13 and SORD* respectively, without a known ALS mutation. One Answer ALS individual carried a neurological variant in *GBA1* and a neurodevelopmental variant in *GMPPB*. Collectively, these oligogenic carriers showed diverse clinical presentations, including spinal, bulbar, and cognitive onset, frequent family histories of ALS or other neurological disease, and survival ranging from 14 to 118 months, suggesting that combinations of ALS and non-ALS variants may contribute to ALS heterogeneity. Of note, among the five cases carrying at least two variants in genes not associated with ALS, three had a significant family history of ALS despite no known ALS-causing mutation identified.

**Table 4.**
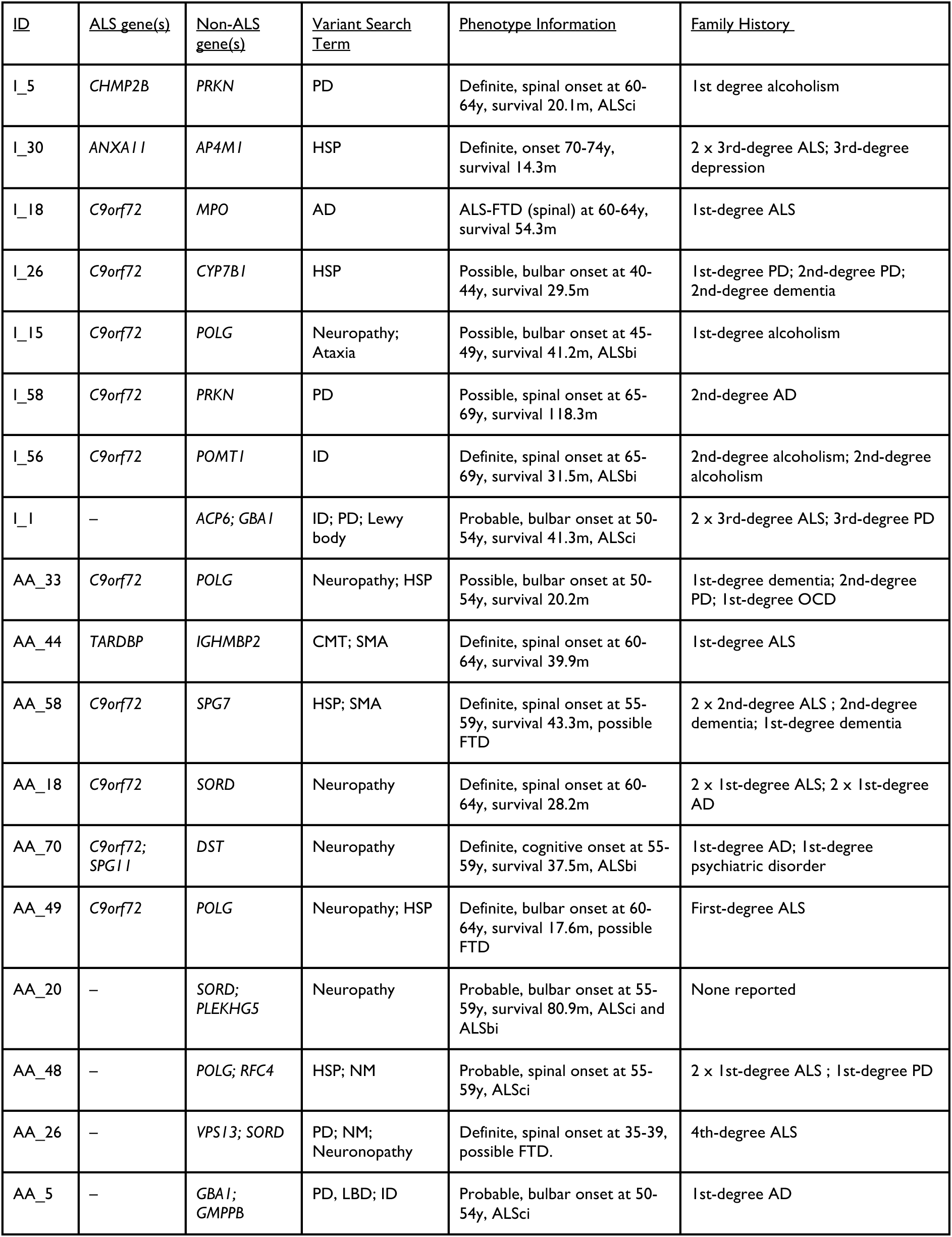
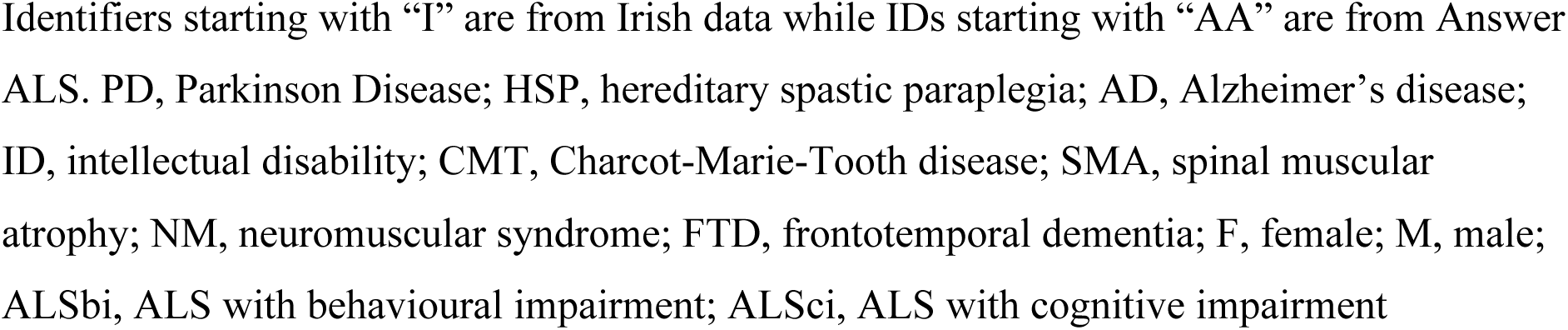
ALS cases carrying ≥2 mutations within ALS and non-ALS (neurological and neurodevelopmental) associated genes.

| ID | ALS gene(s) | Non-ALS gene(s) | Variant Search Term | Phenotype Information | Family History |
| --- | --- | --- | --- | --- | --- |
| I_5 | CHMP2B | PRKN | PD | Definite, spinal onset at 60-64y, survival 20.1m, ALSci | 1st degree alcoholism |
| I_30 | ANXA11 | AP4M1 | HSP | Definite, onset 70-74y, survival 14.3m | 2 x 3rd-degree ALS; 3rd-degree depression |
| I_18 | C9orf72 | MPO | AD | ALS-FTD (spinal) at 60-64y, survival 54.3m | 1st-degree ALS |
| I_26 | C9orf72 | CYP7B1 | HSP | Possible, bulbar onset at 40-44y, survival 29.5m | 1st-degree PD; 2nd-degree PD; 2nd-degree dementia |
| I_15 | C9orf72 | POLG | Neuropathy; Ataxia | Possible, bulbar onset at 45-49y, survival 41.2m, ALSbi | 1st-degree alcoholism |
| I_58 | C9orf72 | PRKN | PD | Possible, spinal onset at 65-69y, survival 118.3m | 2nd-degree AD |
| I_56 | C9orf72 | POMT1 | ID | Definite, spinal onset at 65-69y, survival 31.5m, ALSbi | 2nd-degree alcoholism; 2nd-degree alcoholism |
| I_1 | – | ACP6; GBA1 | ID; PD; Lewy body | Probable, bulbar onset at 50-54y, survival 41.3m, ALSci | 2 x 3rd-degree ALS; 3rd-degree PD |
| AA_33 | C9orf72 | POLG | Neuropathy; HSP | Possible, bulbar onset at 50-54y, survival 20.2m | 1st-degree dementia; 2nd-degree PD; 1st-degree OCD |
| AA_44 | TARDBP | IGHMBP2 | CMT; SMA | Definite, spinal onset at 60-64y, survival 39.9m | 1st-degree ALS |
| AA_58 | C9orf72 | SPG7 | HSP; SMA | Definite, spinal onset at 55-59y, survival 43.3m, possible FTD | 2 x 2nd-degree ALS ; 2nd-degree dementia; 1st-degree dementia |
| AA_18 | C9orf72 | SORD | Neuropathy | Definite, spinal onset at 60-64y, survival 28.2m | 2 x 1st-degree ALS; 2 x 1st-degree AD |
| AA_70 | C9orf72; SPG11 | DST | Neuropathy | Definite, cognitive onset at 55-59y, survival 37.5m, ALSbi | 1st-degree AD; 1st-degree psychiatric disorder |
| AA_49 | C9orf72 | POLG | Neuropathy; HSP | Definite, bulbar onset at 60-64y, survival 17.6m, possible FTD | First-degree ALS |
| AA_20 | – | SORD; PLEKHG5 | Neuropathy | Probable, bulbar onset at 55-59y, survival 80.9m, ALSci and ALSbi | None reported |
| AA_48 | – | POLG; RFC4 | HSP; NM | Probable, spinal onset at 55-59y, ALSci | 2 x 1st-degree ALS ; 1st-degree PD |
| AA_26 | – | VPS13; SORD | PD; NM; Neuronopathy | Definite, spinal onset at 35-39, possible FTD. | 4th-degree ALS |
| AA_5 | – | GBA1; GMPPB | PD, LBD; ID | Probable, bulbar onset at 50-54y, ALSci | 1st-degree AD |

Identifiers starting with “I” are from Irish data while IDs starting with “AA” are from Answer ALS. PD, Parkinson Disease; HSP, hereditary spastic paraplegia; AD, Alzheimer’s disease; ID, intellectual disability; CMT, Charcot-Marie-Tooth disease; SMA, spinal muscular atrophy; NM, neuromuscular syndrome; FTD, frontotemporal dementia; F, female; M, male; ALSbi, ALS with behavioural impairment; ALSci, ALS with cognitive impairment

Detailed variant annotations, ClinVar IDs and population frequencies for all oligogenic carriers are provided in Supplementary Table S3.1, S3.2, S4.1 and S4.2.

Using the observed carrier frequencies for ALS-associated mutations and non-ALS neurological and neurodevelopmental variant carriers, the expected proportion of individuals carrying both under an independence model was slightly lower than the observed proportion of oligogenic carriers (expected 6.6 vs 8 observed in the Irish cohort; expected 8.5 vs observed 10 in Answer ALS; expected 15.1 vs observed 18 across both cohorts). This suggests that, at current sample size, co-occurrence of ALS and non-ALS pathogenic variants is broadly compatible with chance.

## Discussion

This study positions rare variants in genes typically associated with other neurological and neurodevelopmental disorders as a potential additional component of ALS genetic architecture. Our findings suggest that these variants, although classically linked to other diagnoses, may influence disease presentation and mortality when they occur in ALS. In doing so, this work contributes to growing evidence supporting an expanded view of pleiotropy in ALS, in which risk and modifier alleles extend beyond ALS-related genes to a broader spectrum of neural network disorders. To our knowledge, this is the first study to systematically examine neurodevelopmental variants within an ALS cohort. Furthermore, by systematically cataloguing these pleiotropic variants across two ALS cohorts and coupling them with detailed phenotype data for each carrier, this study contributes individual-level variant-phenotype data to a literature that has largely focused on large-scale genetic studies and group-level analyses.

### Cohort Differences

Functional and demographic differences between cohorts are more likely to reflect study design and underlying population structure rather than true biological distinctions. The Irish ALS Register^33^ is population based, whereas Answer ALS^35^ uses a cohort study design. Thus, certain clinical measures may differ, for example, ALSFRS-R scores were higher in the Irish cohort, where they were recorded at first clinic visit, whereas in Answer ALS they were obtained at study enrolment, which is more likely to occur at later disease stages and therefore yields a lower ‘baseline’ score. Later age at symptom onset in Irish pwALS is consistent with underlying demographic differences, as Ireland has a slightly higher life expectancy^63^ than the USA (from which the Answer ALS cohort is drawn),^64^ and ALS is strongly age-dependent; populations with greater longevity tend to show later onset of age-related diseases.^65,66^ In addition, Answer ALS recruitment into an intensive longitudinal research programme was not designed as a population-based study and may have favoured relatively younger and more motivated participants, introducing a subtle age bias. The higher prevalence of reported family history of ALS, FTD, neurological, and psychiatric disease in the Irish cohort is plausibly influenced by differences in ascertainment. The Irish population-based surveillance uses a detailed, repeated family history questionnaire and, when possible, confirms reports with close relatives or spouses, whereas family history in Answer ALS is captured via a more limited, self-reported framework. Disparities in data collection, combined with the biological and clinical heterogeneity already associated with ALS, contribute to challenges in data analysis and interpretation, as previously described in the literature.^67,68^

### Phenotypic Associations of Neurological Variant Carriers

The carrier frequency of neurological likely-pathogenic/pathogenic (LP/P) variants was similar across cohorts (∼9.5% carrier rate; Figure 1, Table 2), which is consistent with emerging evidence that variants in other neurological genes are not uncommon in ALS populations.^11,16^ Genotype-phenotype correlations were observed only in the Irish cohort, where carriers had a significantly higher proportion of respiratory-onset disease and less severe behavioural changes pre- and post-ALS diagnosis. Although no such clinical associations were seen in Answer ALS carriers, these comparisons are limited by differences in cohort data collection. For example, premorbid behavioural change, which is measured by the BBI in Ireland, is not measured by the ALS-CBS used in Answer ALS. While the higher proportion of respiratory-onset among Irish neurological variant carriers may suggest differences in motor-system involvement, this finding is based on small numbers (5/47) and will require replication. Overall, these findings are in keeping with current evidence indicating that neurological variants outside of the typical ALS genetic spectrum may affect the phenotypic expression of ALS, potentially through shared pathomechanisms in genes such as *POLG,*^11,69^ *PRKN,*^11^ *SORD,*^70^ and *SPG7,*^48,71^ which have been implicated in neurodegenerative disease spectra and, for *SPG7*, proposed as an ALS risk or modifying factor.^48^ However, the lack of replicated clinical findings in Answer ALS likely derives from limited statistical power and cohort heterogeneity, as well as population-specific genetic architectures. Notably, *SORD* variants occurred nearly three times more frequently in Answer ALS compared to the Irish cohort. Taken together, these findings indicate that other neurological variants may have a cohort-specific impact on ALS presentation, and that careful harmonisation of clinical data is necessary when comparing modifying signals across studies.

HSP was the most frequent variant-to-phenotype relationship in both cohorts and can notably mimic ALS clinically and pathologically. However, HSP-variant carriers in both cohorts predominantly exhibited a phenotype more consistent with ALS than HSP, including symptom onset in the late 50s, contrasting with the more typical adult-onset HSP peak in the second to fourth decades of life.^72^ Furthermore, survival (symptom onset to death) for these individuals averaged about 49 and 42 months in the Irish and Answer ALS cohorts, respectively, which differs from the decades-long progression described in HSP.^72^ Most HSP-variant carriers met definite or probable El Escorial diagnostic criteria for ALS, suggesting that these variants act within the ALS spectrum rather than producing a typical HSP presentation. No HSP-associated variants were in genes with an autosomal dominant inheritance pattern, further supporting a modifying effect rather than misdiagnosis. Despite the rarity of the HSP-variants identified in the current annotation (allele frequency <0.1%, prevalence 1-6/100,000 people),^72^ carriers accounted for approximately 3% of patients in both ALS cohorts (Supplementary Table S3.1), raising the possibility of enrichment and aligning with current hypotheses that HSP-associated variants may contribute to ALS risk.^48,71,73^ Although SKAT-O variant-burden analyses showed no enrichment between cases and controls, these analyses were underpowered relative to the thousands of individuals typically required for variant-burden studies,^74^ particularly given the limited number of Answer ALS controls (*n* = 94). At the gene level, signals spanned *SPAST, SPG11, POLG*, AP-complex genes (*AP4M1, AP4B1, AP5Z1*), *SPG7, PNPLA6, DDHD1, FA2H, FARS2*, and *MTRFR*. This pattern mirrors recent large-scale ALS sequencing efforts, including Project MinE^34^ and related multicentre cohorts, which have reported enrichment of variants in HSP-associated genes such as *SPG7*, *SPG11* and *AP*-complex among ALS cases.^73^ Notably, biallelic *SPG11* mutations are an established cause of autosomal recessive juvenile-onset ALS^75^ and of classical HSP with thin corpus callosum,^76^ underscoring *SPG11’s* phenotypic continuum in which modifier context may drive ALS- versus HSP-like presentation.^71^ To determine true enrichment of HSP variants within ALS populations and their potential clinical or modifying effects, larger, ancestry-matched case-control datasets and harmonised HSP gene sets will be required.

### Phenotypic and Family History Associations in Neurodevelopmental Variant Carriers

Neurodevelopmental variants emerged as an interesting pleiotropic set in this study that has not, to our knowledge, been considered in prior ALS cohorts. Carriers of neurodevelopmental variants were relatively rare but present in both cohorts, with the affected genes implicated in brain development and broader neural network vulnerability across a range of neurodevelopmental and psychiatric disorders.^77–80^ Emerging evidence indicates that neurodevelopmental genes can underlie adult-onset neurological disease, including spastic paraplegias, dystonias, ataxias, and other progressive movement disorders,^81,82^ highlighting pleiotropy across the lifespan. In this context, investigating the role of such variants in ALS phenotype and outcome may provide insight into how neurodevelopmental risk profiles interact with adult-onset motor neuron pathology.

In the Irish cohort, those carrying neurodevelopmental variants had significantly shorter survival compared to non-carriers, corresponding to more than doubling of hazard of death and a clear separation of Kaplan-Meier curves. This association remained statistically significant after adjustment for site of onset, implying that the survival disadvantage was independent of the tendency towards bulbar-onset disease in carriers. While a survival disadvantage was not observed in Answer ALS, the combined, cohort-stratified analysis continued to show a trend toward poorer survival in carriers (HR = 1.4, *p* = 0.08). Proportional hazards diagnostics showed that the relative risk due to carrier status remained stable across the disease trajectory. The attenuation of the survival association in combined cohorts, in conjunction with a significant interaction effect between variant status and cohort, suggests that background genetic architecture, environmental exposures, or population-specific ALS risk variant spectra may influence the survival impact of these alleles. In the Irish cohort, carriers also exhibited higher familial burden of neurodevelopmental and neuropsychiatric traits, which suggests these individuals carry an increased neurodevelopmental and neuropsychiatric genetic load. One possible mechanism is that neurodevelopmental-associated mutations may influence the biological resilience of neural networks, such that carriers may have reduced capacity to compensate when a neurodegenerative process occurs, contributing to poorer survival. This aligns with broader evidence that neurodevelopmental risk genes can affect synaptic, epigenetic, and mitochondrial pathways,^83^ possibly affecting compensatory processes during ageing. To answer whether neurodevelopmental variants truly affect survival in ALS, cohorts with larger sample size required for adequate power in rare-variant burden and modifying analyses, as well as comprehensive clinical data, and systematic variant annotation, will be needed in future studies.

### Expanded Oligogenic Cases

Approximately 1-6% of pwALS have multiple hits in ALS-associated genes, otherwise known as oligogenic events.^6,7^ An oligogenic expanded pattern, which extended to LP/P neurological and/or neurodevelopmental variants as well as known ALS-associated mutations, occurred rarely but was biologically interesting in these cohorts. Among the two datasets, 18 (1.45%) of pwALS carried at least two genetic hits in ALS genes and/or other neurological or neurodevelopmental genes. For example, several individuals positive for the *C9orf72* repeat expansion also carried other LP/P variants associated with HSP, mitochondrial, parkinsonian, neuromuscular, or neurodevelopmental phenotypes. These expanded oligogenic carriers displayed considerable clinical variability in terms of site of onset, behavioural and cognitive involvement, and survival ranging from 12 months to over 10 years. In addition, these individuals often reported a positive family history of either ALS, parkinsonism, dementia, or other neurological diseases, even in cases lacking any ALS-causal mutation. These observations support a view of ALS as part of a broader, complex, overlapping genetic spectrum in which multiple rare variants may contribute to phenotypic heterogeneity.

However, in comparison with expected counts for independence based on the frequency of ALS, neurological, and neurodevelopmental variant carriers in both cohorts, the proportion of oligogenic carriers was slightly elevated but statistically insignificant (15.1 expected versus 18 observed). Thus, while not statistically significant, our observations could point towards an expanded oligogenic landscape of ALS, where combinations of ALS and non-ALS variants may be over-represented in ALS cohorts. Given that a substantial proportion of ALS cases remain genetically unexplained,^84^ identification of causative variants may be incomplete and thus reduce the degree of oligogenic enrichment we see. Nevertheless, the presence of clinically relevant variant combinations such as *C9orf72* with *SPG11, SPG7, POLG, PRKN, GBA1,* or *SORD,* all of which have independent links to motor neuron disease, parkinsonism, neuropathy, or HSP, aligns with broader evidence that multiple rare variants may interact to shape neurodegenerative phenotypes.^11,16,73^ Further research involving large numbers of sequenced cases coupled with formal oligogenic and epistatic modelling and segregation analysis will be required to explore this hypothesis on expanded oligogenic carriers in ALS.

### Limitations

There are several important limitations to note in our study. Firstly, the case-control sample used for rare variant-burden analysis (1,243 cases, 324 controls) has limited power to detect rare allele enrichment, since such studies often require a sample size of thousands of individuals to produce statistically meaningful results.^74^ Consequently, the negative SKAT-O finding does not rule out the possibility of any enrichment effects. More powerful cohorts including larger WGS control samples are required to test for enrichment of other neurological and neurodevelopmental variants in ALS.

Secondly, despite considering LP/P variants from ClinVar (which requires expert review), the possibility of misclassification remains an issue with variants of uncertain significance, incomplete penetrance, and variable expressivity, particularly in pleiotropic genes - meaning this classification might both miss true pathogenic variants and include false positives. Thirdly, because both cohorts consist predominantly of individuals of European descent, the role and range of pleiotropic variants among pwALS of other ancestries cannot be inferred from the current data. Additionally, caution is warranted while interpreting the results as cohorts exhibit differences in study design, which may have influenced lack of replication of clinical findings between carriers. Lastly, although the survival analysis was adjusted for multiple covariates, the possibility of residual confounding by factors such as age, comorbid conditions, and exposure to treatment cannot be excluded. Future research with larger, ancestrally diverse samples, rich covariate data, and formal rare-variant burden, modifier, and oligogenic modelling will be required to quantify how these pleiotropic variants contribute to ALS risk and outcome.

### Conclusions and Future Directions

The current study demonstrates that a measurable proportion of pwALS from two independent cohorts carry likely-pathogenic/pathogenic variants in genes that are not classically associated with ALS. Specifically, 9.5% of individuals carried variants for other neurological traits, 3% had HSP-associated variants, and 2.6% carried neurodevelopmental variants. Our approach to integrate structured variant cataloguing with detailed phenotypic data yields one of the first systematic databases linking variants in non-ALS genes to ALS subphenotypes, which can be used in future meta-analyses and rare-variant burden studies.

These findings contribute to a broader understanding of genetic pleiotropy in ALS, in which genes traditionally associated with other neural network disorders might act as risk alleles or modifiers in ALS. Patterns of clinical presentation of other neurological variant carriers, reduced survival among carriers of neurodevelopmental variants, and instances of expanded oligogenic models with combinations of ALS and non-ALS variants suggest that a multi-gene architecture is a potential determinant of heterogeneity in ALS and warrants further investigation. The incorporation of such pleiotropic variants in future studies with larger samples and harmonised data may enhance genetic risk stratification and identify biologically relevant subgroups which may give insight to prognosis, family counselling, and development of targeted therapies.

## Acknowledgements

We would like to thank Kimberly Kiyui for her contribution to neuropsychological testing in the Irish ALS cohort. Data used in the preparation of this article were obtained from the Answer ALS Data Portal (AALS-01184). For up-to-date information on the study, visit https://dataportal.answerals.org. We also thank REDcap ^33^ which supports the Irish ALS Register and PRECISION ALS.^85^

## Supplementary material

Supplementary material is available at *Brain* online.

