## Supplementary material for "Rare neurological and neurodevelopmental variants in ALS link to onset, survival and family history": S1

- Supplementary Methods
    - o SKAT-O Variant-Set Enrichment Analysis
    - o Literature Review
- Supplementary Results
    - o Neurodevelopmental survival Analysis
        - o Distribution of molecular consequences in neurologic versus neurodevelopmental variants
        - Molecular consequence


### Supplementary Methods

### SKAT-O Analysis

Variant-set enrichment analysis was evaluated using the Sequence Kernel Association Test (SKAT/SKAT-O) in R (version 4.1.2). Three prespecified variant sets were considered based on our literature search: non-ALS neurologic variants, combined neurodevelopmental and neuropsychiatric variants, and hereditary spastic paraplegia (HSP) variant sets. Each set comprised all unique ClinVar variant IDs from each phenotypic category.

Variant genomic coordinates were harmonised and linked to non-Finnish European gnomAD allele frequencies when available as majority of Irish and AnswerALS cohorts were White, Non-Hispanic. Three nested allele frequency thresholds were set (AF NFE $\leq$0.01, $\leq$0.001, and $\leq$0.0001). For each set, we curated a binary genotype matrix across all individuals (per-variant carrier/non-carrier) and modelled ALS case control status as the outcome. SKAT-O was run with the weighted linear kernel and default Beta(1,25) minor allele frequency weights. The only covariate included in the null model was cohort (Irish vs AnswerALS, cases and controls) to account for potential between-cohort differences. As a robustness check for the binary trait, we also applied the SKATBinary_Robust procedure with the same kernel and covariate structure. For each variant set, we report SKAT-O p-values together with the proportion of ALS cases and controls carrying at least one variant in the set in Supplemental Table S2.5.

### Phenotype Literature Review

**Supplementary Table S1.** Full list of phenotypes used as ClinVar search terms, including neurological, neuropsychiatric and neurodevelopmental traits, and rationale for inclusion. Search terms bolded appeared in variant annotation.

| Phenotype (Category) | Evidence Type, Rationale, and Key Refs. |
|---|---|
| *Neurological* | |
| **Alzheimer's disease (neurodegenerative)** | Family aggregation studies showing excess Alzheimer's disease in ALS kindreds versus controls; cross-trait GWAS demonstrating shared common variant architecture between ALS and Alzheimer's disease.[1–3] |
| **Cerebellar Ataxia (neurological)** | Familial co-aggregation and meta-analysis of ATXN2 CAG repeats as shared ALS/SCA2 risk; broader pleiotropy via pathogenic variants in >30 ataxia genes (e.g., *CACNA1A*, *SPG7*) recurrent in ALS cohorts.[4–6] |
| **Charcot-Marie-Tooth disease (neuromuscular)** | Co-occurrence case reports of genetically confirmed CMT1A and CMT2A (MFN2) with superimposed clinically definite ALS; literature/database synthesis demonstrating pleiotropic CMT-ALS genes (e.g., MFN2, NEFH, KIF5A, VCP, FIG4) and shared axonal/mitochondrial pathways supporting true clinicogenetic overlap.[7–10] |
| Cognitive Impairment (neurological) | Systematic review/meta-analysis and population based cohort studies showing that a substantial proportion of ALS patients exhibit cognitive and//or behavioural impairment, including ALS FTD, indicating that frontotemporal network vulnerability is intrinsic to ALS and justifying inclusion of dementia/FTD phenotypes.[11,12] |
| **Hereditary spastic paraplegia (neuromuscular)** | SPG7 rare-variant enrichment in ALS cohorts with HSP-like clinical features; KIF5A, ALS2, ERLIN2, and SPG11 mutations reported across HSP and ALS pedigrees, and genotype phenotype studies showing domain-specific KIF5A variants causing either HSP/CMT2 or ALS, together supporting a motor-neuron continuum and justifying inclusion of HSP variants.[13–16] |

| | |
|---|---|
| **Lewy body dementia (neurodegenerative)** | Neuropathology study showing clinically relevant Lewy body disease in a subset of ALS brain-bank cases at a prevalence higher than the general population; cross-trait GWAS demonstrating shared loci and polygenic overlap between ALS, Lewy body dementia, and related dementias.[3,17] |
| Multiple sclerosis (neurological) | Clinicopathological and literature based series describing reported co-occurrences of MS and ALS, including pathologically confirmed dual MS ALS with TDP-43 inclusions and FUS mutated cases, suggest that although rare, concurrent MS-ALS may reflect shared neuroinflammatory and neurodegenerative mechanisms.[18–20] |
| Multiple system atrophy (neurodegenerative) | C9orf72 hexanucleotide repeat expansion families in which different members develop either ALS or clinically/radiologically defined MSA, and pathologically confirmed case of combined ALS-C9orf72 and MSA with coexisting TDP-43 and alpha-synuclein pathology, demonstrating that motor neuron degeneration and MSA-like synucleinopathy can arise from shared genetic background; although current evidence is largely C9orf72-driven MSA is included as a hypothesis-generating phenotype to screen for rare, non-repeat pathogenic variants in overlapping neurodegenerative pathways that might modify ALS risk or phenotype.[21,22] |
| **Parkinson disease (neurodegenerative)** | Population-based family aggregation and register-based case-control studies demonstrating excess Parkinson's disease in ALS kindreds versus controls, together with cross-trait GWAS evidence for shared polygenic risk between ALS and parkinsonian disorders, supporting pleiotropic variants affecting nigrostriatal and corticospinal networks.[1–3] |
| Progressive supranuclear palsy (neurodegenerative) | Joint GWAS reveals genetic correlation between PSP and ALS, plus a clinicopathological PSP ALS co-morbidity case, suggesting shared susceptibility loci and overlapping tau/TDP-43 related mechanisms across these syndromes.[23,24] |
| **Peripheral neuropathy/axonal hereditary neuropathy (neuromuscular)** | Systematic review and observational NGS cohort showing that sensory neuropathy is present in a sizable minority of ALS patients and that rare variants in axonal hereditary neuropathy genes may influence ALS survival, supporting interrogation of hereditary neuropathy loci as potential ALS modifiers.[25,26] |

| | |
|---|---|
| Other **neuromuscular/neurodegenerative disorders (neuromuscular)** | Population-based cross-sectional genetic analysis demonstrating that people with ALS have higher rates of relatives with other neurodegenerative disorders and that about 10% carry rare variants in genes classically associated with non-ALS neuromuscular/neurodegenerative diseases; underscoring pleiotropy as a major feature of ALS genetics.[27] |
| Inclusion body myopathy/multisystem proteinopathy (neuromuscular) | Case series and narrative review showing that VCP and related genes cause multisystem proteinopathy with combined inclusion body myopathy, Paget disease, FTD and motor neuron involvement, expanding the ALS spectrum to include inclusion-body myopathy phenotypes driven by shared protein-homeostasis pathways.[28,29] |
| *Neuropsychiatric Disorders* | |
| Neuropsychiatric disorders: anxiety, depression, bipolar disorder, psychosis, schizophrenia | Population-based family aggregation, case control and register-based studies showing higher rates of mood, anxiety, psychotic and schizophrenia-spectrum disorders in ALS kindreds relative to controls, plus C9orf72- positive families with bipolar disorder, supporting shared liability to neuropsychiatric phenotypes within ALS pedigrees and pleiotropic ALS psychiatric risk variants.[1,2,30,31] [1,31] |
| Alcohol use disorder (neuropsychiatric/addiction) | Population-based family aggregation and register-based studies exploring alcohol dependency in ALS kindreds relative to controls.[1,31] |
| *Neurodevelopmental Disorders* | |
| **ADHD and autism spectrum disorder (neurodevelopmental)** | Population-based family aggregation and family based cross-sectional work showing enrichment of ADHD and autism diagnoses/traits in ALS families and in relatives of ALS patients with cognitive and neuropsychiatric endophenotypes, supporting shared neurodevelopmental liability.[31,32] |
| Speech and language delay (neurodevelopmental) | Cross-sectional observational and family-based studies indicating that presymptomatic C9orf72 carriers can show developmental verbal fluency and language deficits, and that language endophenotypes segregate in ALS families, justifying inclusion of speech/language developmental phenotypes.[32,33] |

| | |
|---|---|
| **Intellectual disability** | Case series and reports of juvenile ALS due to FUS or C9orf72 mutations presenting with intellectual disability and psychiatric disease, together with family-based endophenotype data linking intellectual impairment to ALS-associated variants, supporting intellectual disability as part of the broader ALS pleiotropic spectrum.[32,34–36] |

SCA, Spinocerebellar Ataxia Type 2; NGS, Next Generation Sequencing; ADHD, Attention-Deficit Hyperactivity Disorder

### Supplementary Results

#### Neurodevelopmental Survival Analyses

In a combined analysis including 921 deaths across both cohorts, a cohort-stratified Cox model demonstrated a trend towards reduced survival in carriers of neurodevelopmental variants that did not reach conventional statistical significance (stratified Cox model: HR 1.44, 95% CI 0.97-2.13, p=0.07; log-rank p=0.082), with proportional hazards supported for variant status (Schoenfeld p=0.37). A model including a variant-by-cohort interaction indicated that the hazard associated with variant status was close to null in AnswerALS (HR ~1.0) but higher in the Irish cohort (interaction HR 2.48, 95% CI 1.13-5.42, p=0.02), suggesting between-cohort heterogeneity in the survival impact of these variants.

In a cohort stratified Cox model including three ALS groups (neurodevelopmental only (ND) (n = 15; n = 8 Irish and n = 7 AnswerALS), neurodevelopmental and neuromuscular (ND+NM) (n = 12; n = 5 Irish; n = 6 AnswerALS), and non-carriers (n = 895; n = 432 Irish; n = 463 Answer ALS), hazard ratios were 1.28 (95% CI 0.77-2.14, p = 0.35) for ND-only carriers and 1.62 (95% CI 0.91-2.87, p = 0.10) for ND + NM carriers compared with non-carriers, with a non-significant overall test (Wald p = 0.20; log rank p = 0.20). The three group Kaplan Meier curves similarly suggested numerically shorter survival in carrier groups, particularly NM+ND but without significant difference (global log rank p = 0.15). Direct comparison of ND+NM versus ND only carriers, both in the full sample (HR 1.27, 95% CI 0.59-2.72, p = 0.54) and when restricting to carriers only (HR 0.76, 95% CI 0.31-1.85, p = 0.54; log-rank p = 0.75), did not demonstrate any statistically reliable survival difference between these two variant categories. Proportional hazards testing for the three sample group model indicated some deviation from proportionality for the grouped term (global Schoenfeld test p = 0.035), so these subgroups estimates should be interpreted cautiously.

#### Kaplan-Meier Survival Curves Neurodevelopmental Variant Carriers

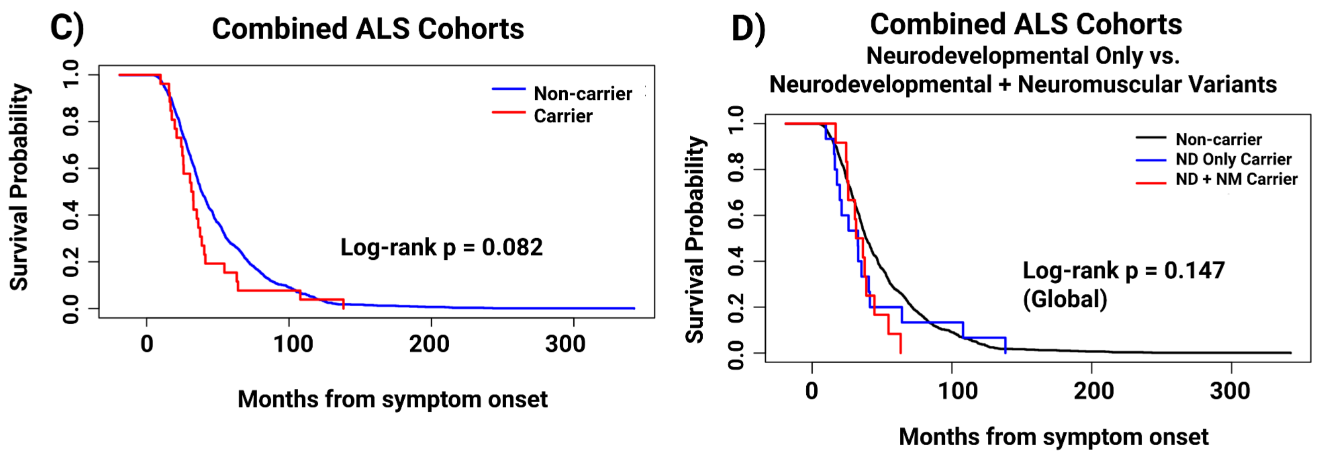


**Supplemental Figure S1.1 Kaplan-Meier survival curves comparing pwALS with neurodevelopmental variants versus non-carriers.** Kaplan Meier survival curves comparing pwALS with and without rare neurodevelopmental variants. C) Combined Irish and AnswerALS cohorts, showing a trend towards shorter survival in carriers that do not reach conventional statistical significance. D) Combined cohort analysis contrasting three groups-non-carriers, neurodevelopmental-only (ND) carriers, and carriers of variants associated with both neurodevelopmental and neuromuscular (ND+NM) variants with no statistically significant differences in survival between groups.

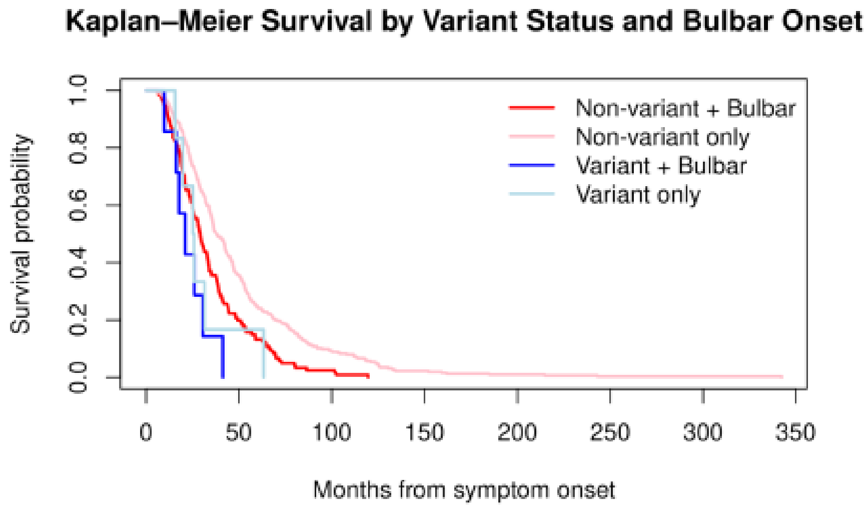

**Supplemental Figure S1.2 Kaplan-Meier survival curves for Irish ALS patients by neurodevelopmental variant status and bulbar onset.** Survival from symptom onset to death (months) is shown for four groups: non-carriers with bulbar onset (red), non-carriers without bulbar onset (pink), ND variant carriers with bulbar onset (dark blue), ND variant carriers without bulbar onset (light blue).

#### Distribution of molecular consequences in neurologic versus neurodevelopmental variants

The distribution of molecular consequences differed subtly between neurological and neurodevelopmental variants (Supplemental Figure 1.3). Across both cohorts combined, neurologic variants were most frequently missense (n=20), followed by nonsense and frameshift changes, with smaller numbers of splice and non-coding transcript variants. Neurodevelopmental variants also showed a predominance of missense consequences (n=10) but relatively fewer nonsense and frameshift changes and no non-coding transcript variants, indicating that both variant sets are enriched for protein-altering changes with comparable molecular consequence profiles in this dataset.

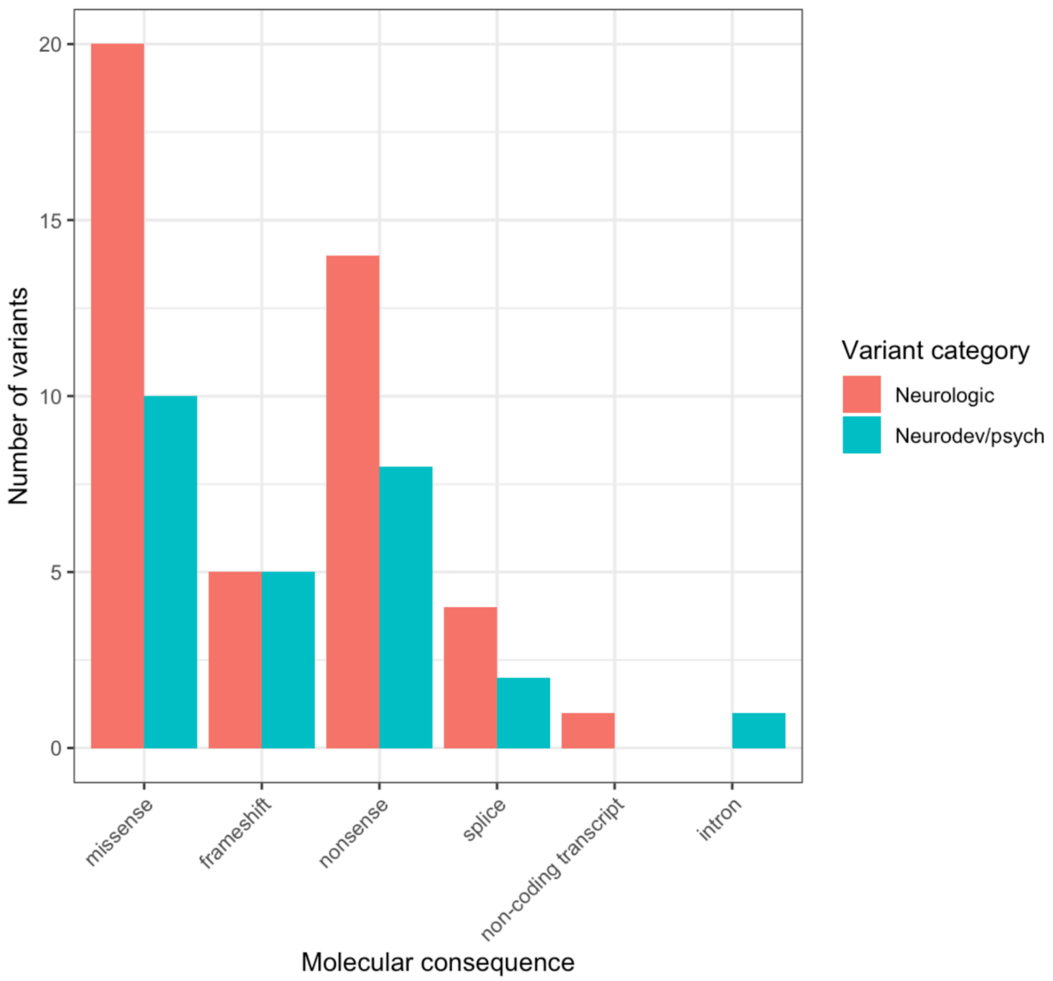


**Supplemental Figure S1.3 Molecular consequences of ClinVar LP/P neurological and neurodevelopmental variants identified in combined ALS cohorts.** Bar plot showing the number of ClinVar LP/P variants by molecular consequence (missense, frameshift, nonsense, splice, non-coding transcript, intronic) for variants that appeared from annotation associated with neurological phenotypes (red) and neurodevelopmental phenotypes (blue) across both ALS cohorts combined. Missense variants predominated in both categories, with a higher absolute burden of missense and nonsense consequences among neurological variants, whereas neurodevelopmental variants showed relatively fewer frameshift events and no non-coding transcript variants in this dataset.

12. Montuschi A, Iazzolino B, Calvo A, et al. Cognitive correlates in amyotrophic lateral sclerosis: a population-based study in Italy. *J Neurol Neurosurg Psychiatry*.

2015;86(2):168-173. doi:10.1136/jnnp-2013-307223
