## Supplementary material for "Rare neurological and neurodevelopmental variants in ALS link to onset, survival and family history": S2

***Supplemental Table S2.1*. Demographic and clinical characteristics with statistical comparisons of pwALS from Irish and AnswerALS cohorts.** Values are shown as percentages (*n*) for categorical variables and mean ± 95% confidence interval (*n*) for continuous variables. For categorical variables, the *n* in parentheses is the count corresponding to the percentage in that cell, not the total cohort size. For continuous variables, the *n* reported is the number of individuals with available data for that measure. Cohort differences between categorical variables were evaluated using $\chi^2$ tests (without Yates' correction if all expected cell counts ≥ 10; with Yates' correction if any expected cell count 5-9), and Fisher's exact tests when any expected cell count <5. Continuous variables were assessed via Welch's two sample t-tests. For categorical variables, effect size is given as Φ for 2x2 tables and Cramér's V for tables with more than two categories. For Fisher's exact test, Φ was calculated from the Pearson chi-squared statistic. For continuous variables, effect size is reported as Cohen's *d*. Bonferoni-corrected significance threshold of $p < 0.0042$ for 12 independent tests was applied. P-values that are < 0.05 but ≥ 0.0042 are considered nominally significant and are described in the main text.

| ***Variable*** | ***Category*** | ***Irish: Count (%) or mean ± 95% CI (n)*** | ***AnswerALS: Count (%) or mean ± CI (n)*** | ***Variable Type*** | ***Statistical Test*** | ***Test Statistic*** | ***df*** | ***p-value*** | ***Effect Size*** |
|---|---|---|---|---|---|---|---|---|---|
| Sex | Female | 40.0 (n=182) | 37.1 (n=287) | Categorical | Chi-Squared | $\chi^2 \sim 1.0$ | 1 | 0.309 | Φ=0.03 |
| Site of Onset | Spinal | 70.1 (n=319) | 68.0 (n=526) | Categorical | Chi-Squared | $\chi^2 \sim 0.6$ | 1 | 0.432 | Φ=0.02 |
| Site of Onset | Bulbar | 27.7 (n=126) | 21.7 (n=168) | Categorical | Chi-Squared | $\chi^2 \sim 5.3$ | 1 | 0.021 | Φ=0.07 |
| El Escorial | Definite | 54.4 (n=227) | 30.0 (n=232) | Categorical | Chi-Squared | $\chi^2 \sim$ *80.5* | *3* | *2.20e-16* | Cramér's V=0.15 |
| El Escorial | Probable | 26.9 (n=112) | 50.3 (n=389) | Categorical | – | – | – | – | – |
| El Escorial | Possible | 17.3 (n=72) | 15.6 (n=121) | Categorical | – | – | – | – | – |
| El Escorial | Suspected | 1.4 (n=6) | 3.1 (n=24) | Categorical | – | – | – | – | – |
| Family History | ALS | 20.7 (n=94) | 13.3 (n=92) | Categorical | Chi-Squared | $\chi^2 \sim 10.3$ | 1 | 0.001 | Φ=0.09 |

| | | | | | | | | | |
|---|---|---|---|---|---|---|---|---|---|
| Family History | FTD | 3.5 (n=16) | 0.6 (n=4) | Categorical | Fisher | NA | NA | 0.0003 | Φ=0.11 |
| Family History | Neurological | 51.4 (n=234) | 42.0 (n=290) | Categorical | Chi-Squared | $\chi^2 \sim 9.4$ | 1 | 0.002 | Φ=0.09 |
| Family History | Psychiatric | 54.9 (n=250) | 10.7 (n=74) | Categorical | Chi-Squared | $\chi^2 \sim 262.1$ | 1 | <2.20e-16 | Φ=0.48 |
| Age of Symptom Onset (Years) | – | 62.96 ± 1.02 (±1.63%) (n=454) | 56.54 ± 0.82 (±1.46%) (n=758) | Continuous | T-Test (Welch) | $t \sim 9.6$ | ~1180 | *4.26e-18* | Cohen's *d*=0.57 |
| Survival (Symptom Onset to Death)(Months) | – | 44.77 ± 3.27 (±7.31%) (n=445) | 43.4593 ±3.14 (±7.22%) (n=353) | Continuous | T-Test (Welch) | $t \sim 0.6$ | ~780 | 0.571 | Cohen's *d*=0.04 |
| ALSFRS-R Baseline | – | 36.72 ± 0.71 (±1.95%) (n=375) | 33.97 ± 0.61 (±1.78%) (n=750) | Continuous | T-Test (Welch) | $t \sim 5.8$ | ~ 835 | 1.17e-08 | Cohen's *d*=0.35 |
| Ethnic Category | White - Not Hispanic or Latino | 100.0 (469) | 88.1 (n=682) | Categorical | Fisher | NA | NA | <2.20e-16 | Φ=0.22 |
| *C9orf72* Repeat Expansion Status | Positive | 10.1 (n=46) | 6.0 (n=46) | Categorical | Chi-Squared | $\chi^2 \sim 6.4$ | 1 | 0.012 | Φ=0.07 |

**Supplemental Table S2.2. Demographic and clinical characteristics of ClinVar LP/P neurological variant carriers versus non-carriers among the Irish and AnswerALS cohorts.** Values are shown as percentages (*n*) for categorical variables and mean ± 95% confidence interval (*n*) for continuous variables. For categorical variables, the *n* in parentheses is the count corresponding to the percentage in that cell, not the total cohort size. For continuous variables, the *n* reported is the number of individuals with available data for that measure. Cohort differences between categorical variables were evaluated using $\chi^2$ tests (without Yates' correction if all expected cell counts ≥ 10; with Yates' correction if any expected cell count 5-9), and Fisher's exact tests when any expected cell count <5. Continuous variables were assessed via Welch's two sample t-tests. For categorical variables, effect size is given as Φ for 2x2 tables and Cramér's V for tables with more than two categories. For Fisher's exact test, Φ was calculated from the Pearson chi-squared statistic. For continuous variables, effect size is reported as Cohen's *d*. Across the Irish cohort, a Bonferroni-corrected significance threshold of $p < 0.00263$ was applied for 19 independent comparisons. In the AnswerALS cohort, a Bonferroni-corrected significance threshold of $p < 0.00294$ was applied for 17 independent comparisons. For p values that are <0.05 but equal to or above the Bonferroni-corrected significance threshold for the relevant cohort, results are described as nominally significant only in text.

| ***Irish Cohort*** | | | | | | | | | |
|---|---|---|---|---|---|---|---|---|---|
| ***Variable*** | ***Category*** | ***Count (%) or mean ± 95% CI with Neurological Variant (n)*** | ***Count (%) or mean ± CI without Neurological Variant (n)*** | ***Variable Type*** | ***Statistical Test*** | ***Test Statistic*** | ***df*** | ***p-value*** | ***Effect Size*** |
| Sex | Female | 34.0% (n=16) | 41.2% (n=170) | Categorical | Chi-Squared | $\chi^2 \sim 0.9$ | 1 | 0.379 | Φ=0.04 |
| Site of Onset | Spinal | 70.2% (n=33) | 69.8% (n=291) | Categorical | Chi-Squared | $\chi^2 \sim 0.004$ | 1 | 0.952 | Φ=0.003 |
| Site of Onset | Bulbar | 19.1% (n=9) | 29.0% (n=121) | Categorical | Chi-Squared | $\chi^2 \sim 2.0$ | 1 | 0.153 | Φ=0.07 |
| Site of Onset | Cognitive/Behavioural | 2.1% (n=1) | 4.3% (n=18) | Categorical | Fisher | NA | NA | 0.708 | Φ=0.03 |
| Site of Onset | Thoracic/Respiratory | 10.6% (n=5) | 1.2% (n=5) | Categorical | Fisher | NA | NA | 0.002 | Φ=0.20 |

| | | | | | | | | | |
|---|---|---|---|---|---|---|---|---|---|
| El Escorial | Definite | 48.9% (n=22) | 55.1% (n=205) | Categorical | Chi-Squared | $\chi^2 \sim 3.5$ | 3 | 0.323 | Cramér's V= 0.09 |
| El Escorial | Probable | 22.2% (n=10) | 27.4% (n=102) | – | – | – | – | – | – |
| El Escorial | Possible | 26.7% (n=12) | 16.1% (n=60) | – | – | – | – | – | – |
| El Escorial | Suspected | 2.2% (n=1) | 1.34% (n=5) | – | – | – | – | – | – |
| Family History | ALS | 23.4% (n=11) | 20.3% (n=83) | Categorical | Chi-Squared | $\chi^2 \sim 0.2$ | 1 | 0.624 | Φ=0.02 |
| Family History | FTD | 4.25% (n=2) | 3.43% (n=14) | Categorical | Fisher | NA | NA | 0.676 | Φ=0.01 |
| Family History | Neurological | 48.9% (n=23) | 51.7% (n=211) | Categorical | Chi-Squared | $\chi^2 \sim 0.1$ | 1 | 0.718 | Φ=0.02 |
| Family History | Psychiatric | 53.2% (n=25) | 55.1% (n=225) | Categorical | Chi-Squared | $\chi^2 \sim 0.1$ | 1 | 0.799 | Φ=0.01 |
| Baseline ALSFRS(R) | – | 36.85 ± 2.22 (n=39) | 36.71 ± 0.75 (n=336) | Continuous | T-Test (Welch) | $t \sim 0.1$ | ~47 | 0.907 | Cohen's D = 0.02 |
| Baseline ECAS Score | – | 97.91 ± 6.73 (n=22) | 96.56 ± 2.79 (n=188) | Continuous | T-Test (Welch) | $t \sim 0.4$ | ~29 | 0.719 | Cohen's D = 0.07 |
| ECAS Ever Abnormal | Ever Abnormal | 27.3% (n=6) | 37.7% (n=71) | Categorical | Chi-Squared | $\chi^2 \sim 0.9$ | 1 | 0.334 | Φ=0.07 |
| Pre-MND BBI Baseline Score | – | 0.92 ± 0.56 (n=13) | 3.08 ± 0.97 (n=130) | Continuous | T-Test (Welch) | $t \sim -3.8$ | ~105 | 0.0003 | Cohen's $d$=-0.40 |
| Post-MND BBI Baseline Score | – | 5.50 ± 2.30 (n=28) | 9.78 ± 1.59 (n=229) | Continuous | T-Test (Welch) | $t \sim -3.0$ | ~57 | 0.004 | Cohen's $d$=-0.36 |
| Pre-MND BBI | Ever Abnormal | 7.14% (n=2) | 10.5% (n=24) | Categorical | Fisher | NA | NA | 0.7497 | Φ=0.03 |

| | | | | | | | | | |
|---|---|---|---|---|---|---|---|---|---|
| Ever Abnormal | | | | | | | | | |
| Post-MND BBI Ever Abnormal | Ever Abnormal | 50.0% (n=14) | 52.0% (n=119) | Categorical | Chi-Squared | $\chi^2 \sim 0.04$ | 1 | 0.844 | Φ=0.01 |
| Age of Symptom Onset (Years) | – | 62.25 ± 3.44 (n=47) | 63.04 ± 1.07 (n=407) | Continuous | T-Test (Welch) | $t \sim -0.4$ | ~55 | 0.669 | Cohen's D=-0.07 |
| Survival (Symptom Onset to Death)(Months) | – | 49.87 ± 10.03 (n=47) | 44.16 ± 3.47 (n=398) | Continuous | T-Test (Welch) | $t \sim 1.1$ | ~58 | 0.296 | Cohen's D=0.16 |
| ***AnswerALS Cohort*** | | | | | | | | | |
| ***Variable*** | ***Category*** | ***Count % or mean ± CI with Neurological Variant (n)*** | ***Count % or mean ± CI without Neurological Variant (n)*** | ***Variable Type*** | ***Statistical Test*** | ***Test Statistic*** | ***df*** | ***p-value*** | ***Effect Size*** |
| Sex | Female | 28.9% (n=20) | 38.0% (n=266) | Categorical | Chi-Squared | $\chi^2 \sim 2.7$ | 1 | 0.103 | Φ=0.06 |
| Site of Onset | Spinal | 68.1% (n=47) | 68.3% (n=479) | Categorical | Chi-Squared | $\chi^2 \sim 0.004$ | 1 | 0.947 | Φ=0.002 |
| Site of Onset | Bulbar | 24.6% (n=17) | 21.9% (n=151) | Categorical | Chi-Squared | $\chi^2 \sim 0.07$ | 1 | 0.785+ | Φ=0.01 |
| Site of Onset | Axial | 1.4% (n=1) | 1.9% (n=13) | Categorical | Fisher | NA | NA | 1.000 | Φ=0.02 |
| El Escorial | Definite | 31.5% (n=23) | 29.8% (n=209) | Categorical | Chi-Squared | $\chi^2 \sim 0.7$ | 3 | 0.867 | Cramér's V=0.03 |
| El Escorial | Probable | 46.6% (n=32) | 51.1% (n=357) | Categorical | – | – | – | – | – |

| | | | | | | | | | |
|---|---|---|---|---|---|---|---|---|---|
| El Escorial | Possible | 15.9% (n=11) | 15.8% (n=110) | Categorical | – | – | – | – | – |
| El Escorial | Suspected | 4.2% (n=3) | 3.5% (n=24) | Categorical | – | – | – | – | – |
| Ethnic Category | White - Not Hispanic or Latino (%) | 92.8% (n=64) | 88.9% (n=618) | Categorical | Chi-Squared (with Yate's Correction) | $\chi^2$~0.01 | 1 | 0.909 | Φ=0.004 |
| Ethnic Category | Black/African American - Not Hispanic or Latino (%) | 5.6% (n=4) | 4.5% (n=32) | Categorical | Fisher | NA | NA | 0.556 | Φ=0.02 |
| Ethnic Category | White - Hispanic or Latino | 1.4% (n=1) | 4.8% (n=33) | Categorical | Fisher | NA | NA | 0.357 | Φ=0.05 |
| Ethnic Category | Asian - Not Hispanic or Latino | 1.4% (n=1) | 1.6% (n=11) | Categorical | Fisher | NA | NA | 1.000 | Φ=0.004 |
| Ethnic Category | Other | 1.4% (n=1) | 1.4% (n=10) | Categorical | Fisher | NA | NA | 1.000 | Φ=0.0007 |
| Family History | ALS | 16.2% (n=11) | 13.0% (n=81) | Categorical | Chi-Squared | $\chi^2$~0.5 | 1 | 0.468 | Φ=0.03 |
| Family History | FTD | 0 (0) | 0.6% (n=4) | Categorical | Fisher | NA | NA | 1.000 | Φ=0.03 |
| Family History | Neurological | 39.1% (n=27) | 42.2% (n=263) | Categorical | Chi-Squared | $\chi^2$~0.01 | 1 | 0.913 | Φ=0.004 |
| Family History | Psychiatric | 8.7% (n=6) | 11.1% (n=68) | Categorical | Chi-Squared (with Yates' correction) | $\chi^2$~4.2e-30 | 1 | 1.000 | Φ=7.85e-17 |
| Baseline ALSFRS(R) | – | 35.33 ± 2.21 (n=69) | 33.94 ±0.63 (n=698) | Continuous | T-Test (Welch) | *t* ~ 0.346 | ~92 | 0.7328 | Cohen's *d*=0.04 |

| | | | | | | | | | |
|---|---|---|---|---|---|---|---|---|---|
| CBS Cognitive Score Baseline | – | 16.18 ± 0.79 (n=61) | 15.99 ± 0.26 (n=583) | Continuous | T-Test (Welch) | *t* ~ 0.5 | ~74 | 0.639 | Cohen's *d*=0.06 |
| CBS Cognitive Score Ever Abnormal | Ever Abnormal | 44.3% (n=27) | 40.0% (n=233) | Categorical | Chi-Squared | $\chi^2$~0.4 | 1 | 0.515 | Φ=0.03 |
| CBS Behavioural Score Baseline | – | 38.76 ± 2.19 (n=50) | 37.77 ± 0.70 (n=497) | Continuous | T-Test (Welch) | *t* ~ 0.8 | ~59 | 0.402 | Cohen's *d*=0.12 |
| CBS Behavioural Score Ever Abnormal | Ever Abnormal | 32.0% (n=16) | 38.8% (n=193) | Categorical | Chi-Squared | $\chi^2$~0.9 | 1 | 0.343 | Φ=0.04 |
| Age of Symptom Onset (Years) | – | 56.20 ± 2.63 (n=71) | 56.58 ± 0.87 (n=698) | Continuous | T-Test (Welch) | *t* ~ 0.3 | ~94 | 0.791 | Cohen's *d*=0.03 |
| Survival (Symptom Onset to Death)(Months) | – | 44.37 ± 9.44 (n=37) | 43.36 ± 3.32 (n=324) | Continuous | T-Test (Welch) | *t* ~ 0.2 | ~45 | 0.844 | Cohen's *d*=0.03 |

**Supplemental Table S2.3. Clinical characteristics of ClinVar LP/P hereditary spastic paraplegia variant carriers versus non-carriers among individuals with amyotrophic lateral sclerosis in the Irish and AnswerALS cohorts.** Values are shown as percentages (*n*) for categorical variables and mean ± 95% confidence interval (*n*) for continuous variables. For categorical variables, the *n* in parentheses is the count corresponding to the percentage in that cell, not the total cohort size. For continuous variables, the *n* reported is the number of individuals with available data for that measure. Cohort differences between categorical variables were evaluated using $\chi^2$ tests (without Yates' correction if all expected cell counts ≥ 10; with Yates' correction if any expected cell count 5-9), and Fisher's exact tests when any expected cell count <5. El Escorial categories were collapsed into binary variables (Definite/Probable vs Other) for analysis. This was done as certain categories had cell count <5 in carriers in both Irish and AnswerALS, making the chi-square test invalid.

Continuous variables were assessed via Welch's two sample t-tests. For categorical variables, effect size is given as Φ for 2x2 tables and Cramér's V for tables with more than two categories. For Fisher's exact test, Φ was calculated from the Pearson chi-squared statistic. For continuous variables, effect size is reported as Cohen's *d*. Across both cohorts, a Bonferroni-corrected significance threshold of $p < 0.013$ was applied for 4 independent comparisons. For p values that are <0.05 but equal to or above the Bonferroni-corrected significance threshold for the relevant cohort, results are described as nominally significant only in text.

| ***Irish Cohort*** | | | | | | | | | |
|---|---|---|---|---|---|---|---|---|---|
| ***Variable*** | ***Category*** | ***Count (%) or mean ± 95% CI with HSP Variant (n)*** | ***Count (%) or mean ± CI without HSP Variant (n)*** | ***Variable Type*** | ***Statistical Test*** | ***Test Statistic*** | ***df*** | ***p-value*** | ***Effect Size*** |
| Site of Onset | Spinal | 80.0 (n=12) | 69.7 (n=306) | Categorical | Fisher | – | – | 0.569 | Φ=0.03 |
| El Escorial | Definite | 53.3 (n=8) | 54.6 (n=216) | Categorical | Fisher | – | – | 0.043 | Φ=0.09 |
| El Escorial | Probable | 6.7 (n=1) | 27.9 (n=112) | Categorical | – | – | – | – | – |
| El Escorial | Possible | 40.0 (n=6) | 16.0 (n=64) | Categorical | – | – | – | – | – |
| El Escorial | Suspected | 0 (n=0) | 1.5 (n=6) | Categorical | – | – | – | – | – |
| Age of Symptom Onset (Years) | – | 57.62 ± 6.65 (n=15) | 63.14 ± 1.05 (n=439) | Continuous | T-Test (Welch) | *t* ~ 1.8 | ~15 | 0.100 | Cohen's *d*= -0.50 |
| Survival (Symptom Onset to Death)(Months) | – | 49.40 ± 21.85 (n=15) | 44.60 ± 3.30 (n=430) | Continuous | T-Test (Welch) | *t* ~ -0.5 | ~15 | 0.653 | Cohen's *d*=0.14 |
| ***AnswerALS Cohort*** | | | | | | | | | |
| ***Variable*** | ***Category*** | ***Count % or mean*** | ***Count % or mean ±*** | ***Variable*** | ***Statistical*** | ***Test Statistic*** | ***df*** | ***p-value*** | ***Effect Size*** |

| | | ± CI with HSP Variant (n) | CI without HSP Variant (n) | Type | Test | | | | |
|---|---|---|---|---|---|---|---|---|---|
| Site of Onset | Spinal | 66.7 (n=14) | 68.6 (n=512) | Categorical | Chi-Squared | $\chi^2$~2.3e-30 | 1 | 1.000 | Φ=5.43e-17 |
| El Escorial | Definite | 42.9 (n=9) | 29.9 (n=223) | Categorical | Fisher | – | – | 1.000 | Φ=6.91e-16 |
| El Escorial | Probable | 38.1 (n=8) | 51.1 (n=381) | Categorical | – | – | – | – | – |
| El Escorial | Possible | 19.0 (n=4) | 15.7 (n=117) | Categorical | – | – | – | – | – |
| El Escorial | Suspected | 0 (n=0) | 3.3 (n=24) | Categorical | – | – | – | – | – |
| Age of Symptom Onset (Years) | – | 56.81 ± 4.20 (n=22) | 56.53 ± 0.87 (n=730) | Continuous | T-Test (Welch) | *t* ~ 0.1 | ~750 | 0.914 | Cohen's *d*=0.02 |
| Survival (Symptom Onset to Death)(Months) | – | 42.80 ± 19.20 (n=14) | 43.50 ± 3.20 (n=339) | Continuous | T-Test (Welch) | *t* ~ 0.1 | ~14 | 0.942 | Cohen's *d*=-0.02 |

**Supplemental Table S2.4. Demographic and clinical characteristics of ClinVar LP/P neurodevelopmental variant carriers versus non-carriers among the Irish and AnswerALS cohorts.** Values are shown as percentages (*n*) for categorical variables and mean ± 95% confidence interval (*n*) for continuous variables. For categorical variables, the *n* in parentheses is the count corresponding to the percentage in that cell, not the total cohort size. For continuous variables, the *n* reported is the number of individuals with available data for that measure. Cohort differences between categorical variables were evaluated using $\chi^2$ tests (without Yates' correction if all expected cell counts ≥ 10; with Yates' correction if any expected cell count 5-9), and Fisher's exact tests when any expected cell count <5. Continuous variables were assessed via Welch's two sample t-tests. For categorical variables, effect size is given as Φ for 2x2 tables and Cramér's V for tables with more than two categories. For Fisher's exact test, Φ was calculated from the Pearson chi-squared statistic. For continuous variables, effect size is reported as

Cohen's *d*. Across the Irish cohort, a Bonferroni-corrected significance threshold of $p < 0.00263$ was applied for 19 independent comparisons. In the AnswerALS cohort, a Bonferroni-corrected significance threshold of $p < 0.00294$ was applied for 17 independent comparisons. For p values that are <0.05 but equal to or above the Bonferroni-corrected significance threshold for the relevant cohort, results are described as nominally significant only in text.

| ***Irish*** | | | | | | | | | |
|---|---|---|---|---|---|---|---|---|---|
| ***Variable*** | ***Category*** | ***Count (%) or mean ± 95% CI With Neurodevelopmental Variant (n)*** | ***Count (%) or mean ± CI Without Neurodevelopmental Variant (n)*** | ***Variable Type*** | ***Statistical Test*** | ***Test Statistic*** | ***df*** | ***p-value*** | ***Effect Size*** |
| Sex | Female | 50.0 (n=6) | 39.9 (n=176) | Categorical | Chi-Squared (with Yate's correction) | $\chi^2 \sim 0.2$ | 1 | 0.618 | Φ=0.04 |
| Site of Onset | Spinal | 50.0 (n=6) | 70.8 (n=313) | Categorical | Fisher | NA | NA | 0.062 | Φ=0.10 |
| Site of Onset | Bulbar | 50.0 (n=6) | 26.9 (n=119) | Categorical | Fisher | NA | NA | 0.062 | Φ=0.10 |
| El Escorial | Definite | 66.7 (n=8) | 53.8 (n=218) | Categorical | Chi-Squared | $\chi^2 \sim 2.2$ | 3 | 0.699 | Cramér's V=0.07 |
| El Escorial | Probable | 25.0 (n=3) | 27.7 (n=112) | Categorical | – | – | – | – | – |
| El Escorial | Possible | 8.3 (n=1) | 17.0 (n=69) | Categorical | – | – | – | – | – |
| El Escorial | Suspected | 0 | 1.5 (n=6) | Categorical | – | – | – | – | – |
| Family History | ALS | 25.0 (n=3) | 20.8 (n=91) | Categorical | Fisher | NA | NA | 0.737 | Φ=0.01 |

| | | | | | | | | | |
|---|---|---|---|---|---|---|---|---|---|
| Family History | FTD | 0 | 3.6 (n=16) | Categorical | Fisher | NA | NA | 1 | Φ=0.0 |
| Family History | Neurological | 53.8 (n=7) | 51.4 (n=227) | Categorical | Chi-Square | $\chi^2 \sim 0.03$ | 1 | 0.860 | Φ=0.008 |
| Family History | Psychiatric | 69.2 (n=9) | 54.5 (n=241) | Categorical | Fisher | NA | NA | 0.400 | Φ=0.04 |
| Baseline ALSFRS(R) | – | 38.00 ± 4.85 (n=9) | 36.69 ± 0.72 (n=366) | Continuous | T-Test (Welch) | t=0.5 | ~8 | 0.613 | Cohen's *d*=0.19 |
| Baseline ECAS Score | – | 91.00 ± 13.63 (n=7) | 96.89 ± 2.63 (n=203) | Continuous | T-Test (Welch) | t=-0.8 | ~6 | 0.435 | Cohen's d=-0.31 |
| ECAS Ever Abnormal | Abnormal | 42.9 (n=3) | 33.0 (n=67) | Categorical | Fisher | NA | NA | 0.688 | Φ=0.01 |
| Pre-MND BBI Baseline Score | – | 3.14 ± 3.93 (n=7) | 2.88 ± 0.92 (n=136) | Continuous | T-Test (Welch) | t=0.1 | ~7 | 0.900 | Cohne's d=0.05 |
| Post-MND BBI Baseline Score | – | 11.62 ± 10.28 (n=8) | 9.23 ± 1.45 (n=245) | Continuous | T-Test (Welch) | t=0.5 | ~7 | 0.665 | Cohne's d=0.21 |
| Pre-MND BBI Ever Abnormal | Abnormal | 12.5 (n=3) | 10.0 (n=25) | Categorical | Fisher | NA | NA | 0.579 | Φ=0.04 |
| Post-MND BBI Ever Abnormal | Abnormal | 62.5 (n=5) | 51.4 (n=128) | Categorical | Fisher | NA | NA | 0.724 | Φ=0.02 |
| Age of Symptom Onset (Years) | – | 64.9 ± 5.80 (n=12) | 62.92 ± 1.01 (n=442) | Continuous | T-Test (Welch) | t=-0.7 | ~13 | 0.487 | Cohen's *d*=0.16 |
| Survival (Symptom Onset to Death)(Months) | – | 26.9 ± 9.10 (n=12) | 45.12 ± 3.25 (n=433) | Continuous | T-Test (Welch) | $t \sim -4.5$ | ~18 | 0.0003 | Cohen's *d* = -0.52 |

| | | | | | | | | | |
|---|---|---|---|---|---|---|---|---|---|
| Survival (time-to-event) | – | – | – | – | Log-rank | $\chi^2 = 9.5$ | 1 | 0.005 | HR = 2.25 (95% CI 1.26-4.00) |
| **Model in Irish Cohort** | **Group Definition** | **HR (ND/NP Variant Carrier vs Non-carrier)** | **95% CI** | **p-value** | **Proportional Hazards (Schoenfeld)** | | **Notes** | | |
| Cox, adjusted for bulbar onset | ND variant carrier vs non-carrier | 2.11 | 1.21-3.69 | 0.009 | Variant $\chi^2$ =1.39, p=0.24; global p=0.24 | | Hazard higher in carriers, PH assumption supported. Bulbar onset included as covariate. | | |
| Interaction | Variant vs bulbar onset | – | – | – | Interaction HR=0.97, p=0.96 | | Interaction between variant status and bulbar onset was tested and found non-significant, indicating the effect of variants is independent of bulbar onset. | | |
| ***AnswerALS*** | | | | | | | | | |
| ***Variable*** | ***Category*** | ***Count or mean ± CI With Neurodevelopmental Variant (n)*** | ***Count or mean ± CI Without Neurodevelopmental Variant (n)*** | ***Variable Type (Continuous/ Categorical)*** | ***Statistical Test*** | ***Test Statistic*** | ***df*** | ***p-value*** | ***Effect Size*** |
| Sex | Female | 45.0 (n=9) | 36.9 (n=278) | Categorical | Chi-Squared (with Yate's correction) | $\chi^2$~0.6 | 1 | 0.458 | Φ=0.03 |
| Site of Onset | Spinal | 65.0 (n=13) | 68.0 (n=513) | Categorical | Chi-Squared (with Yate's | $\chi^2$~0.002 | 1 | 0.965 | Φ=0.01 |

| | | | | | | | | | |
|---|---|---|---|---|---|---|---|---|---|
| | | | | | correction) | | | | |
| Site of Onset | Bulbar | 35.0 (n=7) | 21.4 (n=161) | Categorical | Chi-Squared (with Yate's correction) | $\chi^2$~0.8 | 1 | 0.380 | Φ=0.04 |
| El Escorial | Definite | 30.0 (n=6) | 30.3 (n=226) | Categorical | Chi-Squared | $\chi^2$ ~ 1.1 | 3 | 0.894 | Cramér's V=0.04 |
| El Escorial | Probable | 50.0 (n=10) | 50.8 (n=379) | Categorical | – | – | – | – | – |
| El Escorial | Possible | 20.0 (n=4) | 15.7 (n=117) | Categorical | – | – | – | – | – |
| El Escorial | Suspected | 0 | 3.2 (n=24) | Categorical | – | – | – | – | – |
| Family History | ALS | 5.3 (n=1) | 13.0 (n=91) | Categorical | Fisher | NA | NA | 0.494 | Φ=0.04 |
| Family History | FTD | 0 | 0.57 (n=4) | Categorical | Fisher | NA | NA | 1.000 | Φ=0.01 |
| Family History | Neurological | 52.6 (n=10) | 40.1 (n=280) | Categorical | Chi-square | $\chi^2$ ~ 1.2 | 1 | 0.270 | Φ=0.04 |
| Family History | Psychiatric | 5.3 (n=1) | 10.4 (n=73) | Categorical | Fisher | NA | NA | 0.710 | Φ=0.03 |
| CBS Cognitive Score Baseline | – | 15.82 ± 1.47 (n=17) | 16.01 ± 0.25 (n=627) | Continuous | T-Test (Welch) | *t* ~ -0.2 | ~17 | 0.810 | Cohen's d = -0.06 |
| CBS Behavioural Score Baseline | – | 41.50 ± 1.89 (n=12) | 37.78 ± 0.68 (n=535) | Continuous | T-Test (Welch) | *t* ~ 3.6 | ~14 | 0.003 | Cohen's d = 0.47 |
| CBS Cognitive Score Ever Abnormal | Abnormal | 58.8 (n=10) | 59.7 (n=374) | Categorical | Chi-square | $\chi^2$ ~ 0.005 | 1 | 0.945 | Φ = 0.003 |

| | | | | | | | | | |
|---|---|---|---|---|---|---|---|---|---|
| CBS Behavioural Score Ever Abnormal | Abnormal | 8.3 (n=1) | 38.9 (n=208) | Categorical | Fisher | NA | NA | 0.035 | Φ= 0.09 |
| Age of Symptom Onset (Years) | – | 62.05 ± 3.45 (n=20) | 56.40 ± 0.84 (n=738) | Continuous | T-Test (Welch) | $t$ ~ 3.1 | ~21 | 0.005 | Cohen's d =0.49 |
| Baseline ALSFRS(R) | – | 34.00 ± 3.30 (n=19) | 33.97 ± 0.62 (n=731) | Continuous | T-Test (Welch) | $t$ ~ 0.02 | ~19 | 0.987 | Cohen's d = 0.003 |
| Survival (Symptom Onset to Death)(Months) | – | 50.79 ± 18.88 (n=13) | 52.79 ± 3.17 (n=463) | Continuous | T-Test (Welch) | $t$~0.2 | ~13 | 0.832 | Cohen's d = -0.06 |
| Survival (time-to-event) | – | – | – | – | Log-rank | $\chi^2$ = 0.01 | 1 | 0.909 | HR = 1.03 (95% CI 0.59- 1.79) |
| Combined Survival | | | | | | | | | |
| Combined Survival Both Cohort (time-to-event) | – | – | – | – | Log-rank | $\chi^2$ = 3.02 | 1 | 0.082 | HR= 1.41 (95% CI 0.96-2.09) |

**Supplemental Table S2.5 Combined cox-proportional hazards for survival in neurodevelopmental variant carriers.**

| *Model* | *HR (Variant vs Non-Variant Carrier)* | *95% CI* | *p-value* | *Proportional Hazards Assumption (Schoenfeld)* | *Notes* |
|---|---|---|---|---|---|

| | | | | | |
|---|---|---|---|---|---|
| Cox (adjusted for cohort) | 1.82 | 1.20-2.76 | 0.005 | Global test indicated non-proportional hazards due to cohort. | Adjusted for cohort; PH violation for cohort motivated use of stratified model. |
| Cox (stratified by cohort) | 1.80 | 1.20-2.73 | 0.006 | Variant $\chi^2$ = 1.39, p=0.24; global p=0.24 (no PH violation) | Baseline hazards allowed to differ by cohort; PH assumption for Variant supported |
| Cox with variant x cohort interaction | 1.35 (Variantt), 1.84 (interaction Variant x Cohort) | 0.72-2.53 (variant), 0.80-4.25 (interaction) | 0.35 (variant), 0.15 (interaction) | PH passes for Variant (p=0.16) and Variant x Cohort (p=0.98); PH violated for Cohort (p=0.03); global p=0.03 | No significant interaction. The effect of Variant appears consistent across cohorts, but Cohort shows some non-proportional hazards. |
| Combined cox proportional hazards for survival in neurodevelopmental variant carriers | | | | | |
| Model | HR (Variant vs Non-Variant Carrier) | 95% CI | p-value | Proportional Hazards Assumption (Schoenfeld) | Notes |
| Cox (adjusted for cohort) | 1.48 | 0.99-2.19 | 0.05 | Variant $\chi^2$ =0.45, p=0.50; Cohort $\chi^2$=11.74, p=0.0006; global $\chi^2$=12.64, p=0.002 | Adjusted for cohort; PH holds for Variant but violated for Cohort, motivating stratified model. |

| | | | | | |
|---|---|---|---|---|---|
| Cox (stratified by cohort) | 1.44 | 0.970-2.126 | 0.070 | Variant $\chi^2$ =0.8, p=0.370 (no PH violation) | Baseline hazards differ by cohort; PH fully supported for Variant. |
| Cox with variant x cohort interaction | 1.03 (Variant), 2.48 (interaction Variant x Irish with WGS) | 0.60-1.80 (variant), 1.13-5.42 (interaction) | 0.91 (variant), 0.02 (interaction) | Use stratified for primary; interaction model shows heterogeneity | Significant interaction indicates stronger Variant effect in the Irish cohort; directionally consistent HRs >1. |
| Survival by neurodevelopmental only vs neurodevelopmental + neuromuscular variant subgroups (combined Irish and answerals cohorts, cohort-stratified Cox models) | | | | | |
| Comparison/Model | Group definition | HR (vs reference) | 95% CI | p-value | Notes |
| Three group stratified Cox (non-variant reference) | ND only | 1.28 | 0.77-2.14 | 0.35 | Stratified by cohort; no significant difference vs non-carriers (global Ward p=0.20) |
| | ND plus NM | 1.62 | 0.91-2.87 | 0.10 | Trend toward shorter survival, but not statistically significant. |
| Direct contrast (ND only reference) | Non-variant | 0.78 | 0.47-1.31 | 0.35 | Reverse of ND only vs non-variant comparison above. |

| | | | | | |
|---|---|---|---|---|---|
| | ND plus NM | 1.27 | 0.59-2.72 | 0.54 | No evidence that ND plus NM variant carriers differ from ND only variant carriers. |
| Carriers only, stratified Cox (ND only reference) | ND plus NM | 0.76 | 0.31-1.85 | 0.54 | Analysis restricted to carriers (n=27 events); no survival difference between carrier subgroups. |
| PH assumption (three-group model) | Group term | – | – | 0.04 | Global Schoenfeld test suggests some non-proportionality for the grouped exposure; interpret subgroup HRs cautiously. |

**Supplemental Table 2.6 ClinVar variant-set enrichment using SKAT-O (matrix-defined sets, cohort-adjusted) with allele frequency thresholds (gnomAD, NFE).**

| Variant-set category | Cases with ≥1 Variant, n/N (%) | Controls with ≥1 Variant, n/N (%) | SKAT-O p-value |
|---|---|---|---|
| AF NFE ≤ 0.01 | | | |
| Neurological (non-ALS) | 118/1243 (9.5) | 32/323 (9.9) | 0.832 |

| | | | |
|---|---|---|---|
| Neurodevelopmental and Neuropsychiatric | 33/1243 (2.7) | 9/323 (2.8) | 0.845 |
| Hereditary Spastic Paraplegia | 36/1243 (2.9) | 9/323 (2.8) | 1.000 |
| AF NFE ≤ 0.001 | | | |
| Neurological (non-ALS) | 64/1243 (5.1) | 21 (6.5) | 0.336 |
| Neurodevelopmental and Neuropsychiatric | 22/1243 (1.8) | 2/323 (0.6) | 0.201 |
| Hereditary Spastic Paraplegia | 23/1243 (1.9) | 5/323 (1.5) | 1.000 |
| AF NFE ≤ 0.0001 | | | |
| Neurological (non-ALS) | 26/1243 (2.1) | 10/323 (3.1) | 0.297 |
| Neurodevelopmental and Neuropsychiatric | 7/1243 (0.6) | 1/323 (0.3) | 1.000 |
| Hereditary Spastic Paraplegia | 9/1243 (0.7) | 1/323 (0.3) | 0.697 |

**Supplemental Table S2.7. Irish ALS neurodevelopmental variant carriers vs non-carriers: family history burden**

| Neurodevelopmental/Neuropsychiatric Variant Carriers | | | | | | | | | |
|---|---|---|---|---|---|---|---|---|---|
| Metric | n Carrier | n Non-Carrier | Mean Carrier | Mean Non-Carrier | Wilcoxon p | NB IRR | NB IRR (low) | NB IRR (high) | NB p-value |
| n_np_rel | 11 | 411 | 1.91 | 1.32 | 0.11 | 2.41 | 1.12 | 5.18 | 0.025 |

| | | | | | | | | | |
|---|---|---|---|---|---|---|---|---|---|
| np_rel_prop | 11 | 411 | 0.297 | 0.099 | 0.083 | – | – | – | – |
| n_neuro_rel | 11 | 411 | 0.7 | 1.1 | 0.375 | – | – | – | – |
| neuro_rel_prop | 11 | 411 | 0.1 | 0.1 | 0.694 | – | – | – | – |
| total_family_size | 11 | 411 | 11.4 | 14.1 | 0.157 | – | – | – | – |
| Neurological Variant Carriers | | | | | | | | | |
| Metric | N Carrier | N Non-Carrier | Mean Carrier | Mean Non-Carrier | Wilcoxon p | NB IRR | NB IRR (low) | NB IRR (high) | NB p-value |
| n_neuro_rel | 31 | 300 | 1.1 | 1.1 | 0.771 | 0.99 | 0.67 | 1.47 | 0.968 |
| neuro_rel_prop | 31 | 300 | 0.1 | 0.1 | 0.960 | – | – | – | – |
| n_np_rel | 31 | 300 | 1.5 | 1.7 | 0.336 | – | – | – | – |
| np_rel_prop | 31 | 300 | 0.1 | 0.1 | 0.355 | – | – | – | – |
| total_family_size | 31 | 300 | 14.3 | 14.5 | 0.782 | – | – | – | – |

Family history data were obtained from the Irish ALS Register. Only first- and second- degree relatives were included. Statistical analysis included Wilcoxon rank-sum tests to compare carrier vs non-carrier distributions. Negative binomial regression (NB IRR) models count of affected relatives with log family size as offset; Poisson models were rejected due to overdispersion. IRR: Incidence rate ratio; 95% CI shown for NB IRR.

N_np_rel: number of neurodevelopmental/neuropsychiatric relatives per individual

Np_rel_prop: proportion of neurodevelopmental/neuropsychiatric relatives (relative count/total family size)

N_neuro_rel: number of neurological relatives

Total_family_size: sum of reported brothers, sisters, paternal uncles/aunts, and maternal uncles/aunts
