## Supplementary material for "Rare neurological and neurodevelopmental variants in ALS link to onset, survival and family history": S3

Supplemental Table S3.1 and S3.2 Navigation:

Table S3.1: Neurologic Variant Information

Table S3.2: Individual Neurologic Variant Carrier Information

Supplemental Table S3.1. Information on LP/P non-ALS neurologic-associated variants identified in Irish (n=47) and AnswerALS (n=69) pwALS. ClinVar IDs are listed with molecular consequence, inheritance patterns for associated genes from OMIM (AR: autosomal recessive; AD: autosomal dominant), variant associated phenotypes, and population allele frequencies. Population frequency values reported from TOPMed, gnomAD, and 1000 Genomes Project (1000G) are provided where available. Cohort-specific allele frequencies were calculated assuming 2 alleles per individual using the following denominators: Irish cases (n=469, 938 alleles), Irish controls (n=230, 460 alleles), AnswerALS cases (n=774; 1548 alleles), AnswerALS controls (n=94; 188 alleles). All pwALS carrying variants in this dataset were heterozygotes. Case-level clinical and neuropsychological characteristics for these variant carriers can be cross-referenced in Supplementary Table S3.2.

| ClinVar Variant ID | Gene | Molecular Consequence | Inheritance Pattern for associated Gene | Associated Phenotype with Variant | Population Frequency | Irish Case ID | AnswerALS Case ID |
|---|---|---|---|---|---|---|---|
| 4290 | *GBA1* | missense | AD | Primary phenotype considered in current analysis:<br>Parkinson disease;<br>Lewy Body Dementia<br><br>Other phenotypes associated with this variant:<br>Gaucher disease; Akinesia Rigidity | TOPMed: 0.00195<br>gnomAD: 0.00191<br>1000G: 0.00060<br>Irish ALS: 0.0011<br>AnswerALS ALS: 0.0045<br>Irish Controls: 0<br>AnswerALS Controls: 0 | I_1 | AA_1<br>AA_2<br>AA_3<br>AA_4<br>AA_5<br>AA_6<br>AA_7 |

| | | | | | | | |
|---|---|---|---|---|---|---|---|
| 7050 | *PRKN* | missense | AR | Primary phenotype considered in current analysis:<br>Autosomal recessive juvenile Parkinson disease; Parkinson Disease<br><br>Other phenotypes associated with this variant:<br>-- | TOPMed: 0.00213<br>gnomAD: 0.00226<br>1000G: 0.00040<br>Irish ALS: 0.0053<br>AnswerALS ALS: 0.0039<br>Irish Controls: 0.0065<br>AnswerALS Controls: 0 | I_2<br>I_3<br>I_4<br>I_5<br>I_6 | AA_8<br>AA_9<br>AA_10<br>AA_11<br>AA_12<br>AA_13 |
| 641981 | *FBXO7* | frameshift | AR | Primary phenotype considered in current analysis:<br>Parkinsonian-Pyramidal Syndrome<br><br>Other phenotypes associated with this variant:<br>-- | TOPMed: –<br>gnomAD v4: *chr22-32498165-C-CAA*<br>Joint Max Group AF *0.0000994 (NFE)*<br>Genomes Max Group AF *0.00006804 (NFE)*<br>Exomes Max Group AF *0.00009754 (NFE)*<br>1000G: –<br>Irish ALS: 0.00213<br>AnswerALS ALS: 0.0006<br>Irish Controls: 0<br>AnswerALS Controls: 0 | I_7<br>I_8 | AA_14 |
| 444787 | *SURF1* | Frameshift and 5 prime UTR | AR | Primary phenotype considered in current analysis:<br>Charcot-Marie-Tooth disease type<br><br>Other phenotypes associated with this variant:<br>Mitochondrial complex deficiency, nuclear | TOPMed: –<br>gnomAD: 0.00036<br>1000G: –<br>Irish ALS: 0.0011<br>AnswerALS ALS: 0.0006<br>Irish Controls: 0<br>AnswerALS Controls: 0 | I_9 | AA_15 |

| | | | | | | | |
|---|---|---|---|---|---|---|---|
| 2482 | *SH3TC2* | nonsense | AR | Primary phenotype considered in current analysis:<br>Charcot-Marie-Tooth disease type<br><br>Other phenotypes associated with this variant:<br>Susceptibility to mononeuropathy of the median nerve, mild | TOPMed: 0.00060<br>gnomAD: 0.00065<br>1000G: –<br>Irish ALS: 0.0011<br>AnswerALS ALS: 0.0006<br>Irish Controls: 0<br>AnswerALS Controls: 0.0106383 | I_10 | AA_16 |
| 31166 | *POLR3B* | missense | AD | Primary phenotype considered in current analysis:<br>Charcot-Marie-Tooth disease, demyelinating<br><br>Other phenotypes associated with this variant:<br>Hypomyelinating leukodystrophy with or without oligodontia and-or hypogonadotropic hypogonadism; *POLR*-related leukodystrophy; *POLR3B*-related disorder | TOPMed: 0.00036<br>gnomAD: 0.00031<br>1000G: 0.00020<br>Irish ALS: 0.0011<br>AnswerALS ALS: 0.0006<br>Irish Controls: 0<br>AnswerALS Controls: 0 | I_11 | AA_17 |
| 929258 | *SORD* | frameshift | AR | Primary phenotype considered in current analysis:<br>Neuromuscular disease; Neuronopathy<br><br>Other phenotypes associated with this variant:<br>distal hereditary motor, autosomal recessive; Inborn genetic diseases | TOPMed: –<br>gnomAD v4: 0.003345 (European, non-Finnish)<br>1000G: –<br>Irish ALS: 0.0032<br>AnswerALS ALS: 0.00841<br>Irish Controls: 0.00217<br>AnswerALS Controls: 0 | I_12<br>I_13<br>I_14 | AA_18<br>AA_19<br>AA_20<br>AA_21<br>AA_22<br>AA_23<br>AA_24<br>AA_25<br>AA_26<br>AA_27<br>AA_28<br>AA_29<br>AA_30 |
| 13513 | *POLG* | missense | AR | Primary phenotype identified in current analysis: | TOPMed: 0.00094<br>gnomAD: 0.00093 | I_15<br>I_16 | AA_31<br>AA_32 |

| | | | | | | | |
|---|---|---|---|---|---|---|---|
| | | | | Hereditary Spastic Paraplegia; Sensory Ataxic Neuropathy<br><br>Other phenotypes associated with this variant:<br>*POLG*-related Disorder; Progressive Sclerosing Poliodystrophy; Progressive External Ophthalmoplegia with Mitochondrial Deletions; Mitochondrial DNA Depletion Syndrome; Mitochondrial Disease | 1000G: 0.00020<br>Irish ALS: 0.00213<br>AnswerALS ALS: 0.00323<br>Irish Controls: 0.00652<br>AnswerALS Controls: 0 | | AA_33<br>AA_34<br>AA_35 |
| 13496 | *POLG* | missense | AR | Primary phenotype identified in current analysis:<br>Hereditary Spastic Paraplegia; Spinocerebellar Ataxia (with Epilepsy); Sensory Ataxic Neuropathy<br><br>Other phenotypes associated with this variant:<br>Progressive external ophthalmoplegia with mitochondrial DNA deletions; Mitochondrial Disease; Progressive Sclerosing Poliodystrophy; Neurodevelopmental delay; *POLG*-Related Disorder | TOPMed: 0.00063<br>gnomAD: 0.00067<br>1000G: 0.00020<br>Irish ALS: 0.0011<br>AnswerALS ALS: 0.0006<br>Irish Controls: 0<br>AnswerALS Controls: 0 | I_17 | AA_36 |
| 3626 | *MPO* | Missense | AD | Primary phenotype identified in current analysis:<br>Alzheimer's Disease Type | TOPMed: 0.00178<br>gnomAD: 0.00173<br>1000G: 0.00040 | I_18<br>I_19<br>I_20 | AA_37<br>AA_38 |

| | | | | | | | |
|---|---|---|---|---|---|---|---|
| | | | | Other phenotypes associated with this variant:<br>Myeloperoxidase Deficiency | Irish ALS: 0.00320<br>AnswerALS ALS: 0.00129<br>Irish Controls: 0.002174<br>AnswerALS Controls: 0 | | |
| 206528 | *POLG* | Missense | AD and AR | Primary phenotype identified in current analysis:<br>Hereditary Spastic Paraplegia<br><br>Other phenotypes associated with this variant:<br>Progressive Sclerosing Poliodystrophy; Mitochondrial DNA Depletion Syndrome; Progressive External Ophthalmoplegia with Mitochondrial Deletions | TOPMed: 0.00008<br>gnomAD: 0.00007<br>1000G: –<br>Irish ALS: 00.0011<br>AnswerALS ALS: 0<br>Irish Controls: 0<br>AnswerALS Controls: 0 | I_21 | -- |
| 596302 | *SPG11* | frameshift | AR | Primary phenotype identified in current analysis:<br>Hereditary Spastic Paraplegia<br><br>Other phenotypes associated with this variant:<br>-- | TOPMed: 0.00003<br>gnomAD: 0.00003<br>1000G: –<br>Irish ALS: 0.0011<br>AnswerALS ALS: 0<br>Irish Controls: 0<br>AnswerALS Controls: 0 | I_22 | -- |
| 1705300 | *AMPD2* | Nonsense | AR | Primary phenotype identified in current analysis:<br>Hereditary Spastic Paraplegia<br><br>Other phenotypes associated with this variant: | TOPMed: --<br>GnomAD v4: 0.00003051 (European, non-Finnish)<br>1000G: –<br>Irish ALS: 0.0011 | I_23 | -- |

| | | | | | | | |
|---|---|---|---|---|---|---|---|
| | | | | Pontocerebellar Hypoplasia | AnswerALS ALS: 0<br>Irish Controls: 0<br>AnswerALS Controls: 0 | | |
| 217269 | *SPG7* | Missense | AR | Primary phenotype identified in current analysis:<br>Hereditary Spastic Paraplegia<br><br>Other phenotypes associated with this variant:<br>-- | TOPMed: 0.00028<br>gnomAD: 0.00015<br>1000G: 0.00020<br>Irish ALS: 0.0011<br>AnswerALS ALS: 0<br>Irish Controls: 0<br>AnswerALS Controls: 0 | I_24 | -- |
| 411675 | *SPG7* | frameshift | AR | Primary phenotype identified in current analysis:<br>Hereditary Spastic Paraplegia<br><br>Other phenotypes associated with this variant:<br>-- | TOPMed: –<br>gnomAD v3: 0.0002206 (European, non-Finnish)<br>1000G: –<br>Irish ALS: 0.0011<br>AnswerALS ALS: 0<br>Irish Controls: 0.0022<br>AnswerALS Controls: 0 | I_25 | -- |
| 6107 | *CYP7B1* | Missense and intron variant | AR | Primary phenotype identified in current analysis:<br>Hereditary Spastic Paraplegia<br><br>Other phenotypes associated with this variant:<br>-- | TOPMed: 0.00051<br>gnomAD: 0.00051<br>1000G: 0.00060<br>Irish ALS: 0.0011<br>AnswerALS ALS: 0<br>Irish Controls: 0.0022<br>AnswerALS Controls: 0 | I_26 | -- |
| 375313 | *AP5Z1* | Nonsense and non-coding transcript variant | AR | Primary phenotype identified in current analysis:<br>Hereditary Spastic Paraplegia<br><br>Other phenotypes associated with this variant:<br>-- | TOPMed: 0.00008<br>gnomAD: 0.00009<br>1000G: –<br>Irish ALS: 0.0011<br>AnswerALS ALS: 0<br>Irish Controls: 0<br>AnswerALS Controls: 0 | I_27 | -- |

| | | | | | | | |
|---|---|---|---|---|---|---|---|
| 234924 | *AP4S1* | nonsense | AR | Primary phenotype identified in current analysis:<br>Hereditary Spastic Paraplegia<br><br>Other phenotypes associated with this variant:<br>Intellectual Disability | TOPMed: 0.00011<br>gnomAD: 0.00009<br>1000G: 0.00020<br>Irish ALS: 0.0011<br>AnswerALS ALS: 0<br>Irish Controls: 0<br>AnswerALS Controls: 0 | I_28 | -- |
| 810060 | *AP5Z1* | Nonsense and non-coding transcript variant | AR | Primary phenotype identified in current analysis:<br>Hereditary Spastic Paraplegia<br><br>Other phenotypes associated with this variant:<br>Macular Dystrophy with or without Extraocular Features | TOPMed: 0.00007<br>gnomAD: 0.00006<br>1000G: –<br>Irish ALS: 0.0011<br>AnswerALS ALS: 0<br>Irish Controls: 0.0022<br>AnswerALS Controls: 0 | I_29 | -- |
| 450738 | *AP4M1* | frameshift | | Primary phenotype identified in current analysis:<br>Hereditary Spastic Paraplegia<br><br>Other phenotypes associated with this variant:<br>-- | TOPMed: –<br>gnomAD v4: 0.00003051 (European, non-Finnish)<br>1000G: –<br>Irish ALS: 0.0011<br>AnswerALS ALS: 0<br>Irish Controls: 0<br>AnswerALS Controls: 0 | I_30 | -- |
| 214335 | *FARS2* | missense | AR | Primary phenotype identified in current analysis:<br>Hereditary Spastic Paraplegia<br><br>Other phenotypes associated with this variant:<br>Combined Oxidative Phosphorylation Defect Type | TOPMed: 0.00012<br>gnomAD: 0.00011<br>1000G: –<br>Irish ALS: 0.0011<br>AnswerALS ALS: 0<br>Irish Controls: 0<br>AnswerALS Controls: 0 | I_31 | -- |

| | | | | | | | |
|---|---|---|---|---|---|---|---|
| 579997 | *FARS2* | frameshift | AR | Primary phenotype identified in current analysis:<br>Hereditary Spastic Paraplegia<br><br>Other phenotypes associated with this variant:<br>Combined Oxidative Phosphorylation Defect; Leigh Syndrome | TOPMed: 0.00006<br>gnomAD: 0.00011<br>1000G: –<br>Irish ALS: 0.0011<br>AnswerALS ALS: 0<br>Irish Controls: 0.0043<br>AnswerALS Controls: 0 | I_32 | -- |
| 6195 | *PLA2G6* | nonsense | AR | Primary phenotype identified in current analysis:<br>Autosomal recessive Parkinson Disease<br><br>Other phenotypes associated with this variant:<br>Infantile Neuroaxonal Dystrophy; *PLA2G6*-associated neurodegeneration; Neurodegeneration with brain iron accumulation | TOPMed: 0.00010<br>gnomAD: 0.00006<br>1000G: –<br>Irish ALS: 0.0011<br>AnswerALS ALS: 0<br>Irish Controls: 0<br>AnswerALS Controls: 0 | I_33 | -- |
| 372790 | *MFN2* | nonsense | AD and AR | Primary phenotype identified in current analysis:<br>Charcot-Marie-Tooth Disease;<br><br>Other phenotypes associated with this variant:<br>Hereditary motor and sensory neuropathy with optic atrophy | TOPMed: --<br>gnomAD v4: 0.00001271 (European, non-Finnish)<br>1000G: –<br>Irish ALS: 0.0011<br>AnswerALS ALS: 0<br>Irish Controls: 0<br>AnswerALS Controls: 0 | I_34 | -- |
| 246120 | *FIG4* | splice donor | AD and AR | Primary phenotype identified in current analysis:<br>Charcot-Marie-Tooth Disease | TOPMed: 0.00004<br>gnomAD: 0.00003<br>1000G: – | I_35<br>I_36 | -- |

| | | | | | | | |
|---|---|---|---|---|---|---|---|
| | | | | Other phenotypes associated with this variant:<br>-- | Irish ALS: 0.0021<br>AnswerALS ALS: 0<br>Irish Controls: 0.0043<br>AnswerALS Controls: 0 | | |
| 1426154 | *IGHMBP2* | missense | AR | Primary phenotype identified in current analysis:<br>Charcot-Marie-Tooth Disease Axonal<br><br>Other phenotypes associated with this variant:<br>Autosomal recessive distal spinal muscular atrophy | TOPMed: –<br>gnomAD v4: 0.00001104 (European, non-Finnish)<br>1000G: –<br>Irish ALS: 0.0011<br>AnswerALS ALS: 0<br>Irish Controls: 0<br>AnswerALS Controls: 0 | I_37 | -- |
| 2280 | *MFN2* | missense | AR | Primary phenotype identified in current analysis:<br>Charcot-Marie-Tooth Disease, axonal, autosomal recessive<br><br>Other phenotypes associated with this variant:<br>Peripheral axonal neuropathy; Neuropathy, hereditary motor and sensory | TOPMed: 0.00025<br>gnomAD: 0.00027<br>1000G: 0.00020<br>Irish ALS: 0.0011<br>AnswerALS ALS: 0<br>Irish Controls: 0<br>AnswerALS Controls: 0 | I_38 | -- |
| 689658 | *NUDT2* | frameshift | AR | Primary phenotype identified in current analysis:<br>Peripheral neuropathy<br><br>Other phenotypes associated with this variant: | TOPMed: –<br>gnomAD v4: 0.0003610 (European, non-Finnish)<br>1000G: –<br>Irish ALS: 0.0011<br>AnswerALS ALS: 0<br>Irish Controls: 0 | I_39 | -- |

| | | | | | | | |
|---|---|---|---|---|---|---|---|
| | | | | Intellectual developmental disorder with or without peripheral neuropathy; Complex neurodevelopmental disorder | AnswerALS Controls: 0 | | |
| 2136466 | *NEMF* | nonsense | AR | Primary phenotype identified in current analysis:<br>Axonal peripheral neuropathy<br><br>Other phenotypes associated with this variant:<br>Intellectual developmental disorder with speech delay | TOPMed: –<br>gnomAD: 0.00004<br>1000G: –<br>Irish ALS: 0.0011<br>AnswerALS ALS: 0<br>Irish Controls: 0<br>AnswerALS Controls: 0 | I_40 | -- |
| 162016 | *ANO10* | frameshift | AR | Primary phenotype identified in current analysis:<br>Autosomal recessive Spinocerebellar Ataxia<br><br>Other phenotypes associated with this variant:<br>-- | TOPMed: –<br>GnomAD v4: 0.0008157 (European, non-Finnish)<br>1000G: –<br>Irish ALS: 0.0043<br>AnswerALS ALS: 0<br>Irish Controls: 0.0022<br>AnswerALS Controls: 0 | I_41<br>I_42<br>I_43<br>I_44 | -- |
| 977147 | *MSTO1* | missense | AD and AR | Primary phenotype identified in current analysis:<br>Mitochondrial myopathy-cerebellar ataxia-pigmentary retinopathy syndrome<br><br>Other phenotypes associated with this variant:<br>-- | TOPMed: 0.00014<br>gnomAD: 0.00012<br>1000G: –<br>Irish ALS: 0.0011<br>AnswerALS ALS: 0<br>Irish Controls: 0<br>AnswerALS Controls: 0 | I_45 | -- |
| 3778778 | *RNF216* | splice donor | AR | Primary phenotype identified in current analysis:<br>Cerebellar ataxia-hypogonadism syndrome | TOPMed: 0.000132<br>gnomAD: 0.0001699<br>1000G: –<br>Irish ALS: 0.0011 | I_46 | -- |

| | | | | | | | |
|---|---|---|---|---|---|---|---|
| | | | | Other phenotypes associated with this variant:<br>-- | AnswerALS ALS: 0<br>Irish Controls: 0.0022<br>AnswerALS Controls: 0 | | |
| 846957 | *PRKN* | Frameshift and intron variant | AR | Primary phenotype identified in current analysis:<br>Autosomal recessive juvenile Parkinson disease<br><br>Other phenotypes associated with this variant:<br>Lung cancer; Ovarian Cancer | TOPMed: --<br>gnomAD: 0.00016<br>1000G: –<br>Irish ALS: 0.0011<br>AnswerALS ALS: 0<br>Irish Controls: 0<br>AnswerALS Controls: 0 | I_58 | -- |
| 1323796 | *AARS1* | Frameshift | AD | Primary phenotype identified in current analysis:<br>Charcot-Marie-Tooth Disease<br><br>Other phenotypes associated with this variant:<br>-- | TOPMed: –<br>gnomAD v4: 0.000002542 (European, non-Finnish)<br>1000G: –<br>Irish ALS: 0<br>AnswerALS ALS: 0.00065<br>Irish Controls: 0<br>AnserALS Controls: 0 | -- | AA_39 |
| 162195 | *IGHMBP2* | frameshift | AR | Primary phenotype identified in current analysis:<br>Charcot-Marie-Tooth Disease Axonal Type<br><br>Other phenotypes associated with this variant:<br>Distal Spinal Muscular Atrophy, autosomal recessive | TOPMed: –<br>GnomAD v4: 0.0001848 (European, non-Finnish)<br>1000G: –<br>Irish ALS: 0<br>AnswerALS ALS: 0.00065<br>Irish Controls: 0 | -- | AA_44 |

| | | | | | | | |
|---|---|---|---|---|---|---|---|
| | | | | | AnwerALS Controls: 0 | | |
| 1908695 | *PRX* | Frameshift and 3 prime UTR | AR | Primary phenotype identified in current analysis:<br>Charcot-Marie-Tooth Disease<br><br>Other phenotypes associated with this variant:<br>-- | TOPMed: –<br>gnomAD v3: 0.00001315 (Total population)<br>1000G: –<br>Irish ALS: 0<br>AnswerALS ALS: 0.00065<br>Irish Controls: 0<br>AnswerALS Controls: 0 | -- | AA_45 |
| 234316 | *IGHMBP2* | nonsense | AR | Primary phenotype identified in current analysis:<br>Charcot-Marie-Tooth Disease Axonal Type<br><br>Other phenotypes associated with this variant:<br>Autosomal recessive Distal Spinal Muscular Atrophy; Neurodevelopmental Disorder | TOPMed:0.00015<br>gnomAD:0.00016<br>1000G: –<br>Irish ALS: 0<br>AnswerALS ALS: 0.00065<br>Irish Controls: 0<br>AnswerALS Controls: 0 | -- | AA_46 |
| 566096 | *PLEKHG5* | nonsense | AR | Primary phenotype identified in current analysis:<br>Charcot-Marie-Tooth Disease Recessive Intermediate<br><br>Other phenotypes associated with this variant:<br>Distal Hereditary Motor Neuronopathy | TOPMed:0.00003<br>gnomAD:0.00006<br>1000G: –<br>Irish ALS: 0<br>AnswerALS ALS: 0.00065<br>Irish Controls: 0.0022<br>AnswerALS Controls: 0 | -- | AA_20 |
| 574671 | *PRX* | nonsense | AR | Primary phenotype identified in current analysis:<br>Charcot-Marie-Tooth Disease | TOPMed: 0.00002<br>gnomAD: 0.000009322 (European, non-Finnish) | -- | AA_91 |

| | | | | | | | |
|---|---|---|---|---|---|---|---|
| | | | | Other phenotypes associated with this variant:<br>-- | 1000G: –<br>Irish ALS: 0<br>AnswerALS ALS: 0.00065<br>Irish Controls: 0<br>AnswerALS Controls: 0 | | |
| 13502 | *POLG* | missense | AR | Primary phenotype identified in current analysis:<br>Hereditary Spastic Paraplegia<br><br>Other phenotypes associated with this variant:<br>Progressive External Ophthalmoplegia with Mitochondrial DNA Deletion, Digenic; Mitochondrial DNA Depletion Syndrome; Progressive Sclerosing Poliodystrophy; Sensory Ataxic Neuropathy; POLG-Related Spectrum Disorders; Mitochondrial Disease | TOPMed: 0.00018<br>gnomAD: 0.00028<br>1000G: 0.00020<br>Irish ALS: 0<br>AnswerALS ALS: 0.0026<br>Irish Controls: 0<br>AnswerALS Controls: 0 | -- | AA_47<br>AA_48<br>AA_49<br>AA_50 |
| 1437494 | *AP4M1* | frameshift | AR | Primary phenotype identified in current analysis:<br>Hereditary Spastic Paraplegia<br><br>Other phenotypes associated with this variant:<br>-- | TOPMed:0.00004<br>gnomAD:0.00003<br>1000G: –<br>Irish ALS: 0<br>AnswerALS ALS: 0.00065<br>Irish Controls: 0<br>AnswerALS Controls: 0 | -- | AA_51 |
| 156414 | *AP4B1* | frameshift | AR | Primary phenotype identified in current analysis:<br>Hereditary Spastic Paraplegia | TOPMed: –<br>GnomAD v4: 0.0001280 (European, non-Finnish) | -- | AA_52 |

| | | | | | | | |
|---|---|---|---|---|---|---|---|
| | | | | Other phenotypes associated with this variant:<br>Intellectual Disability; Abnormality of the nervous system | 1000G: –<br>Irish ALS: 0<br>AnswerALS ALS: 0.00065<br>Irish Controls: 0<br>AnswerALS Controls: 0 | | |
| 214192 | *MTRFR* | frameshift | AR | Primary phenotype identified in current analysis:<br>Hereditary Spastic Paraplegia<br><br>Other phenotypes associated with this variant:<br>Combined oxidative phosphorylation defect type | TOPMed:0.00005<br>gnomAD:0.00007<br>1000G: –<br>Irish ALS: 0<br>AnswerALS ALS: 0.00065<br>Irish Controls: 0.0022<br>AnswerALS Controls: 0 | -- | AA_53 |
| 219448 | *SPG7* | missense | AR | Primary phenotype identified in current analysis:<br>Hereditary Spastic Paraplegia<br><br>Other phenotypes associated with this variant:<br>-- | TOPMed: –<br>gnomAD: 0.00001<br>1000G: –<br>Irish ALS: 0<br>AnswerALS ALS: 0.00065<br>Irish Controls: 0<br>AnswerALS Controls: 0 | -- | AA_54 |
| 2906297 | *PNPLA6* | nonsense | AR | Primary phenotype identified in current analysis:<br>Hereditary Spastic Paraplegia<br><br>Other phenotypes associated with this variant:<br>-- | TOPMed: –<br>gnomAD:0.00002<br>1000G: –<br>Irish ALS: 0<br>AnswerALS ALS: 0.00065<br>Irish Controls: 0<br>AnswerALS Controls: 0 | -- | AA_55 |
| 422933 | *DDHD1* | frameshift | AR | Primary phenotype identified in current analysis:<br>Hereditary Spastic Paraplegia | TOPMed:0.00002<br>gnomAD:0.00003<br>1000G: –<br>Irish ALS: 0 | -- | AA_56 |

| | | | | | | | |
|---|---|---|---|---|---|---|---|
| | | | | Other phenotypes associated with this variant:<br>-- | AnswerALS ALS: 0.00065<br>Irish Controls: 0<br>AnswerALS Controls: 0 | | |
| 465174 | *SPG7* | splice acceptor | AR | Primary phenotype identified in current analysis:<br>Hereditary Spastic Paraplegia<br><br>Other phenotypes associated with this variant:<br>-- | TOPMed:0.00000<br>gnomAD:0.00001<br>1000G: 0.00040<br>Irish ALS: 0<br>AnswerALS ALS: 0.00065<br>Irish Controls: 0<br>AnswerALS Controls: 0 | -- | AA_57 |
| 6816 | *SPG7* | nonsense | AR | Primary phenotype identified in current analysis:<br>Hereditary Spastic Paraplegia<br><br>Other phenotypes associated with this variant:<br>Proximal Spinal Muscular Atrophy; Retinal Dystrophy; *SPG7*-Related Disorder | TOPMed:0.00010<br>gnomAD:0.00014<br>1000G: –<br>Irish ALS: 0<br>AnswerALS ALS: 0.00065<br>Irish Controls: 0<br>AnswerALS Controls: 0 | -- | AA_58 |
| 6819 | *SPG7* | missense | AR | Primary phenotype identified in current analysis:<br>Hereditary Spastic Paraplegia<br><br>Other phenotypes associated with this variant:<br>Retinal Dystrophy; *SPG7*-Related Disorder | TOPMed:0.00107<br>gnomAD:0.00102<br>1000G: 0.00080<br>Irish ALS: 0<br>AnswerALS ALS: 0.00194<br>Irish Controls: 0<br>AnswerALS Controls: 0 | -- | AA_59<br>AA_60<br>AA_61 |
| 930104 | *FA2H* | missense | AR | Primary phenotype identified in current analysis:<br>Hereditary Spastic Paraplegia | TOPMed:0.00006<br>gnomAD:0.00004<br>1000G: –<br>Irish ALS: 0 | -- | AA_62 |

| | | | | | | | |
|---|---|---|---|---|---|---|---|
| | | | | Other phenotypes associated with this variant:<br>-- | AnswerALS ALS: 0.00065<br>Irish Controls: 0<br>AnswerALS Controls: 0 | | |
| 13507 | *POLG* | missense | AR | Primary phenotype identified in current analysis:<br>Spinocerebellar Ataxia with Epilepsy<br><br>Other phenotypes associated with this variant:<br>Mitochondrial DNA Depletion Syndrome; Progressive External Ophthalmoplegia with Mitochondrial Deletions, Autosomal Dominant; *POLG*-Related Spectrum Disorders; Progressive Sclerosing Poliodystrophy; Sensory and Ataxic Neuropathy; Mitochondrial Disease | TOPMed:0.00031<br>gnomAD:0.00081<br>1000G: –<br>Irish ALS: 0<br>AnswerALS ALS: 0.00065<br>Irish Controls: 0<br>AnswerALS Controls: 0 | -- | AA_63 |
| 2443782 | *RNU12* | non-coding transcript variant | AR | Primary phenotype identified in current analysis:<br>Spinocerebellar Ataxia, Autosomal Recessive<br><br>Other phenotypes associated with this variant:<br>-- | TOPMed: --<br>GnomAD v4: 0.00006297 (European, non-Finnish<br>1000G: –<br>Irish ALS: 0<br>AnswerALS ALS: 0.00065<br>Irish Controls: 0<br>AnswerALS Controls: 0 | -- | AA_64 |
| 1940 | *LRRK2* | missense | AD | Primary phenotype identified in current analysis: | TOPMed:0.00059<br>gnomAD:0.00036<br>1000G: 0.00020<br>Irish ALS: 0 | -- | AA_65 |

| | | | | | | | |
|---|---|---|---|---|---|---|---|
| | | | | Young Onset Parkinson Disease; Parkinson Disease, autosomal dominant; Late-onset Parkinson Disease<br><br>Other phenotypes associated with this variant:<br>-- | AnswerALS ALS: 0.00065<br>Irish Controls: 0<br>AnswerALS Controls: 0 | | |
| 857581 | *PSAP* | missense | AD | Primary phenotype identified in current analysis:<br>Parkinson Disease, autosomal dominant<br><br>Other phenotypes associated with this variant:<br>Sphingolipid activator protein deficiency; Combined PSAP deficiency; Krabbe Disease due to Saposin A deficiency; Gaucher Disease due to Saposin C deficiency; Metachromatic Leukodystrophy | TOPMed: –<br>gnomAD:0.00002<br>1000G: –<br>Irish ALS: 0<br>AnswerALS ALS: 0.00065<br>Irish Controls: 0<br>AnswerALS Controls: 0 | -- | AA_66 |
| 870544 | *VPS13* | nonsense | AR | Primary phenotype identified in current analysis:<br>Autosomal recessive early-onset Parkinson Disease<br><br>Other phenotypes associated with this variant:<br>-- | TOPMed:0.00023<br>gnomAD:0.00017<br>1000G: –<br>Irish ALS: 0<br>AnswerALS ALS: 0.00129<br>Irish Controls: 0<br>AnswerALS Controls: 0 | -- | AA_26<br>AA_67 |
| 196723 | *TTN* | splice donor | AR | Primary phenotype identified in current analysis:<br>Neuromuscular Disease | TOPMed:0.00002<br>gnomAD:0.00002<br>1000G: 0.00020<br>Irish ALS: 0 | -- | AA_68 |

| | | | | | | | |
|---|---|---|---|---|---|---|---|
| | | | | Other phenotypes associated with this variant:<br>Primary Dilated Cardiomyopathy; Cardiovascular Phenotype; Myopathy with early respiratory failure; Tibial muscular dystrophy; Autosomal recessive limb-girdle muscular dystrophy Type; Early onset myopathy with fatal cardiomyopathy | AnswerALS ALS: 0.00065<br>Irish Controls: 0<br>AnswerALS Controls: 0 | | |
| 3376522 | *RFC4* | splice donor | AR | Primary phenotype identified in current analysis:<br>Morimoto-Ryu-Malicdan Neuromuscular Syndrome<br><br>Other phenotypes associated with this variant:<br>-- | TOPMed: –<br>gnomAD: –<br>1000G: –<br>Irish ALS: 0<br>AnswerALS ALS: 0.00065<br>Irish Controls: 0<br>AnswerALS Controls: 0 | -- | AA_48 |
| 830327 | *VWA1* | frameshift | AR | Primary phenotype identified in current analysis:<br>Neuromuscular Disease<br><br>Other phenotypes associated with this variant:<br>*VWA1*-Related Disorder; Distal Hereditary Motor Neuronopathy, autosomal recessive | TOPMed: –<br>gnomAD v4: 0.00099906 (European, non-Finnish)<br>1000G: –<br>Irish ALS: 0<br>AnswerALS ALS: 0.00065<br>Irish Controls: 0<br>AnswerALS Controls: 0 | -- | AA_69 |
| 391712 | *DST* | Intron variant and nonsense | AR | Primary phenotype identified in current analysis:<br>Hereditary sensory and autonomic neuropathy | TOPMed:0.00000<br>gnomAD:0.00001<br>1000G: –<br>Irish ALS: 0 | -- | AA_70 |

| | | | | | | | |
|---|---|---|---|---|---|---|---|
| | | | | Other phenotypes associated with this variant:<br>Epidermolysis bullosa simplex | AnswerALS ALS: 0.00065<br>Irish Controls: 0<br>AnswerALS Controls: 0 | | |
| 847629 | *DST* | Intron variant and nonsense | AR | Primary phenotype identified in current analysis:<br>Hereditary sensory and autonomic neuropathy<br><br>Other phenotypes associated with this variant:<br>Epidermolysis bullosa simplex | TOPMed:0.00004<br>gnomAD:0.00003<br>1000G: –<br>Irish ALS: 0<br>AnswerALS ALS: 0.00065<br>Irish Controls: 0<br>AnswerALS Controls: 0 | -- | AA_71 |
| 976691 | *DNAJC30* | missense | AR | Primary phenotype identified in current analysis:<br>Leber-hereditary optic neuropathy, autosomal recessive<br><br>Other phenotypes associated with this variant:<br>Leber Optic Atrophy | TOPMed:0.00065<br>gnomAD:0.00111<br>1000G: 0.00016<br>Irish ALS: 0<br>AnswerALS ALS: 0.00065<br>Irish Controls: 0<br>AnswerALS Controls: 0 | -- | AA_72 |

**Supplemental Table S3.2.** Neurologic associated ClinVar variants identified in Irish ALS (n=47) and AnswerALS (n=69) pwALS. Anonymised IDs, ClinVar Variant IDs, associated genes, ALS clinical phenotype including neuropsychological testing, and family history of neurological or psychiatric disease are displayed per variant carrier. General variant categories that appeared from annotation include Parkinsonian, Charcot-Marie-Tooth Disease, Neuromuscular Disease, Hereditary Spastic Paraplegia, Ataxia, and Alzheimer's Disease. Neuropsychological testing for Irish cohort included Beaumont Behavioural Inventory (BBI) and the Edinburgh Cognitive and Behavioural ALS Screen (ECAS), while cognitive assessment in the AnswerALS cohort utilised the ALS Cognitive and Behavioural Screen (CBS). Any abnormal cognitive/behavioural result recorded at any timepoint during disease course is indicated in the phenotype column. For CBS classification. ALSci, ALS with cognitive impairment; ALSbi, ALS with behavioural impairment.

| Case ID | ClinVar Variant ID | Gene | Phenotype | Family History |
|---|---|---|---|---|
| Parkinsonian Variants | | | | |
| I_1 | 4290 | *GBA1* | Diagnosis: ALS<br>El Escorial: Probable<br>Age of Onset: 50-54 years<br>Survival (months, Sx onset to death): 41.29<br>Site of Onset: Bulbar<br>BBI: Normal<br>ECAS: Abnormal | 2 x third-degree ALS<br>Third-degree Parkinson disease |
| AA_1 | 4290 | *GBA1* | Diagnosis: ALS<br>El Escorial: Probable<br>Age of Onset: Not provided<br>Survival (months, Sx onset to death): Not available<br>Site of Onset: Spinal<br>CBS: Normal cognition, behaviour not performed | Not provided |
| AA_2 | 4290 | *GBA1* | Diagnosis: ALS<br>El Escorial: Possible<br>Age of Onset: 40-44 years<br>Survival (months, Sx onset to death): Not available<br>Site of Onset: Bulbar<br>CBS: Cognition and behaviour normal | Second-degree Alzheimer's dementia |
| AA_3 | 4290 | *GBA1* | Diagnosis: ALS<br>El Escorial: Probable<br>Age of Onset: 60-64 years<br>Survival (months, Sx onset to death): 20.93<br>Site of Onset: Bulbar | 2 x fourth-degree ALS<br>2 x second-degree ALS |

| | | | | |
|---|---|---|---|---|
| | | | CBS: ALSci, behaviour not performed | |
| AA_4 | 4290 | *GBA1* | Diagnosis: ALS<br>El Escorial: Definite<br>Age of Onset: 50-54 years<br>Survival (months, Sx onset to death): 99.64<br>Site of Onset: Bulbar<br>CBS: Cognition and behaviour normal | None |
| AA_5 | 4290 | *GBA1* | Diagnosis: ALS<br>El Escorial: Probable<br>Age of Onset: 50-54 years<br>Survival (months, Sx onset to death): Not available<br>Site of Onset: Bulbar<br>CBS: Cognition and behaviour normal | First-degree Alzheimer’s disease |
| AA_6 | 4290 | *GBA1* | Diagnosis: ALS<br>El Escorial: Probable<br>Age of Onset: 55-59 years<br>Survival (months, Sx onset to death): Not available<br>Site of Onset: Spinal<br>CBS: Not performed | First-degree Parkinson disease |
| AA_7 | 4290 | *GBA1* | Diagnosis: ALS<br>El Escorial: not provided<br>Age of Onset: 60-64 years<br>Survival (months, Sx onset to death): Not available<br>Site of Onset: Spinal<br>CBS: ALSci, normal behaviour | Second-degree dementia |
| I_2 | 7050 | *PRKN* | Diagnosis: ALS<br>El Escorial: Possible<br>Age of Onset: 60-64 years<br>Survival (months, Sx onset to death): 31.87<br>Site of Onset: Thoracic/Respiratory<br>BBI: mild changes<br>ECAS: Abnormal | Second-degree Alzheimer’s disease<br>First-degree alcoholism |
| I_3 | 7050 | *PRKN* | Diagnosis: ALS<br>El Escorial: Definite<br>Age of Onset: 80-84 years<br>Survival (months, Sx onset to death): 40-44 years | First-degree alcoholism<br>First-degree Parkinson disease<br>Third-degree multiple sclerosis |

| | | | | |
|---|---|---|---|---|
| | | | Site of Onset: Spinal<br>BBI: mild changes<br>ECAS: Abnormal | |
| I_4 | 7050 | *PRKN* | Diagnosis: ALS<br>El Escorial: Definite<br>Age of Onset: 65-69 years<br>Survival (months, Sx onset to death): 26.61<br>Site of Onset: Spinal<br>BBI: not performed<br>ECAS: Normal | Second-degree Alzheimer's Disease<br>Third-degree suspected ALS |
| I_5 | 7050 | *PRKN* | Diagnosis: ALS<br>El Escorial: Definite<br>Age of Onset: 60-64 years<br>Survival (months, Sx onset to death): 20.07<br>Site of Onset: Spinal<br>BBI: Normal<br>ECAS: Abnormal | First-degree alcoholism |
| I_6 | 7050 | *PRKN* | Diagnosis: ALS<br>El Escorial: Suspected<br>Age of Onset: 60-64 years<br>Survival (months, Sx onset to death): 84.03<br>Site of Onset: Thoracic/Respiratory<br>BBI: Normal<br>ECAS: Normal | Fist-degree ALS<br>2 x first-degree FTD |
| I_58 | 846957 | *PRKN* | Diagnosis: ALS<br>El Escorial: Possible<br>Age of Onset: 65-69 years<br>Survival (months, Sx onset to death): 118.27<br>Site of Onset: Spinal<br>BBI: Mild Changes<br>ECAS: Abnormal | Second-degree Alzheimer's disease |
| AA_8 | 7050 | *PRKN* | Diagnosis: ALS<br>El Escorial: Probable<br>Age of Onset: 50-54 years<br>Survival (months, Sx onset to death): Not available<br>Site of Onset: Spinal | First-degree dementia |

| | | | | |
|---|---|---|---|---|
| | | | CBS: ALSci behaviour indicates possible FTD | |
| AA_9 | 7050 | *PRKN* | Diagnosis: ALS<br>El Escorial: Suspected<br>Age of Onset: 35-39 years<br>Survival (months, Sx onset to death): Not available<br>Site of Onset: Spinal<br>CBS: ALSci, behaviour not performed | Not provided |
| AA_10 | 7050 | *PRKN* | Diagnosis: ALS<br>El Escorial: Possible<br>Age of Onset: 65-69 years<br>Survival (months, Sx onset to death): Not available<br>Site of Onset: Spinal<br>CBS: ALSci, behaviour normal | First-degree bipolar disorder<br>First-degree dementia |
| AA_11 | 7050 | *PRKN* | Diagnosis: ALS<br>El Escorial: Probable<br>Age of Onset: 50-54 years<br>Survival (months, Sx onset to death): Not available<br>Site of Onset: Spinal<br>CBS: ALSci, behaviour normal | None |
| AA_12 | 7050 | *PRKN* | Diagnosis: ALS<br>El Escorial: Probable<br>Age of Onset: 55-59 years<br>Survival (months, Sx onset to death): 37.55<br>Site of Onset: Spinal<br>CBS: Cognition and behaviour normal | First-degree dementia and Parkinson Disease |
| AA_13 | 7050 | *PRKN* | Diagnosis: ALS<br>El Escorial: Probable<br>Age of Onset: 75-79 years<br>Survival (months, Sx onset to death) Not available<br>Site of Onset: Spinal<br>CBS: ALSci and ALSbi | None |
| I_7 | 641981 | *FBXO7* | Diagnosis: ALS<br>El Escorial: Definite<br>Age of Onset: 60-64 years<br>Survival (months, Sx onset to death): 25.1<br>Site of Onset: Spinal | First-degree multiple sclerosis<br>First-degree alcoholism<br>Second-degree dementia |

| | | | | |
|---|---|---|---|---|
| | | | BBI: Not performed<br>ECAS: Not performed | |
| I_8 | 641981 | *FBXO7* | Diagnosis: ALS<br>El Escorial: Probable<br>Age of Onset: 45-49 years<br>Survival (months, Sx onset to death): 29.89<br>Site of Onset: Spinal<br>BBI: Mild changes<br>ECAS: Normal | 2 x third-degree ALS |
| AA_14 | 641981 | *FBXO7* | Diagnosis: ALS<br>El Escorial: Definite<br>Age of Onset: 45-49 years<br>Survival (months, Sx onset to death): Not available<br>Site of Onset: Spinal<br>CBS: Cognition normal, behaviour not performed | First-degree epilepsy<br>Third-degree dementia |
| I_33 | 6195 | *PLA2G6* | Diagnosis: ALS<br>El Escorial: Probable<br>Age of Onset: 55-59 years<br>Survival (months, Sx onset to death): 104.47<br>Site of Onset: Spinal<br>BBI: Normal<br>ECAS: Normal | Third-degree multiple sclerosis<br>Second-degree autism |
| AA_65 | 1940 | *LRRK2* | Diagnosis: ALS<br>El Escorial: Probable<br>Age of Onset: 80-84 years<br>Survival (months, Sx onset to death): 15.05<br>Site of Onset: Spinal<br>CBS: Behaviour not performed, Cognition normal | None |
| AA_66 | 857581 | *PSAP* | Diagnosis: ALS<br>El Escorial: Probable<br>Age of Onset: 55-59 years<br>Survival (months, Sx onset to death): 34.59<br>Site of Onset: Spinal<br>CBS: Behaviour normal, ALSci | None |
| AA_26 | 870544 | *VPS13C* | Diagnosis: ALS<br>El Escorial: Definite | Fourth-degree ALS |

| | | | | |
|---|---|---|---|---|
| | | | Age of Onset: 35-39 years<br>Survival (months, Sx onset to death): Not available<br>Site of Onset: Spinal<br>CBS: Behaviour indicates possible FTD, ALSci | |
| AA_67 | 870544 | *VPS13C* | Diagnosis: ALS<br>El Escorial: Probable<br>Age of Onset: Not available<br>Survival (months, Sx onset to death): Not available<br>Site of Onset: Bulbar and Spinal<br>CBS: ALSbi, ALSci | First-degree Parkinson disease |
| Charcot-Marie Tooth Disease Variants | | | | |
| I_9 | 444787 | *SURF1* | Diagnosis: ALS<br>El Escorial: Not performed<br>Age of Onset: 80-84 years<br>Survival (months, Sx onset to death): 29.89<br>Site of Onset: Spinal<br>BBI: Not performed<br>ECAS: Abnormal | Second-degree dementia<br>Third-degree Parkinson disease<br>Second-degree suicide<br>Third-degree autism<br>4 x First-degree alcoholism<br>First-degree other neuropsychiatric |
| AA_15 | 444787 | *SURF1* | Diagnosis: ALS<br>El Escorial: Definite<br>Age of Onset: 55-59 years<br>Survival (months, Sx onset to death): 107.19<br>Site of Onset: Spinal<br>CBS: ALSci, behaviour possible FTD | First-degree dementia<br>Second-degree Alzheimer's disease |
| I_10 | 2482 | *SH3TC2* | Diagnosis: ALS<br>El Escorial: Possible<br>Age of Onset: 70-74 years<br>Survival (months, Sx onset to death): 26.45<br>Site of Onset: Spinal<br>BBI: normal<br>ECAS: normal | Not performed |
| AA_16 | 2482 | *SH3TC2* | Diagnosis: ALS<br>El Escorial: Possible<br>Age of Onset: 40-44 years<br>Survival (months, Sx onset to death): Not available<br>Site of Onset: Spinal | None |

| | | | | |
|---|---|---|---|---|
| | | | CBS: Cognition and behaviour normal | |
| I_11 | 31166 | *POLR3B* | Diagnosis: ALS<br>El Escorial: Definite<br>Age of Onset: 60-64 years<br>Survival (months, Sx onset to death): 87.16<br>Site of Onset: Thoracic/Respiratory<br>BBI: Mild changes<br>ECAS: Normal | Not provided |
| AA_17 | 31166 | *POLR3B* | Diagnosis: ALS<br>El Escorial: Suspected<br>Age of Onset: 45-49 years<br>Survival (months, Sx onset to death): 28.42<br>Site of Onset: Spinal<br>CBS: Cognition normal, behaviour not performed | Second-degree Alzheimer's disease |
| I_34 | 372790 | *MFN2* | Diagnosis: ALS<br>El Escorial: Possible<br>Age of Onset: 85-89 years<br>Survival (months, Sx onset to death): 35.38<br>Site of Onset: Spinal<br>BBI: Normal<br>ECAS: Abnormal | Not provided |
| I_35 | 246120 | *FIG4* | Diagnosis: ALS<br>El Escorial: Probable<br>Age of Onset: 70-74 years<br>Survival (months, Sx onset to death): 19.97<br>Site of Onset: Spinal<br>BBI: None<br>ECAS: Normal | First-degree ALS<br>First-degree alcoholism and FTD<br>Second-degree dementia<br>First-degree alcoholism<br>2 x first-degree other neuropsychiatric<br>Second-degree suicide<br>Second-degree autism |
| I_36 | 246120 | *FIG4* | Diagnosis: ALS<br>El Escorial: Definite<br>Age of Onset: 45-49 years<br>Survival (months, Sx onset to death): 62.16<br>Site of Onset: Spinal<br>BBI: Mild changes<br>ECAS: Normal | Not provided |

| | | | | |
|---|---|---|---|---|
| I_37 | 1426154 | *IGHMBP2* | Diagnosis: ALS<br>El Escorial: Definite<br>Age of Onset: 65-69 years<br>Survival (months, Sx onset to death): 12.94<br>Site of Onset: Bulbar<br>BBI: None<br>ECAS: None | First-degree alcoholism and depression<br>2 x first-degree alcoholism<br>Second-degree alcoholism |
| I_38 | 2280 | *MFN2* | Diagnosis: ALS<br>El Escorial: Possible<br>Age of Onset: 50-54 years<br>Survival (months, Sx onset to death): 134.43<br>Site of Onset: Thoracic/Respiratory<br>BBI: Normal<br>ECAS: Normal | First-degree alcoholism |
| AA_39 | 1323796 | *AARS1* | Diagnosis: ALS<br>El Escorial: Probable<br>Age of Onset: 55-59 years<br>Survival (months, Sx onset to death): Not available<br>Site of Onset: Spinal<br>CBS: ALSbi, ALSci | Second-degree dementia<br>First-degree Alzheimer's dementia and Parkinson disease<br>First-degree Parkinson's Disease |
| AA_44 | 162195 | *IGHMBP2* | Diagnosis: ALS<br>El Escorial: Definite<br>Age of Onset: 60-64 years<br>Survival (months, Sx onset to death): 39.88<br>Site of Onset: Spinal<br>CBS: Behaviour normal, Cognition normal | First-degree ALS |
| AA_45 | 1908695 | *PRX* | Diagnosis: ALS<br>El Escorial: Definite<br>Age of Onset: 55-59 years<br>Survival (months, Sx onset to death): Not available<br>Site of Onset: Spinal<br>CBS: Behaviour normal, Cognition normal | None |
| AA_46 | 234316 | *IGHMBP2* | Diagnosis: ALS<br>El Escorial: Possible<br>Age of Onset: 50-54 years<br>Survival (months, Sx onset to death): Not available | Third-degree dementia<br>First-degree Alzheimer's disease and Parkinson disease |

| | | | | |
|---|---|---|---|---|
| | | | Site of Onset: Spinal<br>CBS: Behaviour normal, Cognition normal | |
| AA_20 | 566096 | *PLEKHG5* | Diagnosis: ALS<br>El Escorial: Probable<br>Age of Onset: 55-59 years<br>Survival (months, Sx onset to death): 80.85<br>Site of Onset: Bulbar<br>CBS: ALSbi, ALSci | None |
| AA_91 | 574671 | *PRX* | Diagnosis: ALS<br>El Escorial: Probable<br>Age of Onset: 60-64 years<br>Survival (months, Sx onset to death): 11.86<br>Site of Onset: Bulbar<br>CBS: ALSbi, Cognition indicates possible FTD | First-degree Alzheimer's disease |
| Neuromuscular Disease Variants | | | | |
| I_12 | 929258 | *SORD* | Diagnosis: ALS<br>El Escorial: Definite<br>Age of Onset: 50-54 years<br>Survival (months, Sx onset to death): 36.3<br>Site of Onset: Spinal<br>BBI: Normal<br>ECAS: Normal | Third-degree ALS<br>First-degree Alzheimer's disease<br>Second-degree Alzheimer's Disease<br>Third-degree autism<br>Third-degree other neurological<br>Third-degree suicide<br>Third-degree suicide<br>Third-degree ALSFTD |
| I_13 | 929258 | *SORD* | Diagnosis: ALS<br>El Escorial: not listed<br>Age of Onset: 75-79 years<br>Survival (months, Sx onset to death): 100.2<br>Site of Onset: Spinal (UMN predominant)<br>BBI: Not performed<br>ECAS: Not performed | Second-degree dementia |
| I_14 | 929258 | *SORD* | Diagnosis: ALS<br>El Escorial: Definite<br>Age of Onset: 60-64 years<br>Survival (months, Sx onset to death): 42.61<br>Site of Onset: Thoracic/Respiratory<br>BBI: Normal | Second-degree Parkinson disease<br>First-degree dementia<br>2 x second-degree with dementia<br>Second-degree bipolar disorder |

| | | | | |
|---|---|---|---|---|
| | | | ECAS: Not performed | |
| AA_18 | 929258 | *SORD* | Diagnosis: ALS<br>El Escorial: Definite<br>Age of Onset: 60-64 years<br>Survival (months, Sx onset to death): 28.19<br>Site of Onset: Spinal<br>CBS: Cognition and behaviour normal | 2 x first-degree Alzheimer's disease<br>2 x first-degree ALS |
| AA_19 | 929258 | *SORD* | Diagnosis: ALS<br>El Escorial: Probable<br>Age of Onset: 35-39 years<br>Survival (months, Sx onset to death): 86.07<br>Site of Onset: Spinal<br>CBS: ALSci and behaviour normal | None |
| AA_20 | 929258 | *SORD* | Diagnosis: ALS<br>El Escorial: Probable<br>Age of Onset: 55-59 years<br>Survival (months, Sx onset to death): 80.85<br>Site of Onset: Bulbar<br>CBS: ALSci and ALSbi | None |
| AA_21 | 929258 | *SORD* | Diagnosis: ALS<br>El Escorial: Definite<br>Age of Onset: 45-49 years<br>Survival (months, Sx onset to death): 65.34<br>Site of Onset: Spinal<br>CBS: ALSci and behaviour normal | None |
| AA_22 | 929258 | *SORD* | Diagnosis: ALS<br>El Escorial: Probable<br>Age of Onset: 45-49 years<br>Survival (months, Sx onset to death): Not available<br>Site of Onset: Spinal<br>CBS: Cognition normal, behaviour normal | None |
| AA_23 | 929258 | *SORD* | Diagnosis: ALS<br>El Escorial: Probable<br>Age of Onset: 30-34 years<br>Survival (months, Sx onset to death): Not available<br>Site of Onset: Spinal | None |

| | | | | |
|---|---|---|---|---|
| | | | CBS: Cognition normal, behaviour not performed | |
| AA_24 | 929258 | *SORD* | Diagnosis: ALS<br>El Escorial: Possible<br>Age of Onset: 65-69 years<br>Survival (months, Sx onset to death): 52.56<br>Site of Onset: Spinal<br>CBS: ALSci, behaviour normal | None |
| AA_25 | 929258 | *SORD* | Diagnosis: ALS<br>El Escorial: Probable<br>Age of Onset: 70-74 years<br>Survival (months, Sx onset to death): 31.44<br>Site of Onset: Spinal<br>CBS: Not performed | None |
| AA_26 | 929258 | *SORD* | Diagnosis: ALS<br>El Escorial: Definite<br>Age of Onset: 35-39 years<br>Survival (months, Sx onset to death): Not available<br>Site of Onset: Spinal<br>CBS: ALSci behaviour indicates possible FTD | Fourth-degree Alzheimer's disease |
| AA_27 | 929258 | *SORD* | Diagnosis: ALS<br>El Escorial: Suspected<br>Age of Onset: 85-89 years<br>Survival (months, Sx onset to death): Not available<br>Site of Onset: Bulbar<br>CBS: Cognition not performed ALSBi | First-degree dementia |
| AA_28 | 929258 | *SORD* | Diagnosis: ALS<br>El Escorial: Probable<br>Age of Onset: 45-49 years<br>Survival (months, Sx onset to death): Not available<br>Site of Onset: Spinal<br>CBS: Cognition and behaviour normal | Third-degree ALS |
| AA_29 | 929258 | *SORD* | Diagnosis: ALS<br>El Escorial: Definite<br>Age of Onset: 45-49 years<br>Survival (months, Sx onset to death): Not available<br>Site of Onset: Bulbar | Second-degree Alzheimer's disease<br>Second-degree Alzheimer's disease |

| | | | | |
|---|---|---|---|---|
| | | | CBS: Cognition normal behaviour not performed | |
| AA_30 | 929258 | *SORD* | Diagnosis: ALS<br>El Escorial: Definite<br>Age of Onset: 50-54 years<br>Survival (months, Sx onset to death): Not available<br>Site of Onset: Spinal<br>CBS: Cognition and behaviour normal | None |
| I_39 | 689658 | *NUDT2* | Diagnosis: ALS<br>El Escorial: Definite<br>Age of Onset: 75-79 years<br>Survival (months, Sx onset to death): 44.42<br>Site of Onset: Spinal<br>BBI: None<br>ECAS: None | First-degree dementia<br>First-degree alcoholism |
| I_40 | 2136466 | *NEMF* | Diagnosis: ALS<br>El Escorial: Possible<br>Age of Onset: 30-34 years<br>Survival (months, Sx onset to death): 85.28<br>Site of Onset: Spinal<br>BBI: None<br>ECAS: None | Second-degree multiple sclerosis<br>Third-degree depression and suicide<br>First-degree other neuropsychiatric<br>Second-degree alcoholism |
| AA_68 | 196723 | *TTN* | Diagnosis: ALS<br>El Escorial: Possible<br>Age of Onset: 65-69 years<br>Survival (months, Sx onset to death): Not available<br>Site of Onset: Spinal<br>CBS: Behaviour normal, ALSci | First-degree multiple sclerosis<br>First-degree autism and bipolar disorder |
| AA_48 | 3376522 | *RFC4* | Diagnosis: ALS<br>El Escorial: Probable<br>Age of Onset: 55-59 years<br>Survival (months, Sx onset to death): Not available<br>Site of Onset: Spinal<br>CBS: Behaviour normal, ALSci | First-degree Parkinson disease<br>2 x first-degree ALS |
| AA_69 | 830327 | *VWA1* | Diagnosis: Progressive Spinal Muscular Atrophy<br>El Escorial: Possible<br>Age of Onset: 45-49 years | None |

| | | | | |
|---|---|---|---|---|
| | | | Survival (months, Sx onset to death): Not available<br>Site of Onset: Spinal<br>CBS: Behaviour normal, Cognition indicates possible FTD | |
| AA_70 | 391712 | *DST* | Diagnosis: ALS<br>El Escorial: Definite<br>Age of Onset: 55-59 years<br>Survival (months, Sx onset to death): 37.45<br>Site of Onset: Cognitive<br>CBS: ALSbi, ALSci | First-degree Alzheimer's Disease<br>First-degree unknown psychiatric disorder |
| AA_71 | 847629 | *DST* | Diagnosis: ALS<br>El Escorial: Possible<br>Age of Onset: 50-54 years<br>Survival (months, Sx onset to death): 46.94<br>Site of Onset: Bulbar and Spinal<br>CBS: Behaviour indicates possible FTD, ALSci | None |
| AA_72 | 976691 | *DNAJC30* | Diagnosis: ALS<br>El Escorial: Probable<br>Age of Onset: 65-69 years<br>Survival (months, Sx onset to death): 19.02<br>Site of Onset: Spinal<br>CBS: Not performed | None |
| Hereditary Spastic Paraplegia Variants | | | | |
| I_15 | 13513 | *POLG* | Diagnosis: ALS<br>El Escorial: Possible<br>Age of Onset: 45-49 years<br>Survival (months, Sx onset to death): 41.2<br>Site of Onset: Bulbar<br>BBI: Mild changes<br>ECAS: Normal | First-degree alcoholism |
| I_16 | 13513 | *POLG* | Diagnosis: ALS<br>El Escorial: Definite<br>Age of Onset: 70-74 years<br>Survival (months, Sx onset to death): 17.61<br>Site of Onset: Spinal<br>BBI: Mild changes | Not performed |

| | | | | |
|---|---|---|---|---|
| | | | ECAS: Normal | |
| AA_31 | 13513 | *POLG* | Diagnosis: ALS<br>El Escorial: Definite<br>Age of Onset: 45-49 years<br>Survival (months, Sx onset to death): 105.35<br>Site of Onset: Spinal<br>CBS: Cognition and behaviour normal | None |
| AA_32 | 13513 | *POLG* | Diagnosis: ALS<br>El Escorial: Definite<br>Age of Onset: 40-44 years<br>Survival (months, Sx onset to death): 126.35<br>Site of Onset: Spinal<br>CBS: Cognition not performed behaviour normal | None |
| AA_33 | 13513 | *POLG* | Diagnosis: ALS<br>El Escorial: Possible<br>Age of Onset: 50-54 years<br>Survival (months, Sx onset to death): 20.17<br>Site of Onset: Bulbar<br>CBS: Cognition and behaviour normal | First-degree dementia<br>Second-degree Parkinson disease<br>First-degree obsessive-compulsive disorder<br>Second-degree depression |
| AA_34 | 13513 | *POLG* | Diagnosis: ALS<br>El Escorial: Definite<br>Age of Onset: 55-59 years<br>Survival (months, Sx onset to death): Not available<br>Site of Onset: Spinal<br>CBS: Not performed | First-degree ALS<br>First-degree Parkinson disease |
| AA_35 | 13513 | *POLG* | Diagnosis: ALS<br>El Escorial: Definite<br>Age of Onset: 65-69 years<br>Survival (months, Sx onset to death): 22.5<br>Site of Onset: Spinal<br>CBS: Not performed | First-degree alcoholism<br>First-degree multiple sclerosis |
| I_17 | 13496 | *POLG* | Diagnosis: ALS<br>El Escorial: Definite<br>Age of Onset: 40-44 years<br>Survival (months, Sx onset to death): 24.24<br>Site of Onset: Spinal | Second-degree ALS<br>First-degree depression and alcoholism<br>2 x first-degree alcoholism<br>Second-degree alcoholism<br>Second-degree alcoholism |

| | | | | |
|---|---|---|---|---|
| | | | BBI: Not performed<br>ECAS: Not performed | |
| AA_36 | 13496 | *POLG* | Diagnosis: ALS<br>El Escorial: Possible<br>Age of Onset: 60-64 years<br>Survival (months, Sx onset to death): 26.25<br>Site of Onset: Spinal<br>CBS: Cognition and behaviour normal | None |
| I_21 | 206528 | *POLG* | Diagnosis: ALS<br>El Escorial: Definite<br>Age of Onset: 45-49 years<br>Survival (months, Sx onset to death): 45.73<br>Site of Onset: Spinal<br>BBI: None<br>ECAS: None | Not available |
| I_22 | 596302 | *SPG11* | Diagnosis: ALS<br>El Escorial: Definite<br>Age of Onset: 40-44 years<br>Survival (months, Sx onset to death): 161.86<br>Site of Onset: Spinal<br>BBI: Severe changes<br>ECAS: Normal | Third-degree alcoholism and depression<br>Second-degree Parkinson disease |
| I_23 | 1705300 | *AMPD2* | Diagnosis: ALS<br>Elescorial: Definite<br>Age of Onset: 70-74 years<br>Survival (months, Sx onset to death): 22.34<br>Site of Onset: Spinal<br>BBI: Normal<br>ECAS: Normal | First-degree alcoholism<br>First-degree depression |
| I_24 | 217269 | *SPG7* | Diagnosis: ALS<br>El Escorial: Definite<br>Age of Onset: 60-64 years<br>Survival (months, Sx onset to death): 53.65<br>Site of Onset: Spinal (UMN predominant)<br>BBI: Mild changes<br>ECAS: Normal | Not provided |

| | | | | |
|---|---|---|---|---|
| I_25 | 411675 | *SPG7* | Diagnosis: ALS<br>El Escorial: Possible<br>Age of Onset: 60-64 years<br>Survival (months, Sx onset to death): 55.58<br>Site of Onset: Spinal<br>BBI: Normal<br>ECAS: Abnormal | Second-degree dementia<br>First-degree other neurological |
| I_26 | 6107 | *CYP7B1* | Diagnosis: ALS<br>El Escorial: Possible<br>Age of Onset: 40-44 years<br>Survival (months, Sx onset to death): 29.47<br>Site of Onset: Bulbar<br>BBI: Normal<br>ECAS: None | First-degree Parkinson disease<br>Second-degree dementia<br>Second-degree Parkinson disease |
| I_27 | 375313 | *AP5Z1* | Diagnosis: ALS<br>El Escorial: Definite<br>Age of Onset: 55-59 years<br>Survival (months, Sx onset to death): 111.6<br>Site of Onset: Spinal (UMN predominant)<br>BBI: None<br>ECAS: None | First-degree likely ALS<br>Third-degree certain ALS |
| I_28 | 234924 | *AP4S1* | Diagnosis: ALS<br>El Escorial: Possible<br>Age of Onset: 60-64 years<br>Survival (months, Sx onset to death): 63.34<br>Site of Onset: Spinal<br>BBI: Normal<br>ECAS: Abnormal | Third-degree bipolar disorder<br>First-degree bipolar disorder<br>Second-degree alcoholism<br>First-degree dementia |
| I_29 | 810060 | *AP5Z1* | Diagnosis: ALS<br>El Escorial: Possible<br>Age of Onset: 55-59 years<br>Survival (months, Sx onset to death): 21.12<br>Site of Onset: Spinal (monomelic)<br>BBI: Mild changes<br>ECAS: Normal | Second-degree Parkinson’s Disease<br>Third-degree multiple sclerosis<br>Third-degree other neurological<br>Second-degree suicide |
| I_30 | 450738 | *AP4M1* | Diagnosis: ALS | 2 Paternal third-degree ALS |

| | | | | |
|---|---|---|---|---|
| | | | El Escorial: Definite<br>Age of Onset: Unknown<br>Survival (months, Sx onset to death): 14.29<br>Site of Onset: Bulbar<br>BBI: None<br>ECAS: None | Third-degree depression |
| I_31 | 214335 | *FARS2* | Diagnosis: ALS<br>El Escorial: Possible<br>Age of Onset: 70-74 years<br>Survival (months, Sx onset to death): 35.94<br>Site of Onset: Spinal<br>BBI: None<br>ECAS: None | Third-degree multiple sclerosis<br>First-degree Parkinson disease |
| I_32 | 579997 | *FARS2* | Diagnosis: ALS<br>El Escorial: Probable<br>Age of Onset: 45-49 years<br>Survival (months, Sx onset to death): 42.31<br>Site of Onset: Spinal<br>BBI: None<br>ECAS: None | Third-degree Parkinson disease<br>Third-degree multiple sclerosis<br>Second-degree alcoholism |
| AA_47 | 13502 | *POLG* | Diagnosis: ALS<br>El Escorial: Definite<br>Age of Onset: 60-64 years<br>Survival (months, Sx onset to death): 27.76<br>Site of Onset: Bulbar<br>CBS: Behaviour indicates possible FTD, Cognition indicates possible FTD | Third-degree ALS |
| AA_48 | 13502 | *POLG* | Diagnosis: ALS<br>El Escorial: Probable<br>Age of Onset: 55-59 years<br>Survival (months, Sx onset to death): Not available<br>Site of Onset: Spinal<br>CBS: Behaviour normal, ALSci | First-degree Parkinson disease<br>2 x first-degree ALS |
| AA_49 | 13502 | *POLG* | Diagnosis: ALS<br>El Escorial: Definite<br>Age of Onset: 60-64 years | First-degree ALS |

| | | | | |
|---|---|---|---|---|
| | | | Survival (months, Sx onset to death): 17.58<br>Site of Onset: Bulbar<br>CBS: Behaviour not performed, Cognition possible FTD | |
| AA_50 | 13502 | *POLG* | Diagnosis: ALS<br>El Escorial: Possible<br>Age of Onset: 55-59 years<br>Survival (months, Sx onset to death): Not available<br>Site of Onset: Spinal<br>CBS: Behaviour normal, Cognition normal | Third-degree schizophrenia<br>Second-degree schizophrenia |
| AA_51 | 1437494 | *AP4M1* | Diagnosis: ALS<br>El Escorial: Probable<br>Age of Onset: 75-79 years<br>Survival (months, Sx onset to death): 17.38<br>Site of Onset: Spinal<br>CBS: Behaviour normal, ALSci | First-degree Alzheimer's disease |
| AA_52 | 156414 | *AP4B1* | Diagnosis: ALS<br>El Escorial: Probable<br>Age of Onset: 50-54 years<br>Survival (months, Sx onset to death): Not available<br>Site of Onset: Spinal<br>CBS: Behaviour not performed, Cognition normal | First-degree alcoholism |
| AA_53 | 214192 | *MTRFR* | Diagnosis: ALS<br>Elescorial: Probable<br>Age of Onset: 45-49 years<br>Survival (months, Sx onset to death): Not available<br>Site of Onset: Spinal<br>CBS: Behaviour not performed, Cognition normal | None |
| AA_54 | 219448 | *SPG7* | Diagnosis: ALS<br>Elescorial: Probable<br>Age of Onset: 55-59 years<br>Survival (months, Sx onset to death): 43.69<br>Site of Onset: Spinal<br>CBS: Not performed | Second-degree suicide<br>First-degree dementia |

| | | | | |
|---|---|---|---|---|
| AA_55 | 2906297 | *PNPLA6* | Diagnosis: ALS<br>El Escorial: Definite<br>Age of Onset: 50-54 years<br>Survival (months, Sx onset to death): 54.73<br>Site of Onset: Spinal<br>CBS: Behaviour and cognition normal | Second-degree dementia<br>Second-degree Parkinson disease |
| AA_56 | 422933 | *DDHD1* | Diagnosis: ALS<br>El Escorial: Probable<br>Age of Onset: 70-74 years<br>Survival (months, Sx onset to death): Not available<br>Site of Onset: Spinal<br>CBS: Behaviour normal, ALSci | None |
| AA_57 | 465174 | *SPG7* | Diagnosis: ALS<br>El Escorial: Definite<br>Age of Onset: 60-64 years<br>Survival (months, Sx onset to death): 43.4<br>Site of Onset: Bulbar<br>CBS: ALSbi, ALSci | Second-degree Alzheimer's disease |
| AA_58 | 6816 | *SPG7* | Diagnosis: ALS<br>El Escorial: Definite<br>Age of Onset: 55-59 years<br>Survival (months, Sx onset to death): 43.33<br>Site of Onset: Spinal<br>CBS: Behaviour and cognition indicate possible FTD | Second-degree dementia<br>First-degree dementia<br>2 x second-degree ALS |
| AA_59 | 6819 | *SPG7* | Diagnosis: ALS<br>El Escorial: Probable<br>Age of Onset: 45-49 years<br>Survival (months, Sx onset to death): Not available<br>Site of Onset: Bulbar<br>CBS: Behaviour indicates possible FTD, ALSci | 3 x first-degree depression and anxiety<br>First-degree depression and suicide |
| AA_60 | 6819 | *SPG7* | Diagnosis: ALS<br>El Escorial: Probable<br>Age of Onset: 40-44 years<br>Survival (months, Sx onset to death): 30.98<br>Site of Onset: Spinal | Second-degree dementia |

| | | | | |
|---|---|---|---|---|
| | | | CBS: Not performed | |
| AA_61 | 6819 | *SPG7* | Diagnosis: ALS<br>El Escorial: Probable<br>Age of Onset: 60-64 years<br>Survival (months, Sx onset to death): Not available<br>Site of Onset: Bulbar<br>CBS: Behaviour normal, Cognition normal | Third-degree ALS |
| AA_62 | 930104 | *FA2H* | Diagnosis: ALS<br>El Escorial: Possible<br>Age of Onset: 45-49 years<br>Survival (months, Sx onset to death): 20.04<br>Site of Onset: Bulbar<br>CBS: Not performed | Not provided |
| Ataxia Variants | | | | |
| I_41 | 162016 | *ANO10* | Diagnosis: ALS<br>El Escorial: Definite<br>Age of Onset: 60-64 years<br>Survival (months, Sx onset to death): 19.38<br>Site of Onset: Bulbar<br>BBI: None<br>ECAS: None | Not provided |
| I_42 | 162016 | *ANO10* | Diagnosis: ALS<br>El Escorial: Definite<br>Age of Onset: 70-74 years<br>Survival (months, Sx onset to death): 6.64<br>Site of Onset: Bulbar<br>BBI: None<br>ECAS: None | 2 x second-degree multiple sclerosis |
| I_43 | 162016 | *ANO10* | Diagnosis: ALS<br>El Escorial: Probable<br>Age of Onset: 65-69 years<br>Survival (months, Sx onset to death): 76.94<br>Site of Onset: Spinal<br>BBI: None<br>ECAS: None | First-degree alcoholism |

| | | | | |
|---|---|---|---|---|
| I_44 | 162016 | *ANO10* | Diagnosis: ALS<br>El Escorial: Probable<br>Age of Onset: 55-59 years<br>Survival (months, Sx onset to death): 79.96<br>Site of Onset: Spinal<br>BBI: None<br>ECAS: None | Not provided |
| I_45 | 977147 | *MSTO1* | Diagnosis: ALS<br>El Escorial: Probable<br>Age of Onset: 70-74 years<br>Survival (months, Sx onset to death): 27.07<br>Site of Onset: Spinal<br>BBI: Normal<br>ECAS: None | Not provided |
| I_46 | 3778778 | *RNF216* | Diagnosis: ALS<br>El Escorial: Definite<br>Age of Onset: 65-69 years<br>Survival (months, Sx onset to death): 18.04<br>Site of Onset: Bulbar<br>BBI: None<br>ECAS: None | Third-degree certain ALS<br>Second-degree dementia<br>Second-degree dementia<br>Third-degree suicide |
| AA_63 | 13507 | *POLG* | Diagnosis: ALS<br>El Escorial: Definite<br>Age of Onset: 45-49 years<br>Survival (months, Sx onset to death): Not available<br>Site of Onset: Spinal<br>CBS: Behaviour not performed, Cognition normal | Second-degree multiple sclerosis |
| AA_64 | 2443782 | *RNU12* | Diagnosis: ALS<br>El Escorial: Suspected<br>Age of Onset: 40-44 years<br>Survival (months, Sx onset to death): Unknown<br>Site of Onset: Spinal<br>CBS: ALSci, Behaviour not performed | None |
| Alzheimer's Disease Variants | | | | |
| I_18 | 3626 | *MPO* | Diagnosis: ALSFTD<br>El Escorial: Definite | First-degree ALS |

| | | | | |
|---|---|---|---|---|
| | | | Age of Onset: 60-64 years<br>Survival (months, Sx onset to death): 54.3<br>Site of Onset: Spinal and Cognitive/Behavioural<br>BBI: Abnormal<br>ECAS: Abnormal | |
| I_19 | 3626 | *MPO* | Diagnosis: ALS<br>El Escorial: Probable<br>Age of Onset: 75-79 years<br>Survival (months, Sx onset to death): 22.7<br>Site of Onset: Spinal<br>BBI: Severe changes<br>ECAS: Abnormal initially | Second-degree alcoholism<br>Third-degree alcoholism |
| I_20 | 3626 | *MPO* | Diagnosis: ALS<br>El Escorial: Probable<br>Age of Onset: 55-59 years<br>Survival (months, Sx onset to death): 58.77<br>Site of Onset: Bulbar<br>BBI: Not performed<br>ECAS: Normal | First-degree depression<br>First-degree alcoholism<br>First-degree alcoholism<br>2 x second-degree other neuropsychiatric<br>Second-degree suicide and IVDA |
| AA_37 | 3626 | *MPO* | Diagnosis: ALS<br>El Escorial: Suspected<br>Age of Onset: 50-54 years<br>Survival (months, Sx onset to death): Not available<br>Site of Onset: Spinal<br>CBS: Cognition normal behaviour not performed | None |
| AA_38 | 3626 | *MPO* | Diagnosis: ALS<br>El Escorial: Probable<br>Age of Onset: 65-69 years<br>Survival (months, Sx onset to death): 34.33<br>Site of Onset: Spinal<br>CBS: Cognition and behaviour normal | None |
