## Supplementary material for "Rare neurological and neurodevelopmental variants in ALS link to onset, survival and family history": S4

Supplemental Tables Navigation:

Table Supplemental Table 4.1: Neurodevelopmental Variant Information

Table 4.2: Individual Neurodevelopmental Variant Carrier Information

### Supplemental Table 4.1. Neurodevelopmental associated variants identified in Irish (n=12) and AnswerALS (n=20) pwALS.

ClinVar IDs are listed with molecular consequence, inheritance patterns for associated genes from OMIM (AR: autosomal recessive; AD: autosomal dominant), variant associated phenotypes, and population allele frequencies. Population frequency values reported from TOPMed, gnomAD, and 1000 Genomes Project (1000G) are provided where available. Cohort-specific allele frequencies were calculated assuming 2 alleles per individual using the following denominators: Irish cases (n=469, 938 alleles), Irish controls (n=230, 460 alleles), AnswerALS cases (n=774; 1548 alleles), AnswerALS controls (n=94; 188 alleles). All pwALS carrying variants in this dataset were heterozygotes. Case-level clinical and neuropsychological characteristics for these variant carriers can be cross-referenced in Supplemental Table 4.2.

| ClinVar Variant ID | Gene | Molecular Consequence | Inheritance Pattern for associated Gene | Associated Phenotype with Variant | Population Frequency | Irish Case ID | AnswerALS Case ID |
|---|---|---|---|---|---|---|---|
| 224824 | *ACP6* | Missense OR non-coding transcript variant | No Mendelian inheritance reported in OMIM (phenotype association confirmed in GeneCards) | Primary phenotype considered in current analysis:<br>Intellectual Disability<br><br>Other phenotypes associated with this variant:<br>Cerebral visual impairment | TOPMed: 0.00014<br>gnomAD: 0.00012<br>1000G: –<br>Irish ALS: 0.00213<br>AnswerALS ALS: 0<br>Irish Controls: 0<br>AnswerALS Controls: 0 | I_1<br>I_47 | -- |
| 30544 | *PIGL* | missense | AR | Primary phenotype considered in current analysis:<br>Intellectual Disability<br><br>Other phenotypes associated with this variant:<br>CHIME syndrome,<br>Bilateral cleft lip and palate<br>Camptodactyly of finger<br>Hypertelorism<br>Hypoplasia of scrotum | TOPMed: 0.00049<br>gnomAD: 0.00061<br>1000G: 0.00060<br>Irish ALS: 0.0011<br>AnswerALS ALS: 0<br>Irish Controls: 0<br>AnswerALS Controls: 0 | I_48 | -- |

| | | | | | | | |
|---|---|---|---|---|---|---|---|
| | | | | Low-set ears<br>Postaxial hand polydactyly<br>Premature birth<br>Wide intermammillary distance | | | |
| 285471 | *POMT2* | frameshift | AR | Primary phenotype considered in current analysis:<br>Intellectual Disability<br><br>Other phenotypes associated with this variant:<br>Autosomal recessive limb-girdle muscular dystrophy type<br>Muscular dystrophy-dystroglycanopathy (congenital with brain and eye anomalies),<br>Muscular dystrophy-dystroglycanopathy (congenital with brain and eye anomalies), type | TOPMed: --<br>gnomAD v4:<br>chr14-77280458-G-GT<br>Joint Max Group AF<br>0.00000803 (NFE)<br>Genomes Max Group AF<br>0.00000488 (NFE)<br>Exomes Max Group AF<br>0.00000731 (NFE)<br>1000G: --<br>Irish ALS: 0.0011<br>AnswerALS ALS: 0<br>Irish Controls: 0<br>AnswerALS Controls: 0 | I_49 | -- |
| 449095 | *SBDS* | nonsense | AR | Primary phenotype considered in current analysis:<br>Intellectual Disability<br><br>Other phenotypes associated with this variant:<br>Shwachman-Diamond syndrome, Agenesis of permanent teeth<br>Deeply set eye; Microcephaly; Short stature; Splenomegaly | TOPMed: -<br>gnomAD: 0.00099<br>1000G: 0.00060<br>Irish ALS: 0.0011<br>AnswerALS ALS: 0<br>Irish Controls: 0<br>AnswerALS Controls: 0 | I_50 | -- |

| | | | | | | | |
|---|---|---|---|---|---|---|---|
| 1805898 | *PIGG* | nonsense | AR | Primary phenotype considered in current analysis:<br>Intellectual disability, autosomal recessive<br><br>Other phenotypes associated with this variant:<br>-- | TOPMed: -<br>gnomAD v4:<br>chr4-521134-C-G<br>Joint Max Group AF<br>0.00000359 (NFE)<br>Exomes Max Group AF<br>0.00000381 (NFE)<br>1000G: --<br>Irish ALS: 0.0011<br>AnswerALS ALS: 0<br>Irish Controls: 0<br>AnswerALS Controls: 0 | I_51 | -- |
| 234924 | *AP4S1* | nonsense | AR | Primary phenotype considered in current analysis:<br>Intellectual Disability<br><br>Other phenotypes associated with this variant:<br>Hereditary spastic paraplegia | TOPMed: 0.00011<br>gnomAD: 0.00009<br>1000G: 0.00020<br>Irish ALS: 0.0011<br>AnswerALS ALS: 0<br>Irish Controls: 0<br>AnswerALS Controls: 0 | I_28 | -- |
| 1526304 | *LSS* | missense | AR | Primary phenotype considered in current analysis:<br>Intellectual Disability<br><br>Other phenotypes associated with this variant:<br>Alopecia-intellectual disability syndrome<br>Cataract<br>Hypotrichosis | TOPMed: 0.00013<br>gnomAD: 0.00015<br>1000G: --<br>Irish ALS: 0.0011<br>AnswerALS ALS: 0<br>Irish Controls: 0<br>AnswerALS Controls: 0.0053 | I_52 | -- |

| | | | | | | | |
|---|---|---|---|---|---|---|---|
| 266079 | *FAM98C* | Nonsense or intron variant | No Mendelian inheritance reported in OMIM (phenotype association confirmed in SFARI Gene) | Primary phenotype considered in current analysis:<br>Autism Spectrum Disorder<br><br>Other phenotypes associated with this variant:<br>Asphyxiating thoracic dystrophy | TOPMed: 0.00003<br>gnomAD: 0.00004<br>1000G: 0.00100<br>Irish ALS: 0.0011<br>AnswerALS ALS: 0<br>Irish Controls: 0<br>AnswerALS Controls: 0 | I_53 | -- |
| 56591 | *POMGNT1*<br><br>*TSPAN1* | Frameshift or intron variant | AR | Primary phenotype considered in current analysis:<br>Intellectual Disability<br><br>Other phenotypes associated with this variant:<br>Autosomal recessive limb-girdle muscular dystrophy<br>Muscular dystrophy-dystroglycanopathy (congenital with brain and eye anomalies),<br>type<br>Retinitis pigmentosa | TOPMed: --<br>gnomAD v4:<br>chr1-46189476-AC-A<br>Joint Max Group AF<br>0.00000878 (NFE)<br>Genomes Max Group AF<br>0.00000488 (NFE)<br>Exomes Max Group AF<br>0.0000081 (NFE)<br>1000G: --<br>Irish ALS: 0.0011<br>AnswerALS ALS: 0<br>Irish Controls: 0<br>AnswerALS Controls: 0 | I_54 | -- |
| 836262 | *AFG2A* | nonsense | AR | Primary phenotype considered in current analysis:<br>Intellectual Disability<br><br>Other phenotypes associated with this variant:<br>Microcephaly-intellectual disability-sensorineural hearing loss-epilepsy-abnormal muscle tone syndrome | TOPMed: 0.00004<br>gnomAD: 0.00004<br>1000G: --<br>Irish ALS: 0.0011<br>AnswerALS ALS: 0<br>Irish Controls: 0<br>AnswerALS Controls: 0 | I_55 | -- |

| | | | | | | | |
|---|---|---|---|---|---|---|---|
| 2678004 | *POMT1* | frameshift | AR | Primary phenotype considered in current analysis:<br>Intellectual Disability<br><br>Other phenotypes associated with this variant:<br>Autosomal recessive limb-girdle muscular dystrophy type<br>Muscular dystrophy-dystroglycanopathy (congenital with intellectual disability),<br>Walker-Warburg congenital muscular dystrophy | TOPMed: -<br>gnomAD v4:<br>chr9-131515439-G-GC<br>Joint Max Group AF<br>0.00000953 (NFE)<br>Exomes Max Group AF<br>0.00000931 (NFE)<br>1000G: --<br>Irish ALS: 0.0011<br>AnswerALS ALS: 0<br>Irish Controls: 0<br>AnswerALS Controls: 0 | I_56 | -- |
| 56578 | *POMGNT1*<br><br>*TSPAN1* | splice acceptor | AR | Primary phenotype considered in current analysis:<br>Intellectual Disability<br><br>Other phenotypes associated with this variant:<br>Autosomal recessive limb-girdle muscular dystrophy type<br>Muscular dystrophy-dystroglycanopathy (congenital with intellectual disability), type; POMGNT1-related disorder; Muscular dystrophy-dystroglycanopathy (congenital with brain and eye anomalies), type | TOPMed: --<br>gnomAD: 0.00001<br>1000G: --<br>Irish ALS: 0<br>AnswerALS ALS: 0.00065<br>Irish Controls: 0<br>AnswerALS Controls: 0 | -- | AA_73 |
| 959238 | *SLC35A3* | Nonsense or intron variant | AR | Primary phenotype considered in current analysis:<br>Autism Spectrum Disorder<br><br>Other phenotypes associated with this variant:<br>Autism spectrum disorder - epilepsy - arthrogryposis syndrome | TOPMed: 0.00000<br>gnomAD: 0.00000<br>1000G: --<br>Irish ALS: 0<br>AnswerALS ALS: 0.00065<br>Irish Controls: 0<br>AnswerALS Controls: 0 | -- | AA_74 |

| | | | | | | | |
|---|---|---|---|---|---|---|---|
| 60545 | *GMPPB* | missense | AR | Primary phenotype considered in current analysis: Intellectual Disability<br><br>Other phenotypes associated with this variant: Muscular dystrophy-dystroglycanopathy (congenital with intellectual disability); Autosomal recessive limb-girdle muscular dystrophy; Abnormality of the musculature | TOPMed: 0.00023<br>gnomAD: 0.00019<br>1000G: --<br>Irish ALS: 0<br>AnswerALS ALS: 0.00065<br>Irish Controls: 0<br>AnswerALS Controls: 0 | -- | AA_75 |
| 575991 | *GMPPB* | missense | AR | Primary phenotype considered in current analysis: Intellectual Disability<br><br>Other phenotypes associated with this variant: Autosomal recessive limb-girdle muscular dystrophy; Muscular dystrophy-dystroglycanopathy (congenital with brain and eye anomalies)<br>Muscular dystrophy-dystroglycanopathy (congenital with intellectual disability); Abnormality of the musculature | TOPMed: 0.00008<br>gnomAD: 0.00004<br>1000G: --<br>Irish ALS: 0<br>AnswerALS ALS: 0.00065<br>Irish Controls: 0<br>AnswerALS Controls: 0 | -- | AA_76 |

| | | | | | | | |
|---|---|---|---|---|---|---|---|
| 60546 | *GMPPB* | missense | AR | Primary phenotype considered in current analysis: Intellectual Disability<br><br>Other phenotypes associated with this variant: Autosomal recessive limb-girdle muscular dystrophy Muscular dystrophy-dystroglycanopathy (congenital with brain and eye anomalies); Muscular dystrophy-dystroglycanopathy (congenital with intellectual disability); Muscular dystrophy | TOPMed: 0.00049<br>gnomAD: 0.00053<br>1000G: --<br>Irish ALS: 0<br>AnswerALS ALS: 0.00065<br>Irish Controls: 0<br>AnswerALS Controls: 0 | -- | AA_5 |
| 476318 | *PIGG* | nonsense | AR | Primary phenotype considered in current analysis: Intellectual disability, autosomal recessive<br><br>Other phenotypes associated with this variant:<br>-- | TOPMed: 0.00059<br>gnomAD: 0.00073<br>1000G: 0.00040<br>Irish ALS: 0<br>AnswerALS ALS: 0.0013<br>Irish Controls: 0.0065<br>AnswerALS Controls: 0 | -- | AA_77<br>AA_78 |
| 286126 | *PIGO* | Frameshift or intron variant | AR | Primary phenotype considered in current analysis: Intellectual Disability<br><br>Other phenotypes associated with this variant: Hyperphosphatasia with intellectual disability syndrome | TOPMed: --<br>gnomAD v4:<br>chr9-35092076-C-CG<br>Joint Max Group AF<br>0.00036947 (NFE)<br>Genomes Max Group AF<br>0.000182 (NFE)<br>Exomes Max Group AF<br>0.00037599 (NFE)<br>1000G: --<br>Irish ALS: 0<br>AnswerALS ALS: 0.0013 | -- | AA_79<br>AA_80 |

| | | | | | | | |
|---|---|---|---|---|---|---|---|
| | | | | | Irish Controls: 0<br>AnswerALS Controls: 0 | | |
| 617475 | *EIF3F* | missense | AR | Primary phenotype considered in current analysis:<br>Intellectual Disability<br>Neurodevelopmental Disorder<br><br>Other phenotypes associated with this variant:<br>Intellectual developmental disorder, autosomal recessive<br>Intellectual developmental disorder, autosomal recessive<br>Neurodevelopmental disorder with dysmorphic facies and distal limb anomalies | TOPMed: 0.00077<br>gnomAD: 0.00130<br>1000G: 0.00016<br>Irish ALS: 0<br>AnswerALS ALS: 0.0013<br>Irish Controls: 0<br>AnswerALS Controls: 0 | -- | AA_81<br>AA_82 |
| 1262 | *RNASEH2B* | missense | AR | Primary phenotype considered in current analysis:<br>Autism Spectrum Disorder<br><br>Other phenotypes associated with this variant:<br>Abnormality of the nervous system; Hereditary spastic paraplegia; Cerebral palsy; Aicardi-Goutieres syndrome | TOPMed: 0.00150<br>gnomAD: 0.00141<br>1000G: 0.00016<br>Irish ALS: 0<br>AnswerALS ALS: 0.00194<br>Irish Controls: 0.0065<br>AnswerALS Controls: 0 | -- | AA_83<br>AA_84<br>AA_12 |

| | | | | | | | |
|---|---|---|---|---|---|---|---|
| 321216 | *PMM2* | splice donor | AR | Primary phenotype considered in current analysis:<br>Intellectual Disability<br><br>Other phenotypes associated with this variant:<br>*PMM2*-congenital disorder of glycosylation | TOPMed: 0.00009<br>gnomAD: 0.00007<br>1000G: --<br>Irish ALS: 0<br>AnswerALS ALS: 0.00065<br>Irish Controls: 0<br>AnswerALS Controls: 0 | -- | AA_85 |
| 30544 | *PIGL* | missense | AR | Primary phenotype considered in current analysis:<br>Intellectual Disability<br><br>Other phenotypes associated with this variant:<br>CHIME syndrome; Bilateral cleft lip and palate<br>Camptodactyly of finger<br>Hypertelorism<br>Hypoplasia of scrotum<br>Low-set ears<br>Postaxial hand polydactyly<br>Premature birth<br>Wide intermamillary distance | TOPMed: 0.00049<br>gnomAD: 0.00041<br>1000G: 0.00047<br>Irish ALS: 0<br>AnswerALS ALS: 0.0013<br>Irish Controls: 0<br>AnswerALS Controls: 0 | -- | AA_86<br>AA_87 |
| 224645 | *PGAP3* | intron variant | AR | Primary phenotype considered in current analysis:<br>Intellectual Disability<br><br>Other phenotypes associated with this variant:<br>Hyperphosphatasia with intellectual disability syndrome | TOPMed: 0.00011<br>gnomAD: 0.00009<br>1000G: 0.00016<br>Irish ALS: 0<br>AnswerALS ALS: 0.00065<br>Irish Controls: 0<br>AnswerALS Controls: 0 | -- | AA_88 |
| 4847 | *PNKP* | frameshift | AR | Primary phenotype considered in current analysis:<br>Intellectual Disability<br><br>Other phenotypes associated with this variant: | TOPMed: --<br>gnomAD v4:<br>chr19-49861800-T-TGTTGTCGATGGCGACCC<br>Joint Max Group AF 0.00035574 (NFE) | -- | AA_89<br>AA_90 |

| | | | | | |
|---|---|---|---|---|---|
| | | | | Microcephaly, seizures, and developmental delay; Ataxia - oculomotor apraxia; Developmental and epileptic encephalopathy; Abnormality of the nervous system; Seizure | Genomes Max Group AF 0.0002672 (NFE)<br>Exomes Max Group AF 0.000355 (NFE)<br>1000G: --<br>Irish ALS: 0<br>AnswerALS ALS: 0.0013<br>Irish Controls: 0<br>AnswerALS Controls: 0 |

### Supplemental Table 4.2.

Neurodevelopmental associated ClinVar variants identified in Irish ALS cohort (n=12) and AnswerALS datasets (n=20). Anonymised IDs, ClinVar Variant IDs, associated genes, ALS clinical phenotype including neuropsychological testing, and family history of neurological or psychiatric disease are displayed per variant carrier. Neuropsychological testing for Irish cohort included Beaumont Behavioural Inventory (BBI) and the Edinburgh Cognitive and Behavioural ALS Screen (ECAS), while cognitive assessment in the AnswerALS cohort utilised the ALS Cognitive and Behavioural Screen (CBS). Any abnormal cognitive/behavioural result recorded at any timepoint during disease course is indicated in the phenotype column. For CBS classification. ALSci, ALS with cognitive impairment; ALSbi, ALS with behavioural impairment.

| Case ID | ClinVar Variant ID | Gene | Phenotype | Family History |
|---|---|---|---|---|
| Intellectual Disability Variants | | | | |
| I_1 | 224824 | *ACP6* | Diagnosis: ALS<br>El Escorial: Probable<br>Age of Onset: 50-54 years<br>Survival (months, Sx onset to death): 41.29<br>Site of Onset: Bulbar<br>BBI: Normal<br>ECAS: Abnormal | 2 x third-degree ALS<br>Third-degree Parkinson disease |
| I_47 | 224824 | *ACP6* | Diagnosis: ALS<br>El Escorial: Not provided<br>Age of Onset: 55-59 years<br>Survival (months, Sx onset to death): 26.02<br>Site of Onset: Bulbar<br>BBI: not performed<br>ECAS: not performed | Third-degree ALS<br>First-degree Parkinson disease<br>First-degree bipolar disorder<br>First-degree suicide |
| I_48 | 30544 | *PIGL* | Diagnosis: ALS<br>El Escorial: Definite<br>Age of Onset: 50-54 years<br>Survival (months, Sx onset to death): 9.79 | Second-degree bipolar disorder<br>5 x second-degree alcoholism |

| | | | | |
|---|---|---|---|---|
| | | | Site of Onset: Bulbar<br>BBI: not performed<br>ECAS: not performed | |
| I_49 | 285471 | *POMT2* | Diagnosis: ALS<br>El Escorial: Definite<br>Age of Onset: 80-84 years<br>Survival (months, Sx onset to death): 25.13<br>Site of Onset: Spinal<br>BBI: not performed<br>ECAS: not performed | None |
| I_50 | 449095 | *SBDS* | Diagnosis: ALS<br>El Escorial: Definite<br>Age of Onset: 75-79 years<br>Survival (months, Sx onset to death): 17.74<br>Site of Onset: Bulbar<br>BBI: normal<br>ECAS: Abnormal | First-degree depression<br>First-degree depression<br>First-degree bipolar disorder<br>Second-degree alcoholism<br>2 x third-degree autism |
| I_51 | 1805898 | *PIGG* | Diagnosis: ALS<br>El Escorial: Definite<br>Age of Onset: 75-79 years<br>Survival (months, Sx onset to death): 16.23<br>Site of Onset: Bulbar<br>BBI: not performed<br>ECAS: not performed | Not performed |
| I_28 | 234924 | *AP4S1* | Diagnosis: ALS<br>El Escorial: possible<br>Age of Onset: 60-64 years<br>Survival (months, Sx onset to death): 63.34<br>Site of Onset: Spinal<br>BBI: Normal<br>ECAS: Abnormal | First-degree bipolar disorder<br>First-degree dementia<br>Second-degree alcoholism<br>Third-degree bipolar disorder |
| I_52 | 1526304 | *LSS* | Diagnosis: ALS<br>El Escorial Definite<br>Age of Onset: 65-69 years<br>Survival (months, Sx onset to death): 15.7<br>Site of Onset: Spinal<br>BBI: Mild changes<br>ECAS: Normal | 2 x first-degree dementia<br>First-degree alcoholism |
| I_54 | 56591 | *POMGNT1*<br>*TSPAN1* | Diagnosis: ALS<br>El Escorial: Definite<br>Age of Onset: 60-64 years<br>Survival (months, Sx onset to death): 30.52 | Second-degree dementia |

| | | | | |
|---|---|---|---|---|
| | | | Site of Onset: Bulbar<br>BBI: Not performed<br>ECAS: Not performed | |
| I_55 | 836262 | *AFG2A* | Diagnosis: ALS<br>El Escorial: Probable<br>Age of Onset: 65-69 years<br>Survival (months, Sx onset to death): 19.78<br>Site of Onset: Spinal<br>BBI: Normal<br>ECAS: Normal | 2 x first-degree Alzheimer's disease<br>First-degree with autism |
| I_56 | 2678004 | *POMT1* | Diagnosis: ALS<br>El Escorial: Definite<br>Age of Onset: 65-69 years<br>Survival (months, Sx onset to death): 31.47<br>Site of Onset: Spinal<br>BBI: Severe changes<br>ECAS: Normal | Second-degree alcoholism<br>Second-degree alcoholism |
| AA_73 | 56578 | *POMGNT1*<br>*TSPAN1* | Diagnosis: ALS<br>El Escorial: Definite<br>Age of Onset: 55-59 years<br>Survival (months, Sx onset to death): 38.7<br>Site of Onset: Spinal<br>CBS: Not performed | None |
| AA_75 | 60545 | *GMPPB* | Diagnosis: ALS<br>El Escorial: Probable<br>Age of Onset: 45-49 years<br>Survival (months, Sx onset to death): Unknown<br>Site of Onset: Spinal<br>CBS: ALSci, Behaviour normal | None |
| AA_76 | 575991 | *GMPPB* | Diagnosis: ALS<br>El Escorial: Possible<br>Age of Onset: 70-74 years<br>Survival (months, Sx onset to death): 16.8<br>Site of Onset: Bulbar<br>CBS: Not performed | First-degree dementia<br>First-degree alcoholism |
| AA_5 | 60546 | *GMPPB* | Diagnosis: ALS<br>El Escorial: Probable<br>Age of Onset: 50-54 years<br>Survival (months, Sx onset to death): Unknown<br>Site of Onset: Bulbar<br>CBS: ALSci, Behaviour normal | First-degree Alzheimer's disease |
| AA_77 | 476318 | *PIGG* | Diagnosis: ALS | Second-degree Alzheimer's disease |

| | | | | |
|---|---|---|---|---|
| | | | El Escorial: Definite<br>Age of Onset: 60-64 years<br>Survival (months, Sx onset to death): Unknown<br>Site of Onset: Bulbar<br>CBS: ALSci, Behaviour normal | |
| AA_78 | 476318 | *PIGG* | Diagnosis: ALS<br>El Escorial: Probable<br>Age of Onset: 60-64 years<br>Survival (months, Sx onset to death): 35.3<br>Site of Onset: Spinal<br>CBS: Cognition normal, Behaviour not performed | Not performed |
| AA_79 | 286126 | *PIGO* | Diagnosis: ALS<br>El Escorial: Definite<br>Age of Onset: 75-79 years<br>Survival (months, Sx onset to death): Unknown<br>Site of Onset: Bulbar<br>CBS: ALSci, Behaviour normal | None |
| AA_80 | 286126 | *PIGO* | Diagnosis: ALS<br>El Escorial: Probable<br>Age of Onset: 65-69 years<br>Survival (months, Sx onset to death): 64.2<br>Site of Onset: Bulbar<br>CBS: Cognition normal, Behaviour not performed | None |
| AA_81 | 617475 | *EIF3F* | Diagnosis: ALS<br>El Escorial: Possible<br>Age of Onset: 55-59 years<br>Survival (months, Sx onset to death): 24.3<br>Site of Onset: Bulbar<br>CBS: ALSci, Behaviour normal | Second-degree Alzheimer's disease<br>First-degree dementia |
| AA_82 | 617475 | *EIF3F* | Diagnosis: ALS<br>El Escorial: Probable<br>Age of Onset: 60-64 years<br>Survival (months, Sx onset to death): Unknown<br>Site of Onset: Spinal<br>CBS: Cognition normal, Behaviour not performed | None |
| AA_85 | 321216 | *PMM2* | Diagnosis: ALS<br>El Escorial: Possible<br>Age of Onset: 50-54 years<br>Survival (months, Sx onset to death): Unknown | First-degree bipolar disorder<br>2 x second-degree Alzheimer's disease |

| | | | | |
|---|---|---|---|---|
| | | | Site of Onset: Spinal<br>CBS: ALSci, Behaviour normal | |
| AA_86 | 30544 | *PIGL* | Diagnosis: ALS<br>El Escorial: Possible<br>Age of Onset: 65-69 years<br>Survival (months, Sx onset to death): 32.8<br>Site of Onset: Spinal<br>CBS: ALSci, Behaviour normal | Second-degree dementia |
| AA_87 | 30544 | *PIGL* | Diagnosis: ALS<br>El Escorial: Definite<br>Age of Onset: 65-69 years<br>Survival (months, Sx onset to death): Unknown<br>Site of Onset: Spinal<br>CBS: ALSci, Behaviour may indicate FTD | 2 First-degree ALS<br>Second-degree dementia |
| AA_88 | 224645 | *PGAP3* | Diagnosis: ALS<br>El Escorial: Probable<br>Age of Onset: 70-74 years<br>Survival (months, Sx onset to death): Unknown<br>Site of Onset: Spinal<br>CBS: Cognition normal, Behaviour not performed | First-degree alcoholism |
| AA_89 | 4847 | *PNKP* | Diagnosis: ALS<br>El Escorial: Definite<br>Age of Onset: 60-64 years<br>Survival (months, Sx onset to death): 33.0<br>Site of Onset: Spinal<br>CBS: Cognition and Behaviour normal | First-degree Alzheimer's disease |
| AA_90 | 4847 | *PNKP* | Diagnosis: ALS<br>El Escorial: Definite<br>Age of Onset: 65-69 years<br>Survival (months, Sx onset to death): Unknown<br>Site of Onset: Spinal<br>CBS: ALSci, Behaviour normal | None |
| Autism Spectrum Disorder Variants | | | | |
| I_53 | 266079 | *FAM98C* | Biological Sex: Male<br>Diagnosis: ALS<br>El Escorial Definite<br>Age of Onset: 55-59 years<br>Survival (months, Sx onset to death): 25.82<br>Site of Onset: Spinal<br>BBI: Not performed<br>ECAS: Not performed | 2 x second-degree with alcoholism<br>Second-degree dementia |

| | | | | |
|---|---|---|---|---|
| AA_74 | 959238 | *SLC35A3* | Diagnosis: ALS<br>El Escorial: Probable<br>Age of Onset: 65-69 years<br>Survival (months, Sx onset to death): 40.5<br>Site of Onset: Spinal<br>CBS: ALSci, Behaviour normal | None |
| AA_83 | 1262 | *RNASEH2B* | Diagnosis: ALS<br>El Escorial: Probable<br>Age of Onset: 45-49 years<br>Survival (months, Sx onset to death): Unknown<br>Site of Onset: Bulbar<br>CBS: Cognition normal, Behaviour not performed | None |
| AA_84 | 1262 | *RNASEH2B* | Diagnosis: ALS<br>El Escorial: Probable<br>Age of Onset: 65-69 years<br>Survival (months, Sx onset to death): 36.3<br>Site of Onset: Spinal<br>CBS: Not performed | First-degree dementia |
| AA_12 | 1262 | *RNASEH2B* | Diagnosis: ALS<br>El Escorial: Probable<br>Age of Onset: 55-59 years<br>Survival (months, Sx onset to death): 37.5<br>Site of Onset: Spinal<br>CBS: Cognition and Behaviour normal | First-degree Parkinson disease and dementia |
